# Genome-wide association study meta-analysis identifies susceptibility loci informing Ewing sarcoma etiology and potential mechanisms of risk

**DOI:** 10.64898/2026.02.06.26345779

**Authors:** Aubrey K. Hubbard, Hélène Neyret-Kahn, Martina Müller-Nurasiyd, Dominik Löw, Konstantin Strauch, Olivia W. Lee, Andrew R. Raduski, Tianzhong Yang, Weiyin Zhou, Elliot W. Stratton, Orson Jay, Sandrine Grossetête, Andrew J. Song, Diptavo Dutta, Amy A. Hutchinson, Belynda D. Hicks, Michelle Manning, Jia Liu, Carrie Boyce, Wolfgang Hartmann, Uta Dirksen, Andreas E. Kulozik, Markus Metzler, Manuela Krumbholz, Alexander Teumer, Henry Völzke, Uwe Völker, Joshua D. Schiffman, Javed Khan, Melissa M. Hudson, Kirsten K. Ness, Zhaoming Wang, Katherine A. Janeway, Philip J. Lupo, Logan G. Spector, Wen-Yi Huang, Steven C. Moore, Stephen J. Chanock, Thomas G.P. Grünewald, Olivier Delattre, Mitchell J. Machiela, the Ewing Sarcoma Germline Genetics Group

**Author notes:** These authors share senior authorship (Thomas G.P. Grünewald, Olivier Delattre, Mitchell J. Machiela).

## Abstract

Ewing sarcoma (EwS) is a rare, aggressive pediatric malignancy driven by *FET*::*ETS* family fusions (*EWSR1*::*FLI1* in >85% of cases) with no established environmental risk factors. To investigate germline predisposition, we analyzed 2,014 EwS cases and 10,525 cancer-free controls in a two-stage analysis that combined an international genome-wide association study and a case□parent trio study. The combined meta-analysis identified 18 variants at 14 susceptibility loci (9 novel, 5 replicated) with moderate effect sizes (odds ratios≥1.25). Integrative analyses of the EwS loci revealed enrichment of expanded GGAA microsatellites, with evidence for binding of the EWSR1::FLI1 chimeric oncogenic activator. *EWSR1::ETS* knockdown in EwS cell lines resulted in dysregulated genes at susceptibility loci related to skeletal/muscle development, RNA binding/processing, and chromatin regulation. Our findings provide insights into the inherited component of EwS, highlighting a genetic architecture in which common germline variations with moderate effects interact with somatic *EWSR1*::*FLI1* fusions to promote sarcomagenesis by dysregulating local genes.

## Introduction

Ewing sarcoma (EwS) is a rare, aggressive bone and soft tissue cancer that primarily occurs in adolescents and young adults. International incidence studies show the highest rates in Western countries^1^, with EwS most common in individuals genetically similar to European reference samples (1.5 cases per million) and significantly rarer in individuals genetically similar to African or East Asian reference samples (0.2 and 0.8 cases per million, respectively)^2^. The ancestral difference suggests a distinctive genetic component of EwS susceptibility. EwS also demonstrates a modest male predominance (risk ratio ∼ 1.5:1), indicating potential sex-linked genetic factors for EwS susceptibility^2^. EwS primarily affects adolescents and young adults, with a peak incidence between the ages of 10 and 20, implicating developmental and hormonal factors in its pathogenesis^2,3^. The five-year survival rate for EwS patients is 70% overall but less than 30% for patients with metastatic disease at diagnosis, emphasizing the critical need for advancements in early detection and treatment strategies^4,5^.

EwS is defined by a chromosomal translocation most commonly of *EWSR1* but also of *FUS* and *TAF15,* with a member of the erythroblast transformation-specific (ETS) transcription factor family, most commonly Friend leukemia integration 1 (*FLI1)*^6^. Pathognomonic *EWSR1::FLI1* fusion, present in more than 85% of cases, drives extensive epigenetic remodeling. In particular, EWSR1::FLI1 can bind otherwise nonfunctional GGAA microsatellites (GGAA-mSat) and convert them into neoeshancers, with the number of GGAA-mSat repeats (i.e., their length) positively correlated with enhancer activity^7,8^. Compared with other cancers, particularly pediatric cancers, EwS has a low somatic mutation rate (0.15 mutations/Mb of coding sequence)^9,10^ and lacks established evidence for rare germline pathogenic or likely pathogenic variants driving risk^11^. However, recent studies have reported potential enrichment of inherited pathogenic variants or structural alterations in DNA damage repair genes, including *FANCC*, in EwS patients^12,13^. Prior GWAS have identified six common independent EwS susceptibility variants across five genetic loci with moderate effect sizes (odds ratios (ORs) >1.7), highlighting a distinctive genetic architecture in which common variants are predisposed to a rare malignancy^14,15^. Some of these germline susceptibility loci modulate EWSR1::FLI1 activity through variation in GGAA-mSat length^8,16^, resulting in altered EWSR1::FLI1 binding affinity and downstream dysregulation of local gene expression.

To further investigate the genetic etiology of EwS, we performed a meta-analysis of eight international genome-wide association studies (GWAS) and a case□parent trio study combining 2,014 EwS cases and 10,525 cancer-free controls. We also conducted case-only analyses to investigate potential differences in germline risk associated with important clinical features. To gain deeper mechanistic insights into the interplay between genetic risk and tumor biology, we conducted integrated analyses of identified loci utilizing publicly available datasets on EwS cell lines and patient tumors, including data on GGAA-mSat length, chromatin conformation, EWSR1::FLI1 binding, and tumor gene expression. Our investigation identified germline regions harboring GGAA-mSats that correlate with EwS risk. Accordingly, these findings underscore the important oncogenic activity of EWSR1::FLI1 binding and provide new insights into how the germline can inform and interact with the fusion oncodrivers critical for EwS.

## Subjects and Methods

### Overview of the study

We conducted a meta-analysis of 2,014 cases of EwS and 10,525 cancer-free controls. EwS cases for the international GWAS meta-analysis were obtained from the Institut Curie (IC), the Childhood Cancer Survivor Study (CCSS)^17^, the Center for Cancer Research (CCR) at the National Cancer Institute (NCI), the NCI Bone Disease and Injury Study^18^, the Children’s Oncology Group (COG), Michigan Neonatal Biobank^19^, Cooperative Ewing Sarcoma Study (CESS) (https://ewing.uk-essen.de/), and the St. Jude Lifetime Cohort Study (SJLIFE)^20^. EwS cancer-free controls were obtained from the American Cancer Society Prevention Study II (ACS), the Spanish Bladder Cancer Study (SBCS), the NCI-SEER Non-Hodgkin Lymphoma Case□Control Study (NHL)^21^, the Prostate, Lung, Colorectal, and Ovarian Screening Trial (PLCO)^22,23^, and the Study of Health in Pomerania - TREND (SHIP-TREND)^24^ (**Supplemental Table 2**). The participants for the EwS TDT analyses were part of Gabriela Miller Kid’s First (GMKF)^25^.

### Genotype quality control and imputation

Blood-derived DNA samples for COG, Institut Curie (batch 3), Michigan blood spots, and PLCO were genotyped by the National Cancer Institute (NCI) using the Illumina Global Screening Array (GSA). Case-control datasets were provided in six sets based on genotyping array: (1) IC GWAS 1^14^, (2) CCSS cases and matched controls, (3) NCI samples and IC samples with matched controls, (4) COG, Michigan blood spot and IC samples matched with PLCO controls, (5) CESS and SHIP-TREND controls, and (6) SJLIFE and matched controls. Pre-imputation genotyped sets were merged using only the overlapping variants. Marker QC was performed separately within each group. Imputation was not performed on SJLIFE cases or controls, as whole-genome sequencing (WGS) data were used. Imputation was conducted to the Trans-Omics for Precision Medicine (TOPMed)^26^ version r3 reference panel using the TOPMed Imputation Server for all imputed sets except CESS, which was locally imputed to 1000 Genomes phase 3^27^ using the Michigan imputation server (v1.2.7)^28^.

Genotypes derived from WGS for 860 individuals (287 probands, 573 parents) were obtained from the GMKF Pediatric Research Program in Susceptibility to Ewing Sarcoma Based on Germline Risk and Familial History of Cancer (phs001228.v2.p2). The process of analyzing raw sequencing reads to produce called genomic variants was performed using established GMKF protocols^29^ based on GATK best practices prior to downloading the data. Genomic intervals containing variants in linkage disequilibrium with index SNPs (see Table 1 ‘LD Region’) in the discovery dataset were extracted from individual-level genomic variant call format (gVCF) files. The genotypes for the resulting 24,645 variants were then merged into a single variant call format file and converted into binary plink file format. Pedigree and phenotypic data were encoded manually.

**Table 1.**
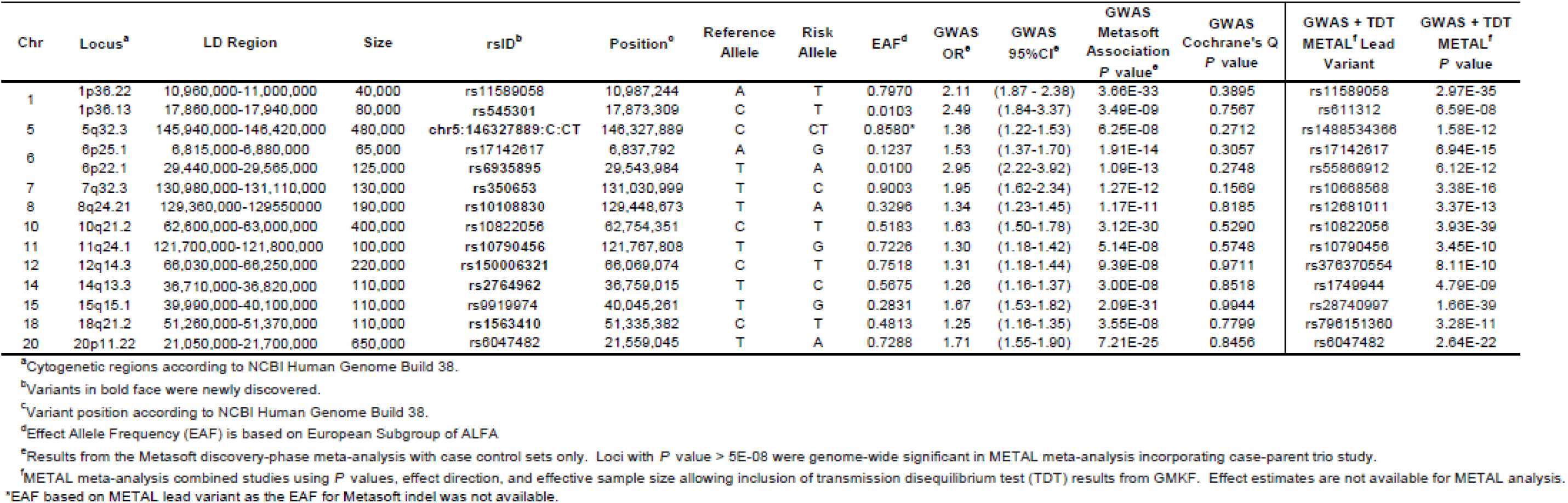
Meta-analysis results for genetic susceptibility to Ewing sarcoma. Results display GW AS fixed effects analysis using Metasoft on the case-control datasets and the results adding the case-trio set to the case-control results by pooling P values and weighting by effective sample size in METAL. GWAS was conducted at a 1% minor allele threshold.

**Table 2.** Lead variant results for sex-stratified case:control analysis and case-only by sex analysis. Case-only analysis represents the statistical effect of the lead variant’s association with male sex among Ewing (EwS) cases.

| Chr | Locus | rsID | Position | Male Case:Control |  |  | Female Case:Control |  |  | Sex Case-Only |
| --- | --- | --- | --- | --- | --- | --- | --- | --- | --- | --- |
|  |  |  |  | OR | 95%CI | P value for EwS | OR | 95%CI | P value for EwS | P value for sex as outcome |
| 1 | 1p36.22 | rs11589058 | 10,987,244 | 2.04 | (1.61-2.56) | 4.40E-09 | 1.96 | (1.51-2.50) | 3.00E-07 | 0.5779 |
| 5 | 5q32.3 | chr5:146327889:C:CT | 146,327,889 | 1.30 | (1.06-1.59) | 0.0123 | 1.31 | (1.06-1.61) | 0.0129 | 0.8273 |
| 6 | 6p25.1 | rs17142617 | 6,837,792 | 1.50 | (1.21-1.86) | 2.30E-04 | 1.48 | (1.19-1.83) | 3.10E-04 | 0.6318 |
| 7 | 7q32.3 | rs350653 | 131,030,999 | 1.82 | (1.28-2.56) | 7.10E-04 | 2.04 | (1.39-2.94) | 2.49E-04 | 0.8363 |
| 8 | 8q24.21 | rs10108830 | 129,448,673 | 1.33 | (1.13-1.56) | 4.40E-04 | 1.27 | (1.07-1.50) | 0.0062 | 0.1881 |
| 10 | 10q21.2 | rs10822056 | 62,754,351 | 1.52 | (1.28-1.79) | 6.20E-07 | 1.72 | (1.45-2.04) | 1.03E-09 | 0.3346 |
| 11 | 11q24.1 | rs10790456 | 121,767,808 | 1.31 | (1.10-1.58) | 0.0033 | 1.30 | (1.07-1.58) | 0.0097 | 0.9974 |
| 12 | 12q14.3 | rs150006321 | 66,069,074 | 1.34 | (1.10-1.62) | 0.0029 | 1.30 | (1.07-1.59) | 0.0092 | 0.1934 |
| 14 | 14q13.3 | rs2764962 | 36,759,015 | 1.35 | (1.15-1.59) | 3.00E-04 | 1.32 | (1.11-1.56) | 0.0015 | 0.9836 |
| 15 | 15q15.1 | rs9919974 | 40,045,261 | 1.79 | (1.52-2.11) | 1.40E-12 | 1.57 | (1.32-1.86) | 2.92E-07 | 0.7532 |
| 18 | 18q21.2 | rs1563410 | 51,335,382 | 1.27 | (1.09-1.49) | 0.0025 | 1.35 | (1.14-1.59) | 4.68E-04 | 0.4235 |
| 20 | 20p11.22 | rs6047482 | 21,559,045 | 1.79 | (1.45-2.17) | 3.00E-08 | 1.69 | (1.37-2.13) | 1.64E-06 | 0.4135 |

Newly discovered variants at 1p36.13 and 6p22.1 were not included in the analysis due to low minor allele frequencies in subgroup analytic sets.
Contributing Studies and numbers for cases in each study are in **Supplemental Table 8**.

Post-imputation QC was performed across all sets. Variants with MAF < 0.01, Hardy-Weinberg Equilibrium (HWE) *P* value <□1×10^−6^ in controls, or imputation quality scores < 0.3 were removed. Samples with excess homozygosity or heterozygosity calculated within an ancestry were removed. To identify duplicate EwS cases within or across case□control sets analyzed at the NCI, one individual from each pair of related subjects (PI_HAT > 0.10) was excluded based on identity-by-descent (IBD) estimates calculated in PLINK v.1.9 using the --genome tag. For EwS genotype sets analyzed by external collaborators, common genotypes were aligned and merged with 1000 Genomes Project^27^ reference samples, and principal components were estimated using smartpca in EIGENSOFT^30^. The projection of PCs onto a common reference space enabled the detection of identical samples contributed across institutions. One sample from each identified set of identical samples was removed before analysis.

### Statistical analysis

#### Covariate adjustment PCA and matching

We performed principal component analysis (PCA) within each set (excluding the GMKF case–parent trios) for evaluation of population stratification and selection of covariates for GWAS. Plink 1.9 was used to extract 12,893 ancestry-informative markers^31^ from each genotyped set and estimate principal components using the --pc flag. Eigenvectors and eigenvalues resulting from PCA were used in PCAmatchR to select controls for each EwS case based on weighted PCs^32^. PCs were assessed against EwS status in each set, and eigenvectors displaying a potential association with EwS (*P*<0.10) were selected as covariates for analysis in each GWAS set.

#### GWAS and TDT

Logistic regression under an additive model was performed using SNPTest (V2) for CESS or PLINK 1.9 for all other case□control sets to obtain ORs and 95% CIs. Transmission disequilibrium tests (TDTs) were performed on the GMKF case–parent trio data using PLINK 1.9 --tdt flag to obtain ORs and 95% CIs for the odds of allelic transmission (transmitted vs. untransmitted allele). All *P* values were two-sided. The inflation factor for each study was evaluated individually for evidence of potential systematic inflation of statistics.

#### Meta-Analysis

A discovery phase fixed effects meta-analysis on case□control datasets with >80% European ancestry determined using grafpop^33^ was performed in Metasoft^34^, which combines case□control GWAS effect estimates (betas) and standard errors. This method was used to estimate ORs and 95% CIs for each lead variant. We then performed a meta-analysis in METAL^35^ by combining the *P* values from all studies and weighting each study’s results by the effective sample size (ESS) for each study where ESS = (4 x *case N* x *control N*) ÷ (*case N* + *control N*). We reported results for lead variants with *P* < 5×10^−8^ in the main fixed effects meta-analysis or variants just below genome-wide significance that were genome-wide significant (*P* < 5×10^−8^) in the METAL meta-analysis. All variants were evaluated for evidence of heterogeneity (Cochrane’s Q *P* < 0.05).

Among the case□control sets, we performed leave one out meta-analysis and evaluated our results in the meta-analysis removing each study. Study-specific estimates were plotted against meta-analysis results without each study to evaluate whether any study demonstrated substantial heterogeneity. We also generated forest plots of the OR and 95% CI for each lead variant in each study to evaluate differences across studies in effect direction or magnitude.

#### Conditional analysis

We performed conditional analysis using Genome-wide Complex Trait Analysis (GCTA) to identify independently associated variants at susceptibility loci with the results of our GWAS meta-analysis^36,37^. The LD for the conditional analysis was calculated using a reference population of 18,794 controls from the Prostate, Lung, Colorectal and Ovarian Screening (PLCO) trial to achieve a desirable accuracy. Variants with imputation quality <0.3 were excluded from the reference set for conditional analysis. Conditional analysis was restricted to 11 loci with lead variants that achieved genome-wide significance in the case□control GWAS meta-analysis. Conditional analysis was performed on variants with a MAF>1% within the region of 1 Mb up- and downstream of the index variant. The analysis was performed using the GCTA-COJO v1.94.1^37^, starting with the index variant at each locus and using a forward stepwise selection approach. Only variants with *P* < 5×10^−8^ in the conditional analysis were considered independently significant.

#### Case-only and subgroup analyses

To investigate the established difference EwS incidence by sex and age at diagnosis and the impact of metastatic EwS on prognosis, we examined genetic architecture by sex (male:female), metastasis at diagnosis (yes/no), and age at diagnosis. Three different categories for age at diagnosis were assessed in the case-only GWAS: 1) children (< 10) vs. adolescents and adults (≥10) to evaluate genetic susceptibility differences in children prior to the onset of puberty, 2) pre-puberty (≤15) and post-puberty (>15) by using the mean end of puberty (15), and 3) pediatric (<20) vs. non-pediatric age range (≥20). PCs were estimated for EwS cases. We conducted case-only analysis for each dataset with available data on sex, age, and metastasis, adjusting for PCs associated (*P*<0.10) with the trait of interest. Study-specific effects were meta-analyzed in a fixed effects meta-analysis using Metasoft^34^. Additional GWAS was performed among males and females to evaluate potential differences in subgroups.

#### Integration with Ewing sarcoma cell line atlas (ESCLA) published data

Published data from the ESCLA^38^ were utilized for integrated analyses of all discovered susceptibility loci. ESCLA data have been described elsewhere^38^. Briefly, the ESCLA comprises 18 EwS cell lines with inducible *EWSR1::ETS* (*EWSR1::FLI1* or *EWSR1::ERG*, respectively) knockdown (EETS KD) and whole-genome, DNA methylation, transcriptome, proteome, and chromatin immunoprecipitation sequencing (ChIP-seq) data. A GGAA microsatellite (mSat) track was included to assess whether mSats reside in or near the discovered EwS susceptibility loci where the EWSR1::ETS fusion protein could bind. ChIP-seq in A-673/TR/shEF1 (a clone of A-673) EwS cell lines under normal conditions and *EWSR1::ETS* KD were used to evaluate open chromatin, marked by H3K27ac, as well as FLI1 or ERG binding. We further incorporated chromatin conformation under normal and EETS-KD conditions to identify potential connections between de novo enhancers at EWSR1::ETS binding sites and nearby genes^39^.

The UCSC genome browser was used to visualize each discovered locus, with the discovery phase GWAS results overlaid with tracks from the ESCLA detailing additional features. To formally evaluate the statistical enrichment of the discovered loci to GGAA-mSats and FLI1 binding, we utilized R to randomly sample 10,000 variants with replacement on each chromosome from the GWAS meta-analysis. The distance between each variant and consecutive GGAA-mSats or high FLI1/ERG binding was calculated to create a null distribution. The distance between consecutive GGAA-mSats or FLI1/ERG binding and the lead variant in the discovery phase GWAS was compared against the sample distribution on the lead variant’s chromosome to estimate the proportion of values that are less than or equal in distance to mSats or FLI1 compared with the lead variant. In previous studies, the EWSR1::FLI1 oncoprotein bound to a minimum of four consecutive GGAA-mSats and displayed exponential increases in EWSR1::FLI1-dependent enhancer activity at greater than 12 consecutive repeats^8,38,40,41^; thus, GGAA statistical enrichment was evaluated at both thresholds.

#### Gene expression analysis with EETS KD

To investigate the possible impact of *EWSR1::ETS* on gene expression near GWAS-identified loci, we selected candidate genes located 1 Mb up- or downstream of the lead GWAS variants and present in ESCLA (n=207)^38^. Using Affymetrix gene expression data from 18 EwS cell lines with dox-inducible KD of *EWSR1::FLI1* (15 cell lines) or *EWSR1::ERG* (3 cell lines), we examined the effects of EETS KD on gene mRNA expression. We calculated the changes in mRNA expression between the KD and EwS conditions across three replicates per cell line and averaged them for one difference per cell line. The mean expression change for each gene across all the cell lines was assessed using a one-sample t test to determine statistical significance. Multiple testing correction was performed using a Bonferroni-adjusted threshold of *P* < 2.42×10^−4^ (0.05/207 genes). We applied the same method across a subset of five cell lines with higher KD efficiency for comparison.

#### eQTL analysis in EwS tumors

To test for potential expression quantitative trait loci (eQTLs) in identified EwS susceptibility loci, we evaluated available genotypes and matched tumor samples from EwS cases^14^. A total of 113 samples were evaluated on the Affymetrix chip, and 48 were evaluated using RNA-seq, with 28 tumor samples in common between the two platforms. Variants located in LD regions of discovered loci were used to test for associations with gene expression for genes 1 Mb up- or downstream (n=209 genes) of the GWAS lead variant. Filtering of nonexpressed/very low-expressed genes was performed, retaining genes with an average expression of > 2 fragments per kilobase of transcript per million reads (FPKM) for RNA-Seq and an intensity of > 2.5 for the microarray data. Association testing of the effect of genotype (number of minor allele copies) on gene expression was performed using linear regression in R. A Bonferroni correction was applied to account for multiple testing of genes, and results with *P* < 2.39×10^−4^ (0.05/209) were deemed statistically significant.

#### Physical interaction between selected genes and mSats

We evaluated the physical interaction between gene promoters and GGAA repeats using the GGAA repeat track from ESLCA and the Hi-ChIP chromatin capture data from the A-673 cell line^38,42^. Hi-ChIP data were filtered to overlap only true peaks in CTCF and H3K27ac using Model-based Analysis of ChIP-Seq (MACS2)^43^ to filter ChIP-seq for improved peak calling and retain only Hi-ChIP interactions that overlap with peaks called by MACS2 prior to the evaluation of physical interactions between gene promoters and GGAA-mSats. We conducted the analysis at a 5 kb resolution. The positions of four or more GGAA repeats within 1 Mb up- or downstream from the lead variant were used to verify the presence of interactions between the GGAA mSats and the gene promoters of the genes (N=38) identified in gene expression KD analysis and eQTL analysis. The gene promoter locations for the 5 kb bin were defined as those 1 kb upstream from the gene transcription start site.

## Results

### Study population

The studied EwS cases were from a representative case population that was predominantly male (55.9%), with a mean age at diagnosis of 15.1 years for males and 13.7 years for females (*P*=3.25×10^−4^). A total of 28.7% of patients reported metastatic disease at diagnosis, with the age at diagnosis being older for metastatic patients (mean ages 16.1 vs. 14.4 for nonmetastatic patients, *P*=0.03). Tumors were most common in the lower extremities (27.8%) and pelvis (23.0%), and tumors in the pelvis were most likely to present metastatically (*P*=5.54×10^−7^). Phenotypic characteristics, such as age at diagnosis, metastasis status and tumor location, were available for approximately half of the EwS patients (**Supplemental Table 1**).

### Meta-analysis identifies 14 EwS susceptibility loci

We performed GWAS across six independent analytic sets (comprising eight studies) of EwS cases and ancestry-matched unrelated controls of at least 80% genetic similarity to European reference samples (**Supplemental Figure 1, Supplemental Table 2**). A fixed effect meta-analysis was conducted using the resulting 1,730 EwS cases and 9,957 ancestry-matched cancer-free controls to estimate association ORs and 95% confidence intervals (CIs) for 10,853,557 total genomic variants generated by imputation. The genomic inflation factor showed minimal evidence of systematic inflation (lambda=1.03). All five previously reported EwS loci at 1p36.22, 6p25.1, 10q21.3, 15q15.1, and 20p11.22 (as well as the conditional independent association at 20p11.22) replicated, with *P* values becoming stronger with the addition of the expanded GWAS sets. The GWAS identified six new loci reaching genome-wide significance (*P*_meta_<□5×10^−8^) at 1p36.13, 6p22.1, 7q32.3, 8q24.21, 14q13.3 and 18q21.2, resulting in a total of 11 EwS loci initially reaching genome-wide significance. The effect allele frequency (EAF) ranged from 0.01 to 0.90 in European populations and did not display a clear pattern across genetic ancestry, despite EwS being predominantly observed in individuals of European descent (**Supplemental Table 3**).

Our GWAS meta-analysis also revealed three loci that have not yet reached genome-wide significance (*P*<1×10^−6^) at 5q32.3, 11p15.3 and 12q14.3 (**Table 1**). We selected the LD regions for each locus and performed a transmission disequilibrium test (TDT) for these regions in an independent set of 287 EwS cases and 573 parents. The TDT results from the trios were meta-analyzed with the case-control GWAS sets, resulting in a final meta-analysis of 2,014 EwS cases and 10,525 cancer-free controls. These three nominal loci reached genome-wide significance in the combined meta-analysis, resulting in a total of 14 EwS susceptibility loci with estimated ORs ranging from 1.25□2.49 (**Table 1**, **Figure 1**).

**Figure 1.**
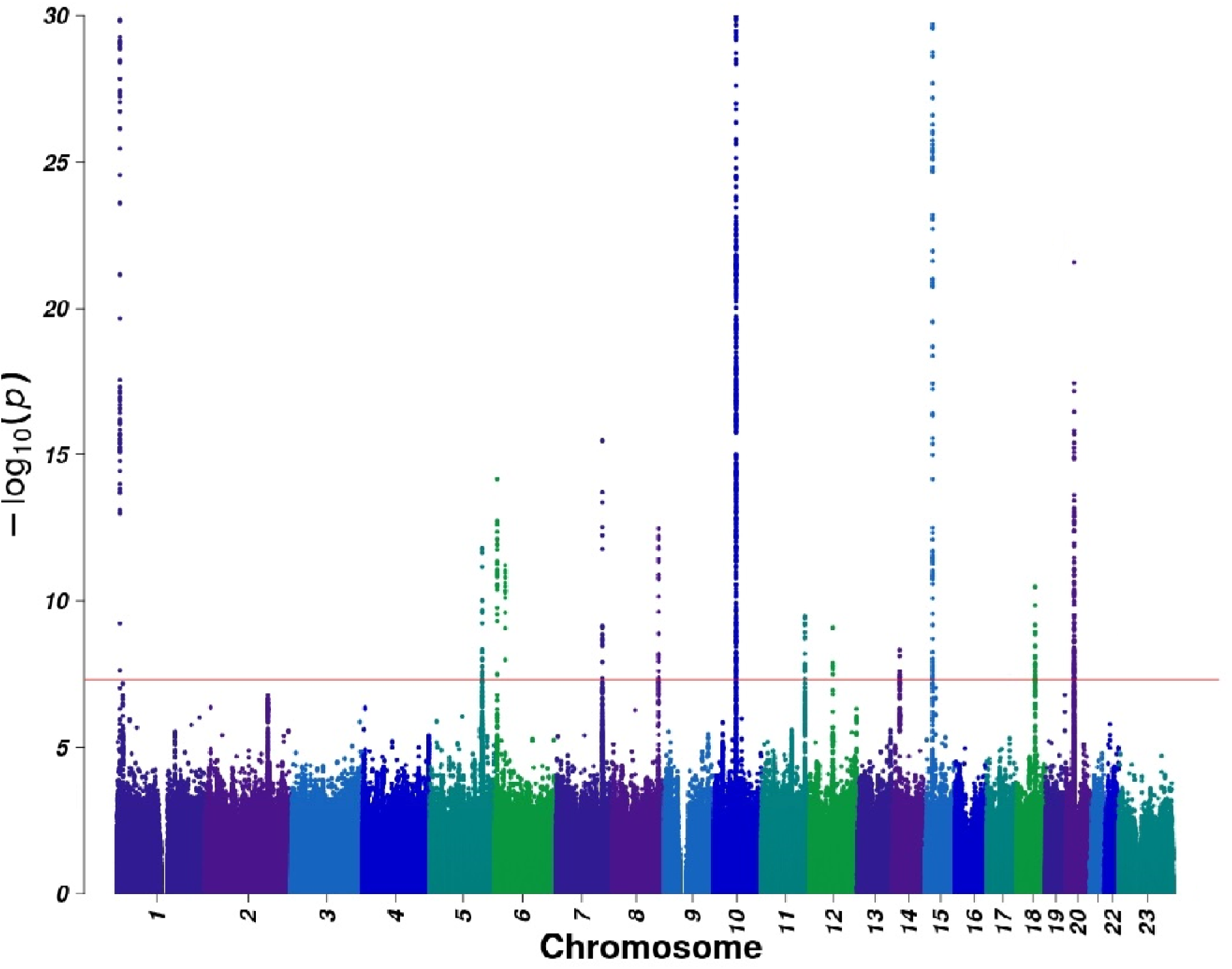
Manhattan plot of the METAL meta-analysis –log10 P values for the association of each SNP with EwS risk. Association P values for each tested genetic variant are plotted. Chromosomes are plotted sequentially across the x-axis, with the scale proportional to chromosomal size. Colors are used to visualize differences in chromosomes. The red line indicates genome-wide significance (P<5×10-8). Variants with < 1% minor allele frequency were excluded from each study’s results prior to meta-analysis. A total of 10,853,557 variants were included in the analysis. Variants with heterogeneity (Q<0.05) or present in only one study were excluded from the Manhattan plot. The estimated level of genomic inflation was lambda=1.03.

The lead variants displayed considerable consistency in effect direction and magnitude across all contributing studies (**Supplemental Table 4, Supplemental Figure 2**), and the meta-analysis results remained robust in the leave-one-out analyses **(Supplemental Figure 3, Supplemental Table 5**), with no one study driving association results for any of the EwS susceptibility signals.

The case□parent trio results were highly concordant with the case□control sets in the effect direction for the discovered loci. The TDT results revealed genome-wide replication for nine of the discovered loci. However, lead variants in the TDT analysis were not always the same as those in the discovery-phase case□control meta-analysis (**Supplemental Table 6**) but were in linkage disequilibrium (LD) (R^2^>0.25). The TDT ORs for the lead variants within the GWAS were highly correlated with the fixed effects meta-analysis (**Supplemental Figure 4**). For the low-frequency variants (minor allele frequency (MAF)∼ 1%; 1p36.13 and 6p22.1), the TDT OR was inflated or uncalculatable due to low or no un-transmitted alleles (**Supplemental Table 4**).

We performed stepwise conditional analysis using Genome-wide Complex Trait Analysis (GCTA) to identify independently associated variants at EwS susceptibility loci with the results of our GWAS meta-analysis. We identified independent variants at 6p25.1, 7q32.3, 15q15.1, and 20q22.11 (**Figure 2, Supplemental Table 7**). The confirmed conditional variant at 20q22.11 was previously reported^15^. Cumulatively, these analyses resulted in 18 independent variants across 14 loci associated with EwS susceptibility.

**Figure 2.**
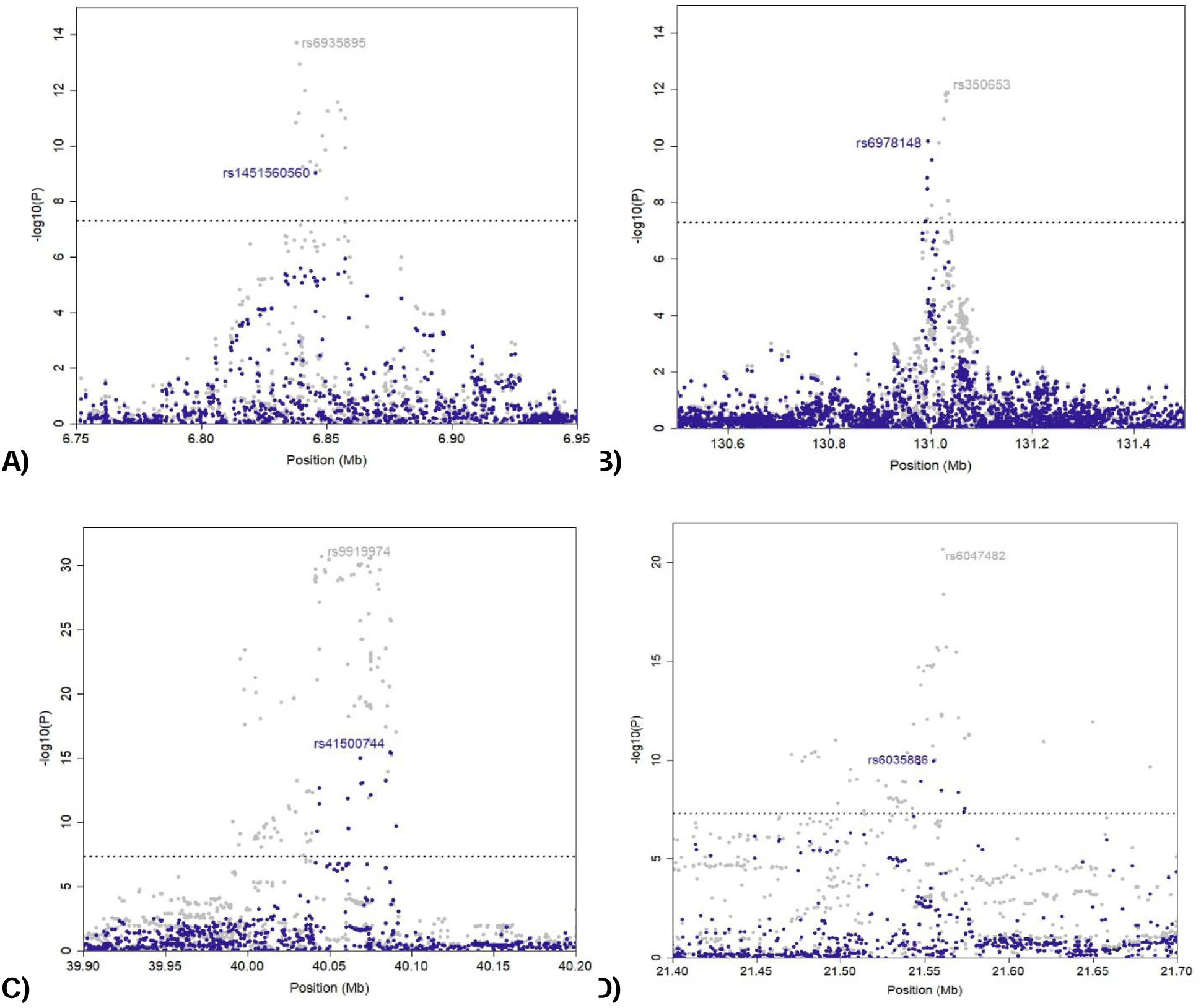
Conditional analysis revealed independent loci at the A) 6p25.1, B) 7q32.3, 15q15.1 and D) 20p11.22--20p11.23 regions. Meta-analysis -log10 P values are plotted in gray in the background. The meta-analysis -log10P values after conditioning the lead SNP in each region are plotted in navy blue. The detailed results are provided in **Supplemental Table 7.**

### Examination of EwS germline susceptibility by case characteristics

Case-only GWAS were performed to investigate potential genetic associations with EwS case characteristics and included GWAS by sex, metastasis at diagnosis, and age at diagnosis.

Although our study population displayed a 1.3:1 male-to-female ratio, there was no genome-wide significant association on the X chromosome, and the case-only GWAS by sex did not yield genome-wide significant loci (**Supplemental Figure 5**). In addition, interrogation of prior and newly discovered EwS susceptibility GWAS loci found no evidence of heterogeneity in effect by sex (**Supplemental Table 8**). Several EwS loci reached genome-wide significance in GWAS conducted within strata of sex (**Supplemental Figure 6**).

Individual-level data on metastasis at diagnosis were available for 943 EwS cases (**Supplemental Table 8**). Case-only GWAS for metastatic disease did not reveal genome-wide significant associations (**Supplemental Figure 7**); although, suggestive associations (*P*<1×10^−6^) were observed at 1p21.2 and 3p24.1, near *VCAM1* and *LLRC3B*, respectively.

We also examined age at EwS diagnosis using three distinct age categorizations: children (0□9 years) vs. adolescents and adults (≥10 years), children aged 15 years or younger vs. teenagers and adults (> 15 years), and children aged 0□19 years vs. adults (≥ 20). No genome-wide significant evidence for associations in the case-only GWAS was identified (**Supplemental Figure 8**), but two nominal signals (*P*<1×10^−6^) were observed. For the first age categorization, a locus at 10q23.1, near *RPA2P2,* displayed a suggestive association. For the second age categorization, a nominal signal was detected at 15q13.1, proximal to *GOLGAM8*.

### Integration with EwS cell line and patient gene expression data provides etiologic insights

To investigate the underlying mechanism of action at each discovered EwS susceptibility locus, we generated integrative plots for each of the 14 loci with meta-analysis summary statistics and publicly available data from 18 EwS cell lines from the EwS Cell Line Atlas (ESCLA)^38^, including available Hi-C^44^ and Hi-ChIP^42^ data performed on the ESCLA cell line A-673. The resulting plots overlay genomic features with genes in the region (**Figure 3**, **Extended Data**). Upon visualization, 11 (79%) of the 14 EwS susceptibility loci had nearby GGAA-mSat peaks and open chromatin marked by H3K27ac. Ten of the 14 EwS loci presented evidence of EWSR1::FLI1 binding that aligns with these peaks. To test these relationships formally, we performed statistical enrichment tests for GGAA-mSats and EWSR1::FLI1 binding by comparing the proximity of the identified lead GWAS variants to randomly sampled variants on the same chromosome. The distance to four consecutive GGAA-mSats was significantly (empirical *P*<0.05) lower for eight of the lead variants than for 10,000 randomly selected GWAS variants on the same chromosome (**Supplemental Table 9**). Additionally, six of these loci were also significantly closer to mSats with more than 12 consecutive GGAA-mSats, where exponential increases in *EWSR1::FLI1 or EWSR1::ERG*-dependent enhancer activity have been reported.^8,38,40,41^ A similar pattern was observed with enrichment for EWSR1::FLI1 binding, which was detected for nine of the lead variants (**Supplemental Table 9**). The loci with low-frequency lead variants (MAFs∼0.01; 1p36.13 and 6p22.1) and the 5q32.3 locus were outliers and did not display statistical enrichment in proximity to EWSR1::FLI1 binding or GGAA-mSats at any threshold, suggesting that these loci may not have direct relationships with regional EWSR1::FLI1 binding. Visualization of additional ESCLA cell lines confirmed that the absence of EWSR1::FLI1 or EWSR1::ERG binding at rare variant loci and at 5q32.3 was not specific to the A-673 cell line or EWSR1::FLI1.

**Figure 3:**
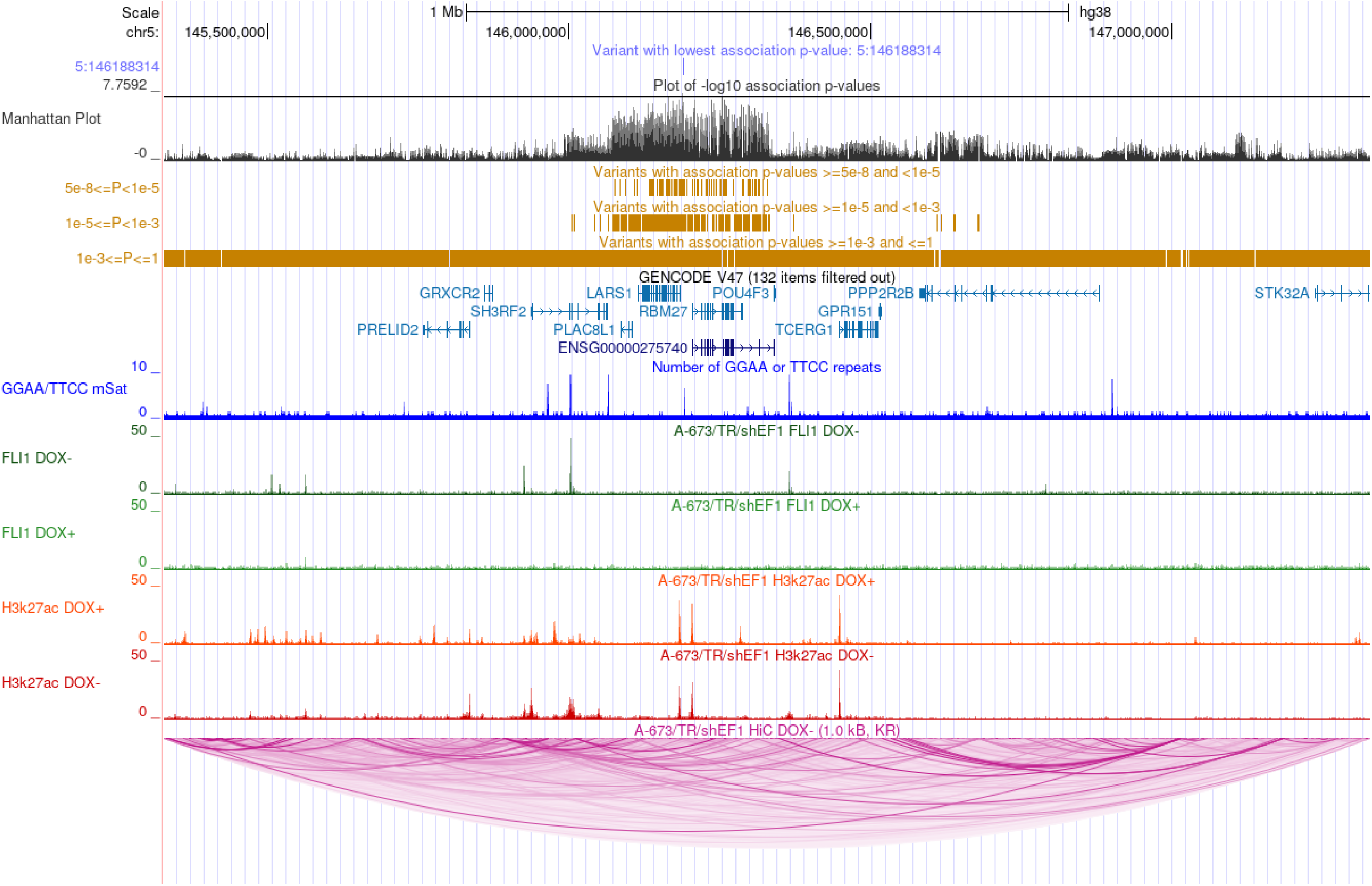
5q32.3 locus. UCSC Genome browser view of meta-analysis results and publicly available data from the EwS cell line atlas. The plot displays the meta-analysis results, genes 1 Mb up- and downstream of the lead variant, GGAA repeats, FLI1 binding, H3K27ac, and HiC loops.

To nominate genes of functional interest at identified EwS susceptibility loci, we performed expression quantitative trait locus (eQTL) analysis for genes within 1 Mb up- or downstream of the lead GWAS variant (n=209 genes) using both gene expression microarray and RNA-seq data from EwS tumors. Genes with *P* < 2.4×10^−4^ (Bonferroni corrected for 209 genes) in at least one data type (microarray or RNA-seq) were deemed statistically significant. Our results support previously published findings that GWAS risk variants are associated with increased expression of *RREB1* (6p25.1) and *EGR2* (10q21.2) (**Supplemental Table 10**)^7,14,40^. Analyses of GWAS risk alleles also identified eQTLs for decreased expression of several genes, including *CASZ1* and *TARDBP* at 1p36.22, *RBM27* and *PLAC8L1* at 5q32.3, *SLC25A21* and *PAX9* at 14q3.3, *SRP14* at 15q15.1, and *MEX3C* at 18q21.2.

We investigated the effects of *EWSR1::ETS* on the expression of genes in the vicinity of identified EwS susceptibility loci. We selected genes 1 Mb up- or downstream of the lead GWAS variant for this analysis and found 207 genes available in ESCLA for evaluation (**Supplemental Table 11**). Twenty genes from eight EwS loci showed evidence of *EWSR1::ETS* dysregulation under knockdown (KD) at a statistically significant threshold (*P*<2.4×10^−4^), with three upregulated genes and 17 downregulated genes. *PAX7* (1p36.13) was the most strongly downregulated gene (mean log_2_FC=-3.33, *P*=1.44×10^−8^), and *ARID5B* was the most strongly upregulated gene (mean log_2_FC=2.02, *P*=1.75×10^−8^).

Available Hi-ChIP^42^ data were used to evaluate evidence for potential physical interactions between GGAA-mSats near lead variants, and gene promoters for genes significant by eQTL or KD analyses were evaluated. There was evidence of physical interaction between nine gene promoters and GGAA-mSats across seven regions in H3K27ac Hi-ChIP and six genes across six regions in CTCF Hi-ChIP. To aid with the interpretation of the results from the integrative analyses, the results for each of the 14 EwS loci were aggregated into a summary table (**Supplemental Table 12**) that aggregated the results from the GGAA enrichment, EWSR1::FLI1 binding, eQTL, KD, and chromatin interaction analyses.

## Discussion

In the largest to date international meta-analysis of 2,014 EwS cases and 10,525 cancer-free controls, we expanded the knowledge of the genetic architecture of EwS with the identification of 18 variants at 14 EwS susceptibility loci. Our findings robustly replicate previously identified loci and identify nine novel EwS loci. The large effect sizes for GWAS provide evidence of a distinctive germline genetic architecture for EwS, in which common variation with moderate effect sizes predisposes individuals to a rare malignancy. The incorporation of a family-based trio study improved power and demonstrated external replication.

We provide strong evidence for EWSR1::FLI1 binding at most EwS susceptibility loci, supporting a model in which germline variation tagged by GWAS alters EWSR1::FLI1 binding affinity at GGAA-mSats through genetic variation in mSat length. This finding is consistent with published literature describing how germline variation in GGAA-mSat length can alter EWSR1::FLI1 binding.^7,8,40,41,45^ Among the 14 identified EwS susceptibility loci, 11 presented evidence for GGAA-mSats or FLI1 binding near the lead variant; however, the complexity of this germline-somatic interaction between the GGAA-mSat motif and fusion protein binding was not fully captured in our analysis. For example, a recent publication indicated highly polymorphic repeat alleles at GGAA-mSats and that the total number of GGAA repeats in the 6p25.1 region was more strongly associated with EwS risk than consecutive stretches^7^. Prior investigations also indicate that GGAA-mSats bound by EWSR1::ETS fusions have additional GGAA-mSats nearby and a low number of interspersed bases contiguous to the adjacent GGAA-mSats^38^.

These findings highlight an opportunity for new technologies, such as long-read sequencing, to characterize variation in these complex microsatellites more precisely and their functional consequences in the identified EwS susceptibility regions.

Integrated expression and chromatin analyses near EwS susceptibility loci identified candidate genes with potential etiologic roles in EwS. These results highlight the underlying genetic mechanisms important for EwS susceptibility. First, eQTL analyses provide evidence for EwS candidate genes that act on EwS risk through allele-specific changes in expression mediated through allelic effects on transcription factor binding and enhancer activity. This mechanism of risk has been described in functional investigations of adult solid malignancies and is not unique to EwS. Second, we observe evidence of a distinctive EwS risk mechanism in which GWAS risk alleles tag germline variation in GGAA-mSats that are recognized and bound by the somatically acquired EWSR1::FLI1 fusion protein. Germline length alterations of GGAA-mSats modulate the binding affinity and regulatory activity of EWSR1::FLI1 neo-enhancers, leading to changes in the transcriptional activity of nearby target genes. EwS susceptibility loci located near GGAA-mSats, which show significant eQTL associations and altered gene expression under EWSR1::FLI1 knockdown provide strong support for this mechanism of genetic risk. Evidence for genes in EwS susceptibility loci is summarized in Supplemental Table 12.

An example of this distinctive EwS risk mechanism involves genes in the paired box (PAX) family involved in skeletal and muscle development. *PAX7*, which is telomeric to the lead variant at 1p36.13, near GGAA-mSats, showed the strongest expression change following EWSR1::FLI1 KD and is highly expressed in EwS, indicating that germline–somatic interactions at 1p36.13 likely dysregulate *PAX7* transcription^46,47^. *PAX7* plays a key role in muscle development and is involved in the PAX7□FOXO1 fusions observed in alveolar rhabdomyosarcoma^48^. Interestingly, evidence from eQTL analyses suggests that *PAX9*, which is located near the susceptibility locus at 14q13.3, is involved in embryonic skeletal patterning^49^ and is overexpressed in multiple cancers; however, there is less evidence for a change in *PAX9* expression with EWSR1::FLI1 KD, and further studies are needed to confirm this observation^50^.

Additional candidate genes may play roles in RNA binding and processing. *MEX3C*, which is implicated in both eQTL and EWSR1::FLI1 KD data, encodes an RNA-binding protein linked to tumor progression in several cancers^51,52^, including osteosarcoma, where it has been linked to tumor aggressiveness^53^. *RBM27*, located near the 5q32.3 locus, also showed substantial expression changes in eQTL and EWSR1::FLI1 KD analyses. *RBM27* is predicted to participate in mRNA processing and splicing, processes frequently altered in oncogenic transcriptome remodeling^54^.

Two additional genes, *SLC25A21 and PADI2,* may act through metabolic and chromatin-related functions. *SLC25A21,* near 14q13.3, encodes a mitochondrial transporter involved in amino acid metabolism and has been reported to have expression mediated by EWSR1::FLI1 binding in chromatin capture analysis^44^. *PADI2*, at 1p36.13, exhibited consistent expression changes under KD across EwS cell lines and catalyzes the citrullination of arginine residues on histones, altering chromatin structure and gene expression^55^. *PADI2* has also been implicated in tumor proliferation; immune invasion; and the development and progression of breast, prostate, and bladder cancers^56^.

We performed EwS case-only GWAS investigations by clinical features for etiologic insights into sex, age at diagnosis, and metastatic disease. Our results did not identify evidence for a germline genetic component that explains the male predominance. There were suggestive associations for metastasis that could implicate two plausible candidate genes, *VCAM1* and *LRRC3B*. *VCAM1* has been shown to promote tumor growth, progression, and metastasis, particularly in bone tumors^57,58^. Furthermore, *VCAM1* in EwS cell lines with altered WT1 promoted angiogenesis^57,58^. *LRRC3B*, an implicated tumor suppressor gene, may play a role in the treatment response or escape immune surveillance^59,60^. However, further studies are needed to confirm these intriguing findings.

Our results expand the understanding of the inherited genetic component to EwS risk and portray a genetic architecture in which many common germline variants with moderately high effect predispose to a rare malignancy through germline–somatic interactions with *EWSR1*::*FLI1,* although some may act through *EWSR1::FLI1*-independent mechanisms. This highlights EwS as a unique model of germline–somatic interaction, as few other cancers have demonstrated such a clear dependence of germline risk on a tumor-specific somatic event. The molecular insights derived from bioinformatic analyses of EwS susceptibility loci have identified a set of plausible candidate genes and biological pathways for further study and could eventually lead to new targets for therapy or early intervention.

## Data Availability

Data from the newly genotyped individuals in the EwS GWAS are available at dbGaP under accession number (in progress). The previously genotyped data are from dbGaP under phs001549.v1.p1 for omni set and phs001327.v1.p1 for CCSS. The genotyping data of the SJLIFE (St. Jude Lifetime Cohort) study are available through the St. Jude Cloud. Data from the Gabriela Miller Kids First program are available under dbGap study accession phs001228). Requests for CESS should be addressed to T.G.P.G. and U.D. The data of the SHIP TREND study cannot be made publicly available owing to the informed consent of the study participants, but they can be accessed through a data application form available at https://transfer.ship-med.uni-greifswald.de for researchers who meet the criteria for access to confidential data.
Ewing sarcoma cell line atlas (ESCLA) data are available under the Gene Expression Omnibus (GEO) Accession series GSE176190. A-673/TR/ShEF1 ChIP-seq data were obtained from accession number GSE129155. Hi-C data are under GSE185132, and H3K27ac Hi-CHiP data are available under GSE116495. The RNA-seq data are deposited in the European Genome‒Phenome Archive (EGA) under dataset ID: EGAS00001003333. All the data were realigned on hg38.

https://transfer.ship-med.uni-greifswald.de

https://www.ncbi.nlm.nih.gov/projects/gap/cgi-bin/GetSampleStatus.cgi?study_id=phs001549.v1.p1&rettype=html

https://www.ncbi.nlm.nih.gov/projects/gap/cgi-bin/study.cgi?study_id=phs001327.v1.p1

https://gcc02.safelinks.protection.outlook.com/?url=https%3A%2F%2Fega-archive.org%2Fstudies%2FEGAS00001003333&data=05%7C02%7Caubrey.hubbard%40nih.gov%7Cf9c4edbaeedb4283928f08de0fae8c32%7C14b77578977342d58507251ca2dc2b06%7C0%7C0%7C638965439558786675%7CUnknown%7CTWFpbGZsb3d8eyJFbXB0eU1hcGkiOnRydWUsIlYiOiIwLjAuMDAwMCIsIlAiOiJXaW4zMiIsIkFOIjoiTWFpbCIsIldUIjoyfQ%3D%3D%7C0%7C%7C%7C&sdata=FvDJqpAfqkj5ROQB3KUPuGz9nfdOtBodfob3Me6FRFo%3D&reserved=0

## Acknowledgments

This research was supported in part by the Intramural Research Program of the National Institutes of Health (NIH). The contributions of the NIH author(s) were made as part of their official duties as NIH federal employees, are in compliance with agency policy requirements, and are considered works of the United States Government. However, the findings and conclusions presented in this paper are those of the author(s) and do not necessarily reflect the views of the NIH or the U.S. Department of Health and Human Services, nor does mention of trade names, commercial products, or organizations imply endorsement by the U.S. Government. This project has been funded in whole or in part with Federal funds from the National Cancer Institute, National Institutes of Health, under Control No. 75N91019D00024. The content of this publication does not necessarily reflect the views or policies of the department.

T. Yang is supported by the St. Baldrick Career Award (1463960). A.T. has been funded by the Deutsche Forschungsgemeinschaft (DFG, German Research Foundation) – 542489987. M.M.H., K.K.N., and Z.W. are partially supported by funding from the National Cancer Institute (CA195547, CA283333).

Children’s Oncology Group (COG) samples were obtained as part of the NCTN Operations Center Grant U10CA180886COG Biospecimen Bank Grant U24CA196173 WWWW Foundation, Inc. (also known as the QuadW Foundation)

The results analyzed here are based in part upon data generated by the Gabriella Miller Kids First Pediatric Research Program projects and were accessed from the Kids First Data Resource Portal (https://kidsfirstdrc.org/).

## Data and code availability

Data from the newly genotyped individuals in the EwS GWAS are available at <u>dbGaP</u> under accession number (*in progress*). The previously genotyped data are from dbGaP under <u>phs001549.v1.p1</u> for omni set and <u>phs001327.v1.p1</u> for CCSS. The genotyping data of the SJLIFE (St. Jude Lifetime Cohort) study are available through the St. Jude Cloud. Data from the Gabriela Miller Kids First program are available under dbGap study accession phs001228).

Requests for CESS should be addressed to T.G.P.G. and U.D. The data of the SHIP-TREND study cannot be made publicly available owing to the informed consent of the study participants, but they can be accessed through a data application form available at https://transfer.ship-med.uni-greifswald.de for researchers who meet the criteria for access to confidential data.

Ewing sarcoma cell line atlas (ESCLA) data are available under the Gene Expression Omnibus (GEO) Accession series GSE176190. A-673/TR/ShEF1 ChIP-seq data were obtained from accession number GSE129155. Hi-C data are under GSE185132, and H3K27ac Hi-CHiP data are available under GSE116495. The RNA-seq data are deposited in the European Genome□Phenome Archive (EGA) under dataset ID: <u>EGAS00001003333</u>. All the data were realigned on hg38.

## Code availability

No previously unreported custom computer code or algorithm was used to generate the results.

## References

1. Spector LG, Hubbard AK, Diessner BJ, Machiela MJ, Webber BR, Schiffman JD. Comparative international incidence of Ewing sarcoma 1988 to 2012. Int J Cancer. 2021;149(5):1054–1066. doi:10.1002/ijc.33674

2. Jawad MU, Cheung MC, Min ES, Schneiderbauer MM, Koniaris LG, Scully SP. Ewing sarcoma demonstrates racial disparities in incidence-related and sex-related differences in outcome: an analysis of 1631 cases from the SEER database, 1973-2005. Cancer. 2009;115(15):3526–3536. doi:10.1002/cncr.24388

3. Grünewald TGP, Cidre-Aranaz F, Surdez D, et al. Ewing sarcoma. Nat Rev Dis Primer. 2018;4(1):1–22. doi:10.1038/s41572-018-0003-x

4. Gaspar N, Hawkins DS, Dirksen U, et al. Ewing Sarcoma: Current Management and Future Approaches Through Collaboration. J Clin Oncol. 2015;33(27):3036–3046. doi:10.1200/JCO.2014.59.5256

5. Ginsberg JP, Goodman P, Leisenring W, et al. Long-term Survivors of Childhood Ewing Sarcoma: Report From the Childhood Cancer Survivor Study. JNCI J Natl Cancer Inst. 2010;102(16):1272–1283. doi:10.1093/jnci/djq278

6. Delattre O, Zucman J, Plougastel B, et al. Gene fusion with an ETS DNA-binding domain caused by chromosome translocation in human tumours. Nature. 1992;359(6391):162–165. doi:10.1038/359162a0

7. Lee OW, Rodrigues C, Lin SH, et al. Targeted long-read sequencing of the Ewing sarcoma 6p25.1 susceptibility locus identifies germline-somatic interactions with EWSR1-FLI1 binding. Am J Hum Genet. 2023;110(3):427–441. doi:10.1016/j.ajhg.2023.01.017

8. Guillon N, Tirode F, Boeva V, Zynovyev A, Barillot E, Delattre O. The oncogenic EWS-FLI1 protein binds in vivo GGAA microsatellite sequences with potential transcriptional activation function. PloS One. 2009;4(3):e4932. doi:10.1371/journal.pone.0004932

9. Lawrence MS, Stojanov P, Polak P, et al. Mutational heterogeneity in cancer and the search for new cancer-associated genes. Nature. 2013;499(7457):214–218. doi:10.1038/nature12213

10. Gröbner SN, Worst BC, Weischenfeldt J, et al. The landscape of genomic alterations across childhood cancers. Nature. 2018;555(7696):321–327. doi:10.1038/nature25480

11. Brohl AS, Solomon DA, Chang W, et al. The Genomic Landscape of the Ewing Sarcoma Family of Tumors Reveals Recurrent STAG2 Mutation. PLOS Genet. 2014;10(7):e1004475. doi:10.1371/journal.pgen.1004475

12. Gillani R, Camp SY, Han S, et al. Germline predisposition to pediatric Ewing sarcoma is characterized by inherited pathogenic variants in DNA damage repair genes. Am J Hum Genet. 2022;109(6):1026–1037. doi:10.1016/j.ajhg.2022.04.007

13. Gillani R, Collins RL, Crowdis J, et al. Rare germline structural variants increase risk for pediatric solid tumors. Science. 2025;387(6729):eadq0071. doi:10.1126/science.adq0071

14. Postel-Vinay S, Véron AS, Tirode F, et al. Common variants near TARDBP and EGR2 are associated with susceptibility to Ewing sarcoma. Nat Genet. 2012;44(3):323–327. doi:10.1038/ng.1085

15. Machiela MJ, Grünewald TGP, Surdez D, et al. Genome-wide association study identifies multiple new loci associated with Ewing sarcoma susceptibility. Nat Commun. 2018;9(1):3184. doi:10.1038/s41467-018-05537-2

16. Gangwal K, Sankar S, Hollenhorst PC, et al. Microsatellites as EWS/FLI response elements in Ewing’s sarcoma. Proc Natl Acad Sci U S A. 2008;105(29):10149–10154. doi:10.1073/pnas.0801073105

17. Robison LL, Armstrong GT, Boice JD, et al. The Childhood Cancer Survivor Study: A National Cancer Institute–Supported Resource for Outcome and Intervention Research. J Clin Oncol. 2009;27(14):2308–2318. doi:10.1200/JCO.2009.22.3339

18. Troisi R, Masters MN, Joshipura K, Douglass C, Cole BF, Hoover RN. Perinatal factors, growth and development, and osteosarcoma risk. Br J Cancer. 2006;95(11):1603–1607. doi:10.1038/sj.bjc.6603474

19. Langbo C, Bach J, Kleyn M, Downes FP. From Newborn Screening to Population Health Research: Implementation of the Michigan BioTrust for Health. Public Health Rep. 2013;128(5):377. doi:10.1177/003335491312800508

20. Howell CR, Bjornard KL, Ness KK, et al. Cohort Profile: The St. Jude Lifetime Cohort Study (SJLIFE) for paediatric cancer survivors. Int J Epidemiol. 2021;50(1):39–49. doi:10.1093/ije/dyaa203

21. Chatterjee N, Hartge P, Cerhan JR, et al. Risk of non-Hodgkin’s lymphoma and family history of lymphatic, hematologic, and other cancers. Cancer Epidemiol Biomark Prev Publ Am Assoc Cancer Res Cosponsored Am Soc Prev Oncol. 2004;13(9):1415–1421.

22. Prorok PC, Andriole GL, Bresalier RS, et al. Design of the Prostate, Lung, Colorectal and Ovarian (PLCO) Cancer Screening Trial. Control Clin Trials. 2000;21(6 Suppl):273S–309S. doi:10.1016/s0197-2456(00)00098-2

23. Machiela MJ, Huang WY, Wong W, et al. GWAS Explorer: an open-source tool to explore, visualize, and access GWAS summary statistics in the PLCO Atlas. Sci Data. 2023;10(1):25. doi:10.1038/s41597-022-01921-2

24. Völzke H, Schössow J, Schmidt CO, et al. Cohort Profile Update: The Study of Health in Pomerania (SHIP). Int J Epidemiol. 2022;51(6):e372–e383. doi:10.1093/ije/dyac034

25. Hudson A, Fournier M, Coulombe J, Daee D. Using existing pediatric cancer data from the Gabriella Miller Kids First Data Resource Program. JNCI Cancer Spectr. 2023;7(6):pkad079. doi:10.1093/jncics/pkad079

26. Sequencing of 53,831 diverse genomes from the NHLBI TOPMed Program | Nature. November 22, 2024. Accessed November 22, 2024. https://www.nature.com/articles/s41586-021-03205-y

27. Auton A, Abecasis GR, Altshuler DM, et al. A global reference for human genetic variation. Nature. 2015;526(7571):68–74. doi:10.1038/nature15393

28. Das S, Forer L, Schönherr S, et al. Next-generation genotype imputation service and methods. Nat Genet. 2016;48(10):1284–1287. doi:10.1038/ng.3656

29. kf-alignment-workflow/docs/KFDRC_GATK_HAPLOTYPECALLER_CRAM_TO_GVCF_WORKFLOW_R EADME.md at master · kids-first/kf-alignment-workflow. GitHub. Accessed January 7, 2025. https://github.com/kids-first/kf-alignment-workflow/blob/master/docs/KFDRC_GATK_HAPLOTYPECALLER_CRAM_TO_GVCF_WORKFLOW_README.md

30. Patterson N, Price AL, Reich D. Population structure and eigenanalysis. PLoS Genet. 2006;2(12):e190. doi:10.1371/journal.pgen.0020190

31. Yu K, Wang Z, Li Q, et al. Population substructure and control selection in genome-wide association studies. PloS One. 2008;3(7):e2551. doi:10.1371/journal.pone.0002551

32. Brown DW, Myers TA, Machiela MJ. PCAmatchR: a flexible R package for optimal case-control matching using weighted principal components. Bioinforma Oxf Engl. 2021;37(8):1178–1181. doi:10.1093/bioinformatics/btaa784

33. Jin Y, Schaffer AA, Feolo M, Holmes JB, Kattman BL. GRAF-pop: A Fast Distance-Based Method To Infer Subject Ancestry from Multiple Genotype Datasets Without Principal Components Analysis. G3 Bethesda Md. 2019;9(8):2447–2461. doi:10.1534/g3.118.200925

34. Han B, Eskin E. Random-Effects Model Aimed at Discovering Associations in Meta-Analysis of Genome-wide Association Studies. Am J Hum Genet. 2011;88(5):586–598. doi:10.1016/j.ajhg.2011.04.014

35. Willer CJ, Li Y, Abecasis GR. METAL: fast and efficient meta-analysis of genomewide association scans. Bioinforma Oxf Engl. 2010;26(17):2190–2191. doi:10.1093/bioinformatics/btq340

36. Yang J, Ferreira T, Morris AP, et al. Conditional and joint multiple-SNP analysis of GWAS summary statistics identifies additional variants influencing complex traits. Nat Genet. 2012;44(4):369–375, S1-3. doi:10.1038/ng.2213

37. Yang J, Lee SH, Goddard ME, Visscher PM. GCTA: a tool for genome-wide complex trait analysis. Am J Hum Genet. 2011;88(1):76–82.

38. Orth MF, Surdez D, Faehling T, et al. Systematic multi-omics cell line profiling uncovers principles of Ewing sarcoma fusion oncogene-mediated gene regulation. Cell Rep. 2022;41(10). doi:10.1016/j.celrep.2022.111761

39. Sanalkumar R, Dong R, Lee L, et al. Highly connected 3D chromatin networks established by an oncogenic fusion protein shape tumor cell identity. Sci Adv. 2023;9(13):eabo3789. doi:10.1126/sciadv.abo3789

40. Grünewald TGP, Bernard V, Gilardi-Hebenstreit P, et al. Chimeric EWSR1-FLI1 regulates the Ewing sarcoma susceptibility gene EGR2 via a GGAA microsatellite. Nat Genet. 2015;47(9):1073. doi:10.1038/ng.3363

41. Johnson KM, Mahler NR, Saund RS, et al. Role for the EWS domain of EWS/FLI in binding GGAA-microsatellites required for Ewing sarcoma anchorage independent growth. Proc Natl Acad Sci. 2017;114(37):9870–9875. doi:10.1073/pnas.1701872114

42. Surdez D, Zaidi S, Grossetête S, et al. STAG2 mutations alter CTCF-anchored loop extrusion, reduce cis-regulatory interactions and EWSR1-FLI1 activity in Ewing sarcoma. Cancer Cell. 2021;39(6):810–826.e9. doi:10.1016/j.ccell.2021.04.001

43. Zhang Y, Liu T, Meyer CA, et al. Model-based Analysis of ChIP-Seq (MACS). Genome Biol. 2008;9(9):R137. doi:10.1186/gb-2008-9-9-r137

44. Showpnil IA, Selich-Anderson J, Taslim C, et al. EWS/FLI mediated reprogramming of 3D chromatin promotes an altered transcriptional state in Ewing sarcoma. Nucleic Acids Res. 2022;50(17):9814–9837. doi:10.1093/nar/gkac747

45. Beck R, Monument MJ, Watkins WS, et al. EWS/FLI-responsive GGAA-microsatellites exhibit polymorphic differences between European and African populations. Cancer Genet. 2012;205(6):304–312. doi:10.1016/j.cancergen.2012.04.004

46. Charville GW, Wang WL, Ingram DR, et al. EWSR1 fusion proteins mediate PAX7 expression in Ewing sarcoma. Mod Pathol. 2017;30(9):1312–1320. doi:10.1038/modpathol.2017.49

47. Toki S, Wakai S, Sekimizu M, et al. PAX7 immunohistochemical evaluation of Ewing sarcoma and other small round cell tumours. Histopathology. 2018;73(4):645–652. doi:10.1111/his.13689

48. Davis RJ, D’Cruz CM, Lovell MA, Biegel JA, Barr FG. Fusion of PAX7 to FKHR by the variant t(1;13)(p36;q14) translocation in alveolar rhabdomyosarcoma. Cancer Res. 1994;54(11):2869–2872.

49. Peters H, Neubüser A, Kratochwil K, Balling R. Pax9-deficient mice lack pharyngeal pouch derivatives and teeth and exhibit craniofacial and limb abnormalities. Genes Dev. 1998;12(17):2735–2747. doi:10.1101/gad.12.17.2735

50. Chen X, Li Y, Paiboonrungruang C, et al. PAX9 in Cancer Development. Int J Mol Sci. 2022;23(10):5589. doi:10.3390/ijms23105589

51. He Z, Zhang H, Xiao H, et al. Ubiquitylation of RUNX3 by RNA-binding ubiquitin ligase MEX3C promotes tumorigenesis in lung adenocarcinoma. J Transl Med. 2024;22(1):216. doi:10.1186/s12967-023-04700-8

52. Chao H, Deng L, Xu F, et al. MEX3C regulates lipid metabolism to promote bladder tumorigenesis through JNK pathway. OncoTargets Ther. 2019;12:3285–3294. doi:10.2147/OTT.S199667

53. Cao Y, Jiang F, Zhang S, Yang L, Sun Y. MEX3C promotes osteosarcoma malignant progression through negatively regulating FGF14. J BUON Off J Balk Union Oncol. 2020;25(3):1554–1561.

54. Silla T, Schmid M, Dou Y, et al. The human ZC3H3 and RBM26/27 proteins are critical for PAXT-mediated nuclear RNA decay. Nucleic Acids Res. 2020;48(5):2518–2530. doi:10.1093/nar/gkz1238

55. Zhu C, Liu C, Chai Z. Role of the PADI family in inflammatory autoimmune diseases and cancers: A systematic review. Front Immunol. 2023;14:1115794. doi:10.3389/fimmu.2023.1115794

56. Leacock SW, Basse AN, Chandler GL, Kirk AM, Rakheja D, Amatruda JF. A zebrafish transgenic model of Ewing’s sarcoma reveals conserved mediators of EWS-FLI1 tumorigenesis. Dis Model Mech. 2012;5(1):95–106. doi:10.1242/dmm.007401

57. VanHeyst KA, Choi SH, Kingsley DT, Huang AY. Ectopic Tumor VCAM-1 Expression in Cancer Metastasis and Therapy Resistance. Cells. 2022;11(23):3922. doi:10.3390/cells11233922

58. Kingsley DT, Choi SH, Myers J, Huang A. Assessing the Role of the VCAM1-VLA4 Interaction in Macrophage Polarization in Pulmonary Metastatic Osteosarcoma. J Immunol. 2023;210(1_Supplement):169.07. doi:10.4049/jimmunol.210.Supp.169.07

59. Kim M, Kim JH, Jang HR, et al. LRRC3B, encoding a leucine-rich repeat-containing protein, is a putative tumor suppressor gene in gastric cancer. Cancer Res. 2008;68(17):7147–7155. doi:10.1158/0008-5472.CAN-08-0667

60. Luo L, Fu S, Du W, et al. LRRC3B and its promoter hypomethylation status predicts response to anti-PD-1 based immunotherapy. Front Immunol. 2023;14:959868. doi:10.3389/fimmu.2023.959868

