## Supplemental Tables for "Genome-wide association study meta-analysis identifies susceptibility loci informing Ewing sarcoma etiology and potential mechanisms of risk"

**Supplemental Table 1**. Case Characteristics by metastatic disease and overall for EwS cases with available clinical data*.

| Trait | N (%) or mean (SD) |
|---|---|
| **Age at diagnosis** | 14.5 (7.6) |
| **Sex** | |
| Male | 709 (55.9) |
| Female | 559 (44.1) |
| **Metastasis at Diagnosis** | |
| Yes | 268 (28.7) |
| No | 666 (71.3) |
| **Tumor Location** | |
| Head/Neck | 71 (7.9) |
| Chest | 152 (16.9) |
| Spine | 125 (13.9) |
| Upper Extremity | 83 (9.2) |
| Abdomen | 11 (1.2) |
| Pelvis | 207 (23.0) |
| Lower Extremity | 250 (27.8) |

*Clinical data was not available for GMKF, St. Jude, CESS cases and was not available across all cases from the original GWAS.

**Supplemental Table 2**. Contributing Studies and case/control totals for each study and each meta-analysis set.

| Analysis Set | Contributing Studies/Institutions for Cases[a] | Genotyping Array | Fixed Effects Meta-analysis[b] | | | METAL Meta-analysis[c] | | | |
|---|---|---|---|---|---|---|---|---|---|
| | | | Cases | Controls[a] | Total | Cases | Controls[a] | Total | ESS[d] |
| IC-GWAS1 | Institut Curie (IC) samples from published GWAS 1 | Illumina 610 Quadv1 array | 401 | 682 | 1,083 | 401 | 682 | 1,083 | 1,010 |
| Omni Set | Centers for Cancer Research (CCR), Bone Disease Study, Institut Curie (IC) | Illumina OmniExpress-24 v1.1 array | 170 | 340 | 510 | 170 | 340 | 510 | 453 |
| CCSS | Childhood Cancer Survivor Study (CCSS) | Human Omni5Exome array | 159 | 319 | 478 | 159 | 319 | 478 | 424 |
| GSA Set | Children's Oncology Group (COG), Institut Curie (IC), Michigan Blood Spots | Infinium Global Screening Array-24 v2-0 | 551 | 5,510 | 6,061 | 551 | 5,510 | 6,061 | 2,004 |
| CESS | Cooperative Ewing Sarcoma Study (CESS) | Infinium Global Screening Array-24 v1-3 | 335 | 2,816 | 3,151 | 335 | 2,816 | 3,151 | 1,198 |
| SJLIFE | St. Jude Lifetime Cohort Study at St. Jude Children's Research Hospital | Whole genome sequencing | 111 | 285 | 396 | 111 | 285 | 396 | 320 |
| GMKF | Gabriela Miller Kids Foundation (GMKF) Parent-Offspring study | Whole genome sequencing | - | - | - | 287 | 573 | 870 | 765 |
| Totals[c] | | | 1,727 | 9,952 | 11,679 | 2,014 | 10,525 | 12,539 | |

[a]Controls were ancestry-matched and sourced from cancer-free individuals from the Prostate, Lung, Colorectal and Ovarian Screening Trial (PLCO), American Cancer Society Prevention Study II, the Spanish Bladder Cancer Study, and Study of Health In Pomerania - TREND (SHIP-TREND) study. Gabriela Miller Kids First (GMKF) controls were parents of affected EwS patients.

[b]Primary Fixed Effects Meta-analysis was retricted to 80% genetic similarity to European samples and case-control study sets.

[c]Meta-analysis in METAL was used to combine *P* values across studies taking sample size and direction of effect into account while combining family-based TDT results with case-control results.

[d] Effective sample size (ESS) used in sample weighted meta-analysis using METAL. ESS=4xcase N X control N/(case N + control N).

**Supplemental Table 3**. Allele frequency of the effect (risk) allele for genome wide significant lead variants in different populations.

| Chr | Locus[a] | rsID[b] | Referent Allele | Effect Allele | Odds Ratio | $EAF^{Global}$ | $EAF^{EUR}$ | $EAF^{AFR}$ | $EAF^{ASN}$ | $EAF^{EAS}$ | $EAF^{SAS}$ | $EAF^{LAT1}$ | $EAF^{LAT2}$ |
|---|---|---|---|---|---|---|---|---|---|---|---|---|---|
| 1 | 1p36.22 | rs11589058 | A | T | 2.11 | 0.7574 | 0.7971 | 0.5346 | 0.9460 | **0.9500** | 0.8800 | 0.7050 | 0.8770 |
| | 1p36.13 | rs545301 | C | T | 2.49 | 0.0728 | 0.0100 | 0.2651 | **0.3420** | 0.3330 | 0.1540 | 0.0960 | 0.2740 |
| 5 | 5q32.3 | rs1488534366* | C | CT | 1.36 | 0.8893 | 0.8580 | 0.9981 | 0.9900 | **1.0000** | **1.0000** | **1.0000** | **1.0000** |
| | 6p25.1 | rs17142617 | A | G | 1.53 | 0.1351 | 0.1237 | 0.2651 | **0.1890** | 0.1840 | 0.0790 | 0.1780 | 0.0769 |
| 6 | 6p22.1 | rs6935895 | T | A | 2.95 | 0.0534 | 0.0100 | **0.2834** | 0.0270 | 0.0300 | 0.0000 | 0.0890 | 0.0260 |
| 7 | 7q32.3 | rs350653 | T | C | 1.95 | 0.8351 | 0.9003 | 0.4956 | 0.9820 | **0.9900** | 0.9100 | 0.8560 | 0.9390 |
| 8 | 8q24.21 | rs10108830 | T | A | 1.34 | 0.3212 | 0.3296 | 0.2168 | 0.4720 | 0.4900 | 0.2400 | 0.4250 | 0.5380 |
| 10 | 10q21.2 | rs10822056 | C | T | 1.63 | 0.5136 | 0.5183 | **0.5933** | 0.3523 | 0.2935 | 0.5460 | 0.5292 | 0.3370 |
| 11 | 11q24.1 | rs10790459 | T | G | 1.30 | 0.7155 | 0.7226 | 0.4357 | 0.8325 | 0.8168 | 0.7370 | 0.6270 | 0.7377 |
| 12 | 12q14.3 | rs150006321 | C | T | 1.31 | 0.7942 | 0.7518 | **0.9538** | 0.8330 | 0.8600 | 0.6800 | 0.7880 | 0.5670 |
| 14 | 14q13.3 | rs2764962 | T | C | 1.26 | 0.5887 | 0.5675 | **0.8154** | 0.9060 | 0.9060 | 0.6138 | 0.6680 | 0.7617 |
| 15 | 15q15.1 | rs9919974 | T | G | 1.67 | 0.2507 | **0.2831** | 0.1466 | 0.0180 | 0.0100 | 0.2000 | 0.1780 | 0.1100 |
| 18 | 18q21.2 | rs1563410 | C | T | 1.25 | 0.4700 | 0.4813 | 0.4019 | **0.7560** | 0.7160 | 0.5820 | 0.4720 | 0.4582 |
| 20 | 20p11.22 | rs6047482 | T | A | 1.71 | 0.6683 | **0.7288** | 0.3487 | 0.6880 | 0.6900 | 0.6800 | 0.6510 | 0.6460 |

EAF = Effect Allele Frequency , EUR=European, AFR=African, ASN=Asian, EAS=East Asian, SAS=South Asian, LAT1=Latin American 1, LAT2=Latin American 2.

EAF is based on the ALFA project which provides aggregate allele frequency from dbGaP.

*EAF based on lead variant from METAL as lead variant in case-control meta-analysis (indel chr5:146327889) was not listed in dbSNP.

**Supplemental Table 4**. Study-specific summary statistics for EwS meta-analysis. Variants were not included if minor allele frequency was < 1%. Study estimates were calculated in Plink for all studies except CESS which used SNPTEST.

| | | | | Institut Curie Set 1 - 1st Published GWAS (N=1,083) | | | | | |
|---|---|---|---|---|---|---|---|---|---|
| Chr | Position[a] | rsID[b] | Risk allele | OR | SE | Lower 95% CI | Upper 95% CI | *P* value | INFO[c] |
| 1 | 10,987,244 | rs11589058 | T | 2.46 | 0.146 | 1.85 | 3.27 | 5.94E-10 | 0.95 |
| 1 | 17,873,309 | **rs545301** | T | 1.86 | 0.471 | 0.74 | 4.68 | 0.1875 | 0.77 |
| 5 | 146,327,889 | **chr5:146327889** | CT | 1.35 | 0.116 | 1.07 | 1.69 | 0.0103 | 0.88 |
| 6 | 6,837,792 | rs17142617 | G | 1.76 | 0.126 | 1.38 | 2.25 | 6.85E-06 | 0.88 |
| 6 | 29,543,984 | **rs6935895** | A | 2.95 | 0.470 | 1.18 | 7.36 | 0.0200 | 0.97 |
| 7 | 131,030,999 | **rs350653** | C | 1.77 | 0.206 | 1.18 | 2.64 | 0.0058 | 0.84 |
| 8 | 129,448,673 | **rs10108830** | A | 1.32 | 0.092 | 1.10 | 1.58 | 0.0027 | 0.98 |
| 10 | 62,754,351 | rs10822056 | T | 1.93 | 0.100 | 1.59 | 2.35 | 4.69E-11 | 0.99 |
| 11 | 121,767,808 | rs10790456 | G | 1.21 | 0.102 | 0.99 | 1.48 | 0.0614 | 0.80 |
| 12 | 66,069,074 | **rs150006321** | T | 1.33 | 0.110 | 1.07 | 1.65 | 0.0092 | 0.95 |
| 14 | 36,759,015 | **rs2764962** | C | 1.19 | 0.094 | 0.99 | 1.44 | 0.0600 | 0.92 |
| 15 | 40,045,261 | rs9919974 | G | 1.61 | 0.098 | 1.33 | 1.95 | 1.09E-06 | 0.98 |
| 18 | 51,335,382 | **rs1563410** | T | 1.19 | 0.090 | 1.00 | 1.42 | 0.0549 | 0.98 |
| 20 | 21,559,045 | rs6047482 | A | 1.81 | 0.117 | 1.44 | 2.27 | 3.78E-07 | 0.95 |

| | Omni: Bone Disease Study, Institut Curie Set # 2, Centers for Cancer Research (N=510) | | | | | | Childhood Cancer Survivor Study (N=478) | | | | | |
|---|---|---|---|---|---|---|---|---|---|---|---|---|
| rsID[b] | OR | SE | Lower 95% CI | Upper 95% CI | *P* value | INFO[c] | OR | SE | Lower 95% CI | Upper 95% CI | *P* value | INFO[c] |
| rs11589058 | 2.05 | 0.184 | 1.43 | 2.94 | 9.49E-05 | 0.82 | 2.00 | 0.212 | 1.32 | 3.02 | 0.0011 | 0.99 |
| **rs545301** | NA | NA | NA | NA | NA | 0.77 | 1.71 | 0.693 | 0.44 | 6.64 | 0.4393 | 1.00 |
| **chr5:146327889** | 1.84 | 0.188 | 1.27 | 2.66 | 0.0012 | 0.91 | 1.64 | 0.183 | 1.15 | 2.35 | 0.0068 | 0.97 |
| rs17142617 | 1.22 | 0.195 | 0.83 | 1.79 | 0.3129 | 0.97 | 1.97 | 0.195 | 1.35 | 2.89 | 0.0005 | 1.00 |
| **rs6935895** | NA | NA | NA | NA | NA | 1.00 | 1.06 | 0.654 | 0.29 | 3.82 | 0.9283 | 1.00 |
| **rs350653** | 2.04 | 0.293 | 1.15 | 3.62 | 0.0152 | 0.85 | 1.08 | 0.268 | 0.64 | 1.83 | 0.7773 | 0.98 |
| **rs10108830** | 1.26 | 0.142 | 0.95 | 1.66 | 0.1070 | 0.98 | 1.33 | 0.141 | 1.01 | 1.75 | 0.0414 | 1.00 |
| rs10822056 | 1.66 | 0.148 | 1.24 | 2.22 | 0.0007 | 0.97 | 1.72 | 0.147 | 1.29 | 2.29 | 0.0002 | 1.00 |
| rs10790456 | 1.33 | 0.152 | 0.99 | 1.80 | 0.0593 | 1.00 | 1.43 | 0.173 | 1.02 | 2.00 | 0.0389 | 1.00 |
| **rs150006321** | 1.24 | 0.168 | 0.89 | 1.73 | 0.1993 | 0.94 | 1.42 | 0.171 | 1.02 | 1.99 | 0.0390 | 1.00 |
| **rs2764962** | 1.21 | 0.140 | 0.92 | 1.59 | 0.1804 | 1.00 | 1.30 | 0.142 | 0.98 | 1.71 | 0.0664 | 1.00 |
| rs9919974 | 1.69 | 0.144 | 1.28 | 2.24 | 0.0003 | 0.92 | 1.64 | 0.146 | 1.23 | 2.18 | 0.0007 | 0.99 |
| **rs1563410** | 1.39 | 0.139 | 1.06 | 1.82 | 0.0185 | 0.99 | 1.26 | 0.147 | 0.95 | 1.68 | 0.1121 | 1.00 |
| rs6047482 | 1.63 | 0.171 | 1.17 | 2.28 | 0.0042 | 0.96 | 1.86 | 0.181 | 1.31 | 2.65 | 0.0006 | 1.00 |

| | GSA: Children's Oncology Group, Michigan Blood Spots, Institut Curie Set # 3 (N=6,061) | | | | | | CESS (N=3,151) | | | | | |
|---|---|---|---|---|---|---|---|---|---|---|---|---|
| rsID[b] | OR | SE | Lower 95% CI | Upper 95% CI | *P* value | INFO[c] | OR | SE | Lower 95% CI | Upper 95% CI | *P* value | INFO[c] |
| rs11589058 | 1.95 | 0.098 | 1.61 | 2.37 | 7.93E-12 | 0.98 | 1.94 | 0.154 | 1.43 | 2.63 | 1.51E-06 | 0.96 |
| **rs545301** | 2.56 | 0.176 | 1.81 | 3.61 | 1.03E-07 | 0.94 | 3.67 | 0.559 | 1.23 | 10.98 | 0.7915 | 0.99 |
| **chr5:146327889** | 1.29 | 0.082 | 1.10 | 1.52 | 1.70E-03 | 0.91 | 1.09 | 0.208 | 0.95 | 1.25 | 0.7161 | 0.92 |
| rs17142617 | 1.40 | 0.085 | 1.18 | 1.65 | 8.34E-05 | 1.00 | 1.53 | 0.139 | 1.16 | 2.00 | 0.0825 | 0.93 |
| **rs6935895** | 3.13 | 0.158 | 2.30 | 4.26 | 4.46E-13 | 0.98 | NA | NA | NA | NA | NA | NA |
| **rs350653** | 2.25 | 0.150 | 1.67 | 3.01 | 4.49E-08 | 0.98 | 2.32 | 0.229 | 1.48 | 3.63 | 1.29E-05 | 0.96 |
| **rs10108830** | 1.31 | 0.065 | 1.15 | 1.48 | 4.36E-05 | 0.99 | 1.50 | 0.104 | 1.22 | 1.84 | 0.0034 | 0.98 |
| rs10822056 | 1.55 | 0.066 | 1.36 | 1.77 | 3.28E-11 | 0.99 | 1.51 | 0.105 | 1.23 | 1.85 | 1.18E-05 | 0.96 |
| rs10790456 | 1.28 | 0.074 | 1.10 | 1.48 | 9.71E-04 | 0.94 | 1.22 | 0.096 | 1.01 | 1.48 | 0.0673 | 0.92 |
| **rs150006321** | 1.31 | 0.077 | 1.13 | 1.53 | 4.04E-04 | 0.99 | 1.25 | 0.094 | 1.04 | 1.50 | 0.6678 | 0.96 |
| **rs2764962** | 1.34 | 0.067 | 1.18 | 1.53 | 1.07E-05 | 1.00 | 1.23 | 0.102 | 1.01 | 1.50 | 0.0321 | 1.00 |
| rs9919974 | 1.70 | 0.066 | 1.49 | 1.93 | 9.88E-16 | 0.99 | 1.67 | 0.106 | 1.35 | 2.05 | 3.66E-08 | 0.99 |
| **rs1563410** | 1.31 | 0.06 | 1.16 | 1.48 | 2.10E-05 | 0.99 | 1.18 | 0.100 | 0.97 | 1.44 | 0.7050 | 0.96 |
| rs6047482 | 1.72 | 0.083 | 1.46 | 2.02 | 6.91E-11 | 0.99 | 1.74 | 0.132 | 1.34 | 2.25 | 2.97E-05 | 0.95 |

| | St. Jude Life Study (N=396) | | | | | Gabriela Miller Kids First EwS Trios[d] (N=860) | | | | | | |
|---|---|---|---|---|---|---|---|---|---|---|---|---|
| rsID[b] | OR | SE | Lower 95% CI | Upper 95% CI | *P* value | T | U | TDT OR | CHISQ | Lower 95% CI | Upper 95% CI | *P* value |
| rs11589058 | 3.45 | 0.2793 | 2.00 | 5.97 | ###### | 48 | 25 | 1.92 | 7.25 | 1.19 | 3.09 | 0.0071 |
| **rs545301** | NA | NA | NA | NA | NA | 9 | 1 | 9.00 | 6.40 | 1.64 | 49.38 | 0.0114 |
| **chr5:146327889** | NA | NA | NA | NA | NA | 69 | 68 | 1.02 | 0.01 | 0.72 | 1.43 | 0.9319 |
| rs17142617 | 1.82 | 0.2345 | 1.15 | 2.88 | 0.0107 | 40 | 15 | 2.67 | 11.36 | 1.51 | 4.72 | 7.49E-04 |
| **rs6935895** | NA | NA | NA | NA | NA | 1 | 0 | N/A | 1.00 | N/A | N/A | 0.3173 |
| **rs350653** | NA | NA | NA | NA | NA | 16 | 8 | 2.00 | 2.67 | 0.87 | 4.59 | 0.1025 |
| **rs10108830** | NA | NA | NA | NA | NA | 123 | 89 | 1.38 | 5.45 | 1.05 | 1.81 | 0.0195 |
| rs10822056 | 1.60 | 0.177 | 1.13 | 2.27 | 0.0078 | 167 | 70 | 2.39 | 39.70 | 1.82 | 3.13 | 2.96E-10 |
| rs10790456 | 1.82 | 0.204 | 0.87 | 1.93 | 0.0033 | 114 | 71 | 1.61 | 10.00 | 1.19 | 2.16 | 0.0016 |
| **rs150006321** | NA | NA | NA | NA | NA | 34 | 24 | 1.42 | 1.72 | 0.84 | 2.39 | 0.1892 |
| **rs2764962** | 1.12 | 0.159 | 0.82 | 1.53 | 0.4799 | 77 | 78 | 0.99 | 0.01 | 0.72 | 1.35 | 0.936 |
| rs9919974 | NA | NA | NA | NA | NA | 111 | 49 | 2.65 | 24.02 | 1.79 | 3.92 | 9.51E-07 |
| **rs1563410** | 1.09 | 0.163 | 0.79 | 1.50 | 0.6043 | 135 | 93 | 1.45 | 7.74 | 1.12 | 1.88 | 0.0054 |
| rs6047482 | 1.35 | 0.196 | 0.92 | 1.98 | 0.1302 | 37 | 35 | 1.05 | 0.06 | 0.70 | 1.57 | 0.8137 |

[a] SNP position according to NCBI Human Genome Build 38.

[b] Variants in bold face were newly discovered.

[c] Imputation information score. No Info score listed for SJLIFE or GMKF because data was whole genome sequencing, not imputation.

[d] OR and test statistics are based on Transmission Disequilibrium Test (TDT). T and U represent numbers of transmitted and untransmitted alleles respectively.

**Supplemental Table 5**. Leave one out (LOO) fixed effects (FE) meta-analysis odds ratio (OR) and 95% confidence intervals (CI).  FE meta-analysis was conducted after leaving out each study one at a time.

| | | Removing GSA | | | | Removing IC-GWAS1 | | | | Removing Omni Set | | | | Removing CCSS Set | | | | Removing CESS set | | | | Removing SJLIFE set | | | |
|---|---|---|---|---|---|---|---|---|---|---|---|---|---|---|---|---|---|---|---|---|---|---|---|---|---|
| **Chr** | **Variant[a]** | **OR** | **Lower 95% CI** | **Upper 95% CI** | ***P* value** | **OR** | **Lower 95% CI** | **Upper 95% CI** | ***P* value** | **OR** | **Lower 95% CI** | **Upper 95% CI** | ***P* value** | **OR** | **Lower 95% CI** | **Upper 95% CI** | ***P* value** | **OR** | **Lower 95% CI** | **Upper 95% CI** | ***P* value** | **OR** | **Lower 95% CI** | **Upper 95% CI** | ***P* value** |
| 1 | rs11589058 | 2.22 | 1.65 | 3.43 | 3.78E-23 | 2.04 | 1.60 | 2.81 | 4.51E-25 | 2.12 | 1.66 | 2.92 | 7.82E-30 | 2.12 | 1.67 | 2.91 | 7.72E-31 | 2.15 | 1.67 | 3.00 | 3.31E-29 | 2.06 | 1.64 | 2.77 | 1.42E-29 |
| | **rs545301** | 2.28 | 1.22 | 4.27 | 9.80E-03 | 2.58 | 1.87 | 3.55 | 6.84E-09 | 2.49 | 1.84 | 3.37 | 3.49E-09 | 2.54 | 1.86 | 3.46 | 4.05E-09 | 2.41 | 1.76 | 3.30 | 4.35E-08 | 2.49 | 1.84 | 3.37 | 3.49E-09 |
| 5 | **chr5:146327889** | 1.44 | 1.23 | 1.68 | 6.82E-06 | 1.37 | 1.20 | 1.56 | 2.60E-10 | 1.32 | 1.18 | 1.49 | 3.38E-06 | 1.34 | 1.19 | 1.50 | 1.54E-06 | 1.39 | 1.24 | 1.56 | 3.57E-08 | 1.36 | 1.22 | 1.53 | 6.25E-08 |
| 6 | rs17142617 | 1.64 | 1.42 | 1.89 | 1.79E-11 | 1.48 | 1.31 | 1.67 | 2.60E-10 | 1.56 | 1.39 | 1.75 | 1.51E-14 | 1.50 | 1.33 | 1.67 | 3.64E-12 | 1.53 | 1.36 | 1.72 | 2.12E-12 | 1.51 | 1.35 | 1.69 | 3.89E-13 |
| | **rs6935895** | 2.09 | 0.99 | 4.40 | 5.22E-02 | 1.40 | 1.24 | 1.58 | 1.63E-12 | 2.95 | 2.18 | 3.98 | 1.06E-13 | 3.11 | 2.32 | 4.17 | 2.87E-14 | 2.95 | 2.22 | 3.92 | 1.06E-13 | 2.95 | 2.22 | 3.92 | 1.06E-13 |
| 7 | **rs350653** | 1.77 | 1.39 | 2.24 | 2.66E-06 | 2.00 | 1.62 | 2.45 | 5.33E-11 | 1.94 | 1.59 | 2.35 | 2.53E-11 | 2.11 | 1.74 | 2.57 | 7.98E-14 | 1.88 | 1.54 | 2.30 | 8.72E-10 | 1.95 | 1.62 | 2.34 | 1.27E-12 |
| 8 | **rs10108830** | 1.36 | 1.22 | 1.52 | 5.50E-08 | 1.34 | 1.22 | 1.48 | 1.16E-09 | 1.35 | 1.23 | 1.47 | 3.94E-11 | 1.34 | 1.22 | 1.46 | 9.78E-11 | 1.31 | 1.19 | 1.43 | 1.25E-08 | 1.34 | 1.23 | 1.45 | 1.17E-11 |
| 10 | rs10822056 | 1.70 | 1.52 | 1.89 | 8.10E-21 | 1.57 | 1.43 | 1.73 | 1.69E-21 | 1.63 | 1.49 | 1.78 | 1.08E-27 | 1.63 | 1.49 | 1.78 | 2.75E-27 | 1.66 | 1.51 | 1.82 | 4.38E-27 | 1.64 | 1.50 | 1.78 | 1.10E-28 |
| 11 | **rs10790456** | 1.31 | 1.16 | 1.48 | 1.43E-05 | 1.32 | 1.19 | 1.47 | 2.32E-07 | 1.29 | 1.17 | 1.42 | 3.17E-07 | 1.29 | 1.17 | 1.42 | 3.90E-07 | 1.31 | 1.18 | 1.45 | 1.79E-07 | 1.27 | 1.15 | 1.40 | 9.76E-07 |
| 12 | **rs150006321** | 1.30 | 1.14 | 1.48 | 6.36E-05 | 1.30 | 1.16 | 1.45 | 3.11E-06 | 1.31 | 1.19 | 1.46 | 2.09E-07 | 1.30 | 1.17 | 1.44 | 7.39E-07 | 1.32 | 1.18 | 1.47 | 6.17E-07 | 1.31 | 1.18 | 1.44 | 9.39E-08 |
| 14 | **rs2764962** | 1.21 | 1.09 | 1.34 | 3.51E-04 | 1.28 | 1.17 | 1.40 | 1.51E-07 | 1.27 | 1.16 | 1.38 | 7.14E-08 | 1.26 | 1.15 | 1.37 | 1.67E-07 | 1.27 | 1.16 | 1.39 | 2.40E-07 | 1.27 | 1.17 | 1.38 | 2.83E-08 |
| 15 | rs9919974 | 1.65 | 1.47 | 1.85 | 2.63E-17 | 1.68 | 1.53 | 1.85 | 3.06E-26 | 1.67 | 1.52 | 1.82 | 1.73E-28 | 1.67 | 1.53 | 1.83 | 6.90E-29 | 1.67 | 1.52 | 1.83 | 2.34E-26 | 1.67 | 1.53 | 1.82 | 2.09E-31 |
| 18 | **rs1563410** | 1.21 | 1.09 | 1.34 | 2.86E-04 | 1.27 | 1.16 | 1.38 | 1.93E-07 | 1.24 | 1.14 | 1.35 | 4.56E-07 | 1.25 | 1.15 | 1.36 | 1.30E-07 | 1.26 | 1.16 | 1.38 | 1.17E-07 | 1.26 | 1.16 | 1.37 | 2.72E-08 |
| 20 | rs6047482 | 1.71 | 1.50 | 1.95 | 1.60E-15 | 1.69 | 1.51 | 1.89 | 2.88E-19 | 1.72 | 1.54 | 1.92 | 4.34E-23 | 1.70 | 1.53 | 1.89 | 2.50E-22 | 1.71 | 1.53 | 1.91 | 4.74E-21 | 1.74 | 1.57 | 1.94 | 1.01E-24 |

[a]Variants in bold face were newly discovered.

**Supplemental Table 6**. Case-trio analysis lead variant in TDT results at each significant locus from meta-analysis results. Case-parent trios were from Gabriela Miller Kid's Foundation (GMKF) set of 287 affected cases and 573 parents.

| Chr | Locus[a] | rsID[b] | Position[c] | Referent Allele | Risk Allele | U | T | OR[d] | CHISQ | Association *P* value | Distance in kb |
|---|---|---|---|---|---|---|---|---|---|---|---|
| 1 | 1p36.22 | **rs879806904** | 10943077 | G | T | 11 | 65 | 5.91 | 38.37 | 5.86E-10 | 44.17 |
| | 1p36.13 | rs594534 | 17897619 | C | T | 5 | 40 | 8.00 | 27.22 | 1.81E-07 | 24.31 |
| 5 | 5q32.3 | **rs1488534366** | 145999736 | A | C | 9 | 73 | 8.11 | 49.95 | 1.58E-12 | 328.15 |
| 6 | 6p25.1 | rs11243125 | 6869665 | G | T | 52 | 105 | 2.02 | 17.89 | 2.34E-05 | 31.87 |
| | 6p22.1 | rs6935895 | 29543124 | C | T | 71 | 140 | 1.97 | 22.56 | 2.03E-06 | 0.86 |
| 7 | 7q32.3 | **rs971539419** | 131109624 | C | A | 5 | 51 | 10.19 | 37.79 | 7.90E-10 | 78.63 |
| 8 | 8q24.21 | rs1172924357 | 129364956 | T | G | 0 | 29 | - | 29 | 7.24E-08 | 83.72 |
| 10 | 10q21.2 | **rs937223645** | 62878036 | C | A | 14 | 89 | 6.36 | 54.61 | 1.47E-13 | 123.67 |
| 11 | 11q24.1 | **rs1307551815** | 121785466 | T | A | 4 | 50 | 12.50 | 39.19 | 3.86E-10 | 17.66 |
| 12 | 12q14.3 | **rs376370554** | 66069186 | A | G | 3 | 46 | 15.33 | 37.73 | 8.11E-10 | 0.11 |
| 14 | 14q13.3 | rs1749944 | 36757328 | C | T | 68 | 33 | 2.06 | 12.13 | 4.97E-04 | 1.69 |
| 15 | 15q15.1 | **rs147622877** | 40077773 | T | G | 7 | 56 | 8.00 | 38.11 | 6.68E-10 | 32.51 |
| 18 | 18q21.2 | **rs796151360** | 51039505 | C | A | 44 | 132 | 3.00 | 44 | 3.28E-11 | 295.88 |
| 20 | 20p11.22 | **rs1202095641** | 21444365 | G | T | 7 | 63 | 9.00 | 44.8 | 2.18E-11 | 114.68 |

[a]Cytogenetic regions according to NCBI Human Genome Build 38.

[b]Variants in bold face were signficant at genome-wide level in TDT analysis.

[c]Variant position according to NCBI Human Genome Build 38.

[d]Odds Ratio (OR) is derived from transmission disequilibrium test (TDT) and effectively the ratio between Transmitted (T) and Untransmitted (U) alleles.

**Supplemental Table 7**. Conditional significant variants in genome-wide significant loci for subjects >80% genetic similarity to European ancestry for Ewing sarcoma susceptibility. GWAS was conducted at a 1% minor allele threshold.

| Chr | Locus[a] | rsID[b] | Position[c] | Reference Allele | Risk Allele | EAF | Marginal effect (Beta) | Marginal SE | Marginal *P* value | Index rsID[d] | Conditional effect (Beta) | Conditional SE | Conditional *P* value | Distance in Kb | LD (r) in Eur |
|---|---|---|---|---|---|---|---|---|---|---|---|---|---|---|---|
| 6 | 6p25.1 | rs367699968 | 6845462 | T | TAC | 0.00021283 | -0.443363 | 0.0724522 | 9.39E-10 | rs17142617 | -0.4437 | 0.0724525 | 9.10E-10 | 7.67 | 0.011318 |
| 7 | 7q32.3 | rs6978148 | 130993395 | C | T | 0.337262 | -0.265796 | 0.0448301 | 3.05E-09 | rs350653 | -0.2954 | 0.0450676 | 5.55E-11 | 37.60 | -0.11276 |
| 15 | 15q15.1 | rs41500744 | 40086829 | C | A | 0.186815 | -0.692191 | 0.0649463 | 1.60E-26 | rs9919974 | -0.5578 | 0.0668998 | 7.53E-17 | 41.57 | -0.26904 |
| 20 | 20p11.23 | rs6035886 | 21555002 | C | T | 0.0977706 | 0.472024 | 0.0591816 | 1.51E-15 | rs6047482 | 0.3908 | 0.059955 | 7.12E-11 | 4.04 | -0.19577 |

[a]Cytogenetic regions according to NCBI Human Genome Build 38.

[b]Variants identified as independent .

[c]Variant position according to NCBI Human Genome Build 38.

[d]Lead variant in GWAS conditioned upon in the model.

**Supplemental Table 8**. Contributing samples for case-only GWAS by sex, metastasis, and age at diagnosis. Case-only analysis was restricted to individuals with > 80% genetic similarlity to European reference populations.

| Analysis Set | Contributing Studies/Institutions for Cases* | Sex N(%) | | Metastasis N (%) | | Age at Diagnosis N (%) | | Age at Diagnosis N (%) | | Age at Diagnosis N (%) | | Total* |
|---|---|---|---|---|---|---|---|---|---|---|---|---|
| | | **Males** | **Females** | **Yes** | **No** | **<10** | **10+** | **<16** | **16+** | **<20** | **20+** | |
| GSA Set | Children's Oncology Group (COG), Institut Curie, Michigan Blood Spots | 305 (55.4) | 246 (44.6) | 112 (25.3) | 331 (74.7) | 126 (25.1) | 376 (74.1) | 267 (60.4) | 175 (39.6) | 392 (88.7) | 50 (11.3) | 551 |
| IC-GWAS1 | Institut Curie samples from published GWAS 1 | 247 (61.9) | 152 (38.1) | 129 (34.7) | 242 (65.3) | 77 (20.1) | 307 (79.9) | 223 (58.1) | 161 (41.9) | 297 (77.3) | 87 (22.7) | 401 |
| Omni Set | Centers for Cancer Research, Bone Disease Study, Institut Curie | 84 (52.8) | 75 (47.2) | 27 (22.5) | 93 (77.5) | 20 (16.8) | 99 (83.2) | 65 (54.6) | 54 (45.4) | 87 (73.1) | 32 (26.9) | 170 |
| CCSS | Childhood Cancer Survivor Study | 73 (45.9) | 86 (54.1) | NA | NA | 49 (30.8) | 110 (69.2) | 116 (73.0) | 43 (27.0) | 151 (95.0) | 8 (5.0) | 159 |
| **Total** | | **709 (55.9)** | **559 (44.1)** | **268 (28.7)** | **666 (71.3)** | **272 (23.4)** | **892 (76.6)** | **671 (60.8)** | **433 (39.2)** | **927 (84.0)** | **177 (16.0)** | **1281** |

*Where phenotype data was unavailable, not all available cases were included and may not sum to total.

**Supplemental Table 9**. Comparing statistical enrichment to consecutive GGAA repeats or FLI1 binding.  10,000 variants from the GWAS meta-analysis were selected with replacement on each chromosome, distance between those variants and GGAA repeats or FLI1 binding was selected to make a distribution to compare against lead variants.

| | | | **GGAA Repeats** | | | | | | **FLI1 binding** | | | | | |
|---|---|---|---|---|---|---|---|---|---|---|---|---|---|---|
| **Chromosome** | **rsID** | **Position[a]** | **4 or more GGAA Repeats** | | | **13 or more GGAA repeats** | | | **30 or more** | | | **45 or more** | | |
| | | | **Position nearest** | **Distance to** | **Percentile[b]** | **Position nearest** | **Distance to** | **Percentile[b]** | **Position nearest** | **Distance to nearest** | **Percentile[b]** | **Position nearest** | **Distance to nearest** | **Percentile[b]** |
| 1 | rs11589058 | 10,987,244 | 10,987,423 | 179 | **0.0017** | 10,992,133 | 4,889 | **0.0065** | 10,987,929 | 685 | **0.0045** | 10,987,951 | 707 | **0.0026** |
| | rs545301 | 17,873,309 | 17,897,593 | 24,284 | 0.1953 | 17,716,773 | 180,820 | 0.1926 | 18,359,405 | 486,096 | 0.7937 | 18,359,469 | 486,160 | 0.6473 |
| 5 | chr5:146327889:C:CT | 146,327,889 | 146,364,395 | 36,506 | 0.2203 | 146,002,535 | 325,354 | 0.2250 | 146,364,182 | 36,293 | 0.1471 | 146,364,258 | 36,369 | 0.1024 |
| 6 | rs17142617 | 6,837,792 | 6,837,048 | 744 | **0.0084** | 6,023,216 | 814,576 | 0.2572 | 6,837,124 | 668 | **0.0050** | 6,837,029 | 763 | **0.0028** |
| | rs6935895 | 29,543,984 | 29,642,448 | 98,464 | 0.5585 | 28,707,629 | 836,355 | 0.5034 | 29,964,995 | 421,011 | 0.8020 | 30,682,721 | 1,138,737 | 0.9186 |
| 7 | rs350653 | 131,030,999 | 131,035,309 | 4,310 | **0.0422** | 131,035,408 | 4,409 | **0.0056** | 131,035,084 | 4,085 | **0.0209** | 131,035,130 | 4,131 | **0.0129** |
| 8 | rs10108830 | 129,448,673 | 129,449,337 | 704 | **0.0071** | 130,456,762 | 1,008,089 | 0.6354 | 129,449,094 | 421 | **0.0030** | 129,449,161 | 488 | **0.0023** |
| 10 | rs10822056 | 62,754,351 | 62,755,043 | 692 | **0.0065** | 62,729,283 | 25,068 | **0.0286** | 62,693,443 | 60,908 | 0.2570 | 62,693,310 | 61,041 | 0.1909 |
| 11 | rs10790459 | 121,767,808 | 121,780,992 | 13,184 | 0.0958 | 121,780,992 | 13184 | **0.0193** | 121,780,889 | 13,081 | 0.0572 | 121,780,911 | 13,103 | **0.0420** |
| 12 | rs150006321 | 66,069,074 | 66,067,134 | 1,940 | **0.0166** | 66,067,134 | 1,940 | **0.0021** | 66,067,464 | 1,610 | **0.0131** | 66,067,413 | 1,661 | **0.0094** |
| 14 | rs2764962 | 36,759,015 | 36,760,335 | 1,320 | **0.0135** | 36,760,335 | 1,320 | **0.0021** | 36,760,150 | 1,135 | **0.0078** | 36,760,189 | 1,174 | **0.0046** |
| 15 | rs9919974 | 40,045,261 | 40,077,321 | 4,310 | 0.2015 | 38,795,337 | 1,249,924 | 0.6525 | 40,046,730 | 1,469 | **0.0090** | 40,046,812 | 1,551 | **0.0057** |
| 18 | rs1563410 | 51,335,382 | 51,327,238 | 8,144 | 0.0645 | 50,959,150 | 376,232 | 0.3362 | 51,330,383 | 4,999 | **0.0287** | 51,327,467 | 7,915 | **0.0293** |
| 20 | rs6047482 | 21,559,045 | 21,555,863 | 3,182 | **0.0314** | 21,576,284 | 17,239 | **0.0239** | 21,556,092 | 2,953 | **0.0168** | 21,576,052 | 17,007 | 0.0523 |

[a]Genomic position based on NCBI Human Genome Build 38.

[b]Proportion of values less than or equal to distance of lead variant to at least 4 or more GGAA repeats, 13 or more GGAA repeats, 30 or more FLI1 binding and 45 or more FLI1 binding.

**Bold**-face indicates evidence of statistical enrichment for shorter distance to GGAA repeats or FLI1 binding compared to the rest of the chromosome.

GGAA repeats and FLI1 binding on the A-673 cell line are based on publicly available data from the Ewing Sarcoma Cell Line Atlas (ESCLA).

**Supplemental Table 10**. SNPs in newly discovered loci were evaluated in patient samples on Affymetrix and RNA-Seq technologies for their effect on gene expression. Effects (Beta), effect direction, and *P* values for the association between the risk allele in the GWAS results and expression quantitative trait loci (eQTLs) in Affymetrix and RNA-seq chips are displayed. There were 28 samples in common across both methods. Results display most significant (in Affymetrix set) SNP-eQTL association per each gene and only results with nominal *P* < 0.05 in one or both methods were listed. Bold-face indicates results under Bonferroni-corrected threshold (0.05/209 genes, *P* < 2.39×10$^{-4}$).

| Locus[a] | rsID | Position[a] | Referent Allele | Risk Allele | EAF[b] | Ewing GWAS Meta-analysis Result | | | | | | Gene | eQTL Analysis Result: Affymetrix microarray (N=113) | | | eQTL Analysis Result: RNA-Seq (N=48) | | |
|---|---|---|---|---|---|---|---|---|---|---|---|---|---|---|---|---|---|---|
| | | | | | | # Sets[c] | Beta | SE | OR | *P* value | Q | | Beta | Direction | *P* value | Beta | Direction | *P* value |
| 1p36.22 | rs9430161 | 10986798 | T | G | 0.7957 | 6 | 0.7420 | 0.0619 | 2.10 | 4.22E-33 | 0.5413 | CASZ1 | -0.3864 | **DOWN** | **6.41E-06** | -0.4814 | DOWN | 0.0640 |
| 1p36.22 | rs11576658 | 10977679 | T | C | 0.7784 | 6 | 0.5147 | 0.0558 | 1.67 | 2.74E-20 | 0.7886 | TARDBP | -0.4911 | **DOWN** | **1.81E-04** | -0.3525 | DOWN | 0.0210 |
| 1p36.22 | rs2003046 | 10972770 | A | C | 0.7754 | 6 | 0.4744 | 0.0545 | 1.61 | 3.20E-18 | 0.7738 | ANGPTL7 | -0.2806 | DOWN | 5.11E-04 | 0.0534 | UP | 0.5700 |
| 1p36.22 | rs9430161 | 10986798 | T | G | 0.7957 | 6 | 0.7420 | 0.0619 | 2.10 | 4.22E-33 | 0.5413 | PEX14 | -0.1188 | DOWN | 0.0293 | -0.1176 | DOWN | 0.2500 |
| 1p36.22 | rs9430161 | 10986798 | T | G | 0.7957 | 6 | 0.7420 | 0.0619 | 2.10 | 4.22E-33 | 0.5413 | C1orf127 | -0.0411 | DOWN | 0.0527 | -0.3337 | DOWN | 0.0410 |
| 1p36.22 | rs2003046 | 10972770 | A | C | 0.7754 | 6 | 0.4744 | 0.0545 | 1.61 | 3.20E-18 | 0.7738 | AGTRAP | 0.1486 | UP | 0.1550 | 0.3282 | UP | 0.0320 |
| 5q32.3 | rs10988 | 146120433 | T | C | 0.7229 | 6 | 0.2334 | 0.04995 | 1.26 | 2.96E-06 | 0.73367 | RBM27 | -1.2839 | **DOWN** | **4.23E-20** | -1.0581 | **DOWN** | **1.30E-05** |
| 5q32.3 | rs2063002 | 146188314 | G | A | 0.7179 | 6 | 0.2449 | 0.05004 | 1.28 | 9.87E-07 | 0.51881 | PLAC8L1 | -0.6480 | **DOWN** | **7.05E-06** | -0.1896 | DOWN | 0.25 |
| 5q32.3 | rs13173462 | 146242909 | C | T | 0.7338 | 6 | 0.2675 | 0.05143 | 1.31 | 1.97E-07 | 0.61253 | TCERG1 | -0.2654 | DOWN | 0.0051 | -0.1859 | DOWN | 0.3 |
| 5q32.3 | rs13173462 | 146242909 | C | T | 0.7338 | 6 | 0.2675 | 0.05143 | 1.31 | 1.97E-07 | 0.61253 | POU4F3 | -0.0303 | DOWN | 0.2664 | -0.3987 | DOWN | 9.20E-04 |
| 5q32.3 | rs2063002 | 146188314 | G | A | 0.7179 | 6 | 0.2449 | 0.05004 | 1.28 | 9.87E-07 | 0.51881 | STK32A | NA | NA | NA | -0.0834 | DOWN | 0.011 |
| 6p25.1 | rs1286037 | 6843500 | A | G | 0.2543 | 6 | 0.2794 | 0.0446 | 1.32 | 3.76E-10 | 0.2077 | RREB1 | 0.1365 | **UP** | **4.23E-05** | 0.1879 | UP | 0.0580 |
| 6p25.1 | rs1286037 | 6843500 | A | G | 0.2543 | 6 | 0.2794 | 0.0446 | 1.32 | 3.76E-10 | 0.2077 | DSP | 1.1097 | UP | 0.0042 | 0.0084 | UP | 0.9700 |
| 6p25.1 | rs1286037 | 6843500 | A | G | 0.2543 | 6 | 0.2794 | 0.0446 | 1.32 | 3.76E-10 | 0.2077 | NRN1 | 0.1399 | UP | 0.6711 | -0.7914 | DOWN | 0.0400 |
| 7q32.3 | rs350655 | 131028942 | A | C | 0.9031 | 5 | 0.6525 | 0.0922 | 1.92 | 1.49E-12 | 0.1517 | CPA4 | -0.0360 | DOWN | 0.0018 | 0.0452 | UP | 0.7200 |
| 7q32.3 | rs6978148 | 130993395 | C | T | 0.6520 | 6 | 0.2658 | 0.0448 | 1.30 | 3.05E-09 | 0.5177 | MKLN1 | 0.0981 | UP | 0.0211 | 0.1125 | UP | 0.1700 |
| 7q32.3 | rs765965 | 131019616 | C | A | 0.8562 | 6 | 0.3663 | 0.0665 | 1.44 | 3.60E-08 | 0.2346 | KLF14 | -0.0750 | DOWN | 0.0210 | NA | NA | NA |
| 7q32.3 | rs6978148 | 130993395 | C | T | 0.6520 | 6 | 0.2658 | 0.0448 | 1.30 | 3.05E-09 | 0.5177 | CEP41 | 0.1979 | UP | 0.0378 | 0.1503 | UP | 0.0880 |
| 7q32.3 | rs6978148 | 130993395 | C | T | 0.6520 | 6 | 0.2658 | 0.0448 | 1.30 | 3.05E-09 | 0.5177 | KLHDC10 | 0.1605 | UP | 0.0381 | 0.1225 | UP | 0.1900 |
| 7q32.3 | rs765965 | 131019616 | C | A | 0.8562 | 6 | 0.3663 | 0.0665 | 1.44 | 3.60E-08 | 0.2346 | CPA5 | 0.0391 | UP | 0.1500 | -0.1164 | DOWN | 0.0033 |
| 7q32.3 | rs6978148 | 130993395 | C | T | 0.6520 | 6 | 0.2658 | 0.0448 | 1.30 | 3.05E-09 | 0.5177 | TSGA13 | 0.0016 | UP | 0.7512 | 0.0084 | UP | 0.0190 |
| 7q32.3 | rs350655 | 131028942 | A | C | 0.9031 | 5 | 0.6525 | 0.0922 | 1.92 | 1.49E-12 | 0.1517 | PODXL | -0.0933 | DOWN | 0.8174 | -1.0792 | DOWN | 0.0170 |
| 8q24.21 | rs1897447 | 129449239 | A | G | 0.3564 | 6 | 0.2753 | 0.0414 | 1.32 | 2.98E-11 | 0.5009 | MYC | 0.2323 | UP | 0.0461 | 0.1687 | UP | 0.3900 |
| 10q21.2 | rs7915131 | 62658896 | T | C | 0.4402 | 6 | 0.3111 | 0.0411 | 1.36 | 3.99E-14 | 0.6324 | ADO | 0.4707 | **UP** | **9.60E-05** | 0.3090 | UP | 0.0450 |
| 10q21.2 | rs1848797 | 62793174 | A | G | 0.3871 | 6 | 0.4030 | 0.0420 | 1.50 | 8.55E-22 | 0.2044 | EGR2 | 1.0298 | UP | 0.0022 | 0.8437 | UP | 0.0240 |
| 10q21.2 | rs10995250 | 62637161 | A | G | 0.6555 | 6 | 0.3033 | 0.0439 | 1.35 | 4.85E-12 | 0.9344 | RTKN2 | -0.3370 | DOWN | 0.0047 | 0.0160 | UP | 0.8900 |
| 10q21.2 | rs224307 | 62841675 | A | G | 0.5950 | 6 | 0.3960 | 0.0439 | 1.49 | 1.82E-19 | 0.1879 | NRBF2 | 0.1852 | UP | 0.0069 | -0.0100 | DOWN | 0.9000 |
| 10q21.2 | rs7915131 | 62658896 | T | C | 0.4402 | 6 | 0.3111 | 0.0411 | 1.36 | 3.99E-14 | 0.6324 | JMJD1C-AS1 | -0.0313 | DOWN | 0.4182 | -0.0892 | DOWN | 0.0360 |
| 10q21.2 | rs10995239 | 62628605 | G | A | 0.6555 | 6 | 0.3025 | 0.0438 | 1.35 | 5.02E-12 | 0.9283 | REEP3 | -0.0278 | DOWN | 0.5397 | -0.4821 | DOWN | 0.0130 |
| 11q24.1 | rs10790459 | 121767808 | T | G | 0.7226 | 6 | 0.2589 | 0.0475 | 1.30 | 5.14E-08 | 0.57485 | SC5D | 0.0347 | UP | 0.0358 | -0.0387 | DOWN | 0.7800 |
| 11q24.1 | rs10790459 | 121767808 | T | G | 0.7226 | 6 | 0.2589 | 0.0475 | 1.30 | 5.14E-08 | 0.57485 | MIR100HG | 0.6519 | UP | 0.0437 | 0.6188 | UP | 0.0680 |
| 11q24.1 | rs7115520 | 121733948 | C | T | 0.7396 | 5 | 0.2016 | 0.0496 | 1.22 | 4.76E-05 | 0.77641 | SORL1 | 0.4479 | UP | 0.0484 | -0.1137 | DOWN | 0.3300 |
| 11q24.1 | rs7937934 | 121764503 | C | T | 0.7407 | 5 | 0.2001 | 0.0492 | 1.22 | 4.79E-05 | 0.89012 | SORL1 | 0.4479 | UP | 0.0484 | -0.1137 | DOWN | 0.3300 |
| 12q14.3 | rs10735933 | 66066014 | G | A | 0.6549 | 6 | 0.2117 | 0.0442 | 1.24 | 1.66E-06 | 0.59345 | TMBIM4 | 0.1632 | UP | 0.0080 | 0.0565 | UP | 0.4800 |
| 12q14.3 | rs1168745 | 66163180 | G | A | 0.7295 | 6 | 0.2268 | 0.0476 | 1.25 | 1.90E-06 | 0.32202 | MSRB3 | 0.1288 | UP | 0.5215 | 0.5704 | UP | 0.0490 |
| 12q14.3 | rs1168745 | 66163180 | G | A | 0.7295 | 6 | 0.2268 | 0.0476 | 1.25 | 1.90E-06 | 0.32202 | HMGA2 | 0.0732 | UP | 0.0194 | 0.1164 | UP | 0.7200 |
| 14q13.3 | rs2764962 | 36759015 | T | C | 0.5675 | 6 | 0.2314 | 0.0418 | 1.26 | 3.00E-08 | 0.8518 | PAX9 | -0.9001 | **DOWN** | **8.85E-06** | -0.7743 | DOWN | 0.0220 |
| 14q13.3 | rs2764962 | 36759015 | T | C | 0.5675 | 6 | 0.2314 | 0.0418 | 1.26 | 3.00E-08 | 0.8518 | MIPOL1 | -0.3510 | **DOWN** | **9.78E-06** | -0.2702 | DOWN | 0.0330 |
| 14q13.3 | rs2764962 | 36759015 | T | C | 0.5675 | 6 | 0.2314 | 0.0418 | 1.26 | 3.00E-08 | 0.8518 | SLC25A21 | -0.7701 | **DOWN** | **4.22E-05** | -0.4845 | DOWN | 0.0015 |
| 14q13.3 | rs2764962 | 36759015 | T | C | 0.5675 | 6 | 0.2314 | 0.0418 | 1.26 | 3.00E-08 | 0.8518 | SLC25A21-AS1 | -0.2954 | DOWN | 7.80E-04 | -0.4752 | **DOWN** | **8.30E-05** |
| 14q13.3 | rs848092 | 36799592 | G | A | 0.3405 | 6 | 0.2320 | 0.0419 | 1.26 | 3.12E-08 | 0.5936 | BRMS1L | -0.1159 | DOWN | 0.0172 | -0.0941 | DOWN | 0.2500 |
| 14q13.3 | rs2764962 | 36759015 | T | C | 0.5675 | 6 | 0.2314 | 0.0418 | 1.26 | 3.00E-08 | 0.8518 | NKX2-8 | -0.0037 | DOWN | 0.0937 | -0.0602 | DOWN | 0.0026 |
| 15q15.1 | rs8026641 | 40046660 | A | G | 0.2774 | 6 | 0.4879 | 0.0426 | 1.63 | 2.28E-30 | 0.3138 | SRP14 | -0.1444 | **DOWN** | **2.00E-04** | -0.1663 | DOWN | 0.0400 |
| 15q15.1 | rs652196 | 40083946 | A | G | 0.7534 | 6 | 0.4596 | 0.0529 | 1.58 | 3.75E-18 | 0.8293 | C15orf56 | -0.2428 | DOWN | 3.22E-04 | NA | NA | NA |
| 15q15.1 | rs8026641 | 40046660 | A | G | 0.2774 | 6 | 0.4879 | 0.0426 | 1.63 | 2.28E-30 | 0.3138 | BMF | -0.6036 | DOWN | 3.60E-04 | -0.7591 | DOWN | 0.0036 |
| 15q15.1 | rs12913170 | 40084007 | T | C | 0.8280 | 6 | 0.6827 | 0.0672 | 1.98 | 2.78E-24 | 0.0780 | CHST14 | -0.3736 | DOWN | 0.0038 | -0.1819 | DOWN | 0.1900 |
| 15q15.1 | rs8026641 | 40046660 | A | G | 0.2774 | 6 | 0.4879 | 0.0426 | 1.63 | 2.28E-30 | 0.3138 | CHAC1 | 0.1329 | UP | 0.0095 | 0.2700 | UP | 0.0250 |
| 15q15.1 | rs8042947 | 40033659 | G | A | 0.7233 | 6 | 0.2640 | 0.0480 | 1.30 | 3.71E-08 | 0.4360 | THBS1 | -0.5399 | DOWN | 0.0220 | -0.2853 | DOWN | 0.5000 |
| 15q15.1 | rs17722526 | 40028066 | A | G | 0.3494 | 6 | 0.3836 | 0.0414 | 1.47 | 1.94E-20 | 0.1115 | VPS18 | 0.0421 | UP | 0.0282 | 0.1243 | UP | 0.1600 |
| 15q15.1 | rs12913170 | 40084007 | T | C | 0.8280 | 6 | 0.6827 | 0.0672 | 1.98 | 2.78E-24 | 0.0780 | PAK6 | -0.4905 | DOWN | 0.0316 | NA | NA | NA |
| 15q15.1 | rs12164905 | 40008490 | T | G | 0.8234 | 6 | 0.3398 | 0.0590 | 1.40 | 8.24E-09 | 0.0500 | KNSTRN | 0.3715 | UP | 0.0462 | 0.3185 | UP | 0.1600 |
| 15q15.1 | rs937213 | 40029923 | T | C | 0.4212 | 6 | 0.3089 | 0.0411 | 1.36 | 5.47E-14 | 0.4473 | DISP2 | 0.2908 | UP | 0.0479 | 0.2294 | UP | 0.1100 |
| 15q15.1 | rs8026641 | 40046660 | A | G | 0.2774 | 6 | 0.4879 | 0.0426 | 1.63 | 2.28E-30 | 0.3138 | PPP1R14D | 0.0402 | UP | 0.0823 | 0.2320 | UP | 0.0300 |
| 15q15.1 | rs8042947 | 40033659 | G | A | 0.7233 | 6 | 0.2640 | 0.0480 | 1.30 | 3.71E-08 | 0.4360 | BUB1B | 0.3573 | UP | 0.1302 | -0.5773 | DOWN | 0.0180 |
| 15q15.1 | rs17722526 | 40028066 | A | G | 0.3494 | 6 | 0.3836 | 0.0414 | 1.47 | 1.94E-20 | 0.1115 | RAD51 | 0.0710 | UP | 0.1519 | 0.2678 | UP | 0.0330 |
| 15q15.1 | rs8026641 | 40046660 | A | G | 0.2774 | 6 | 0.4879 | 0.0426 | 1.63 | 2.28E-30 | 0.3138 | RPUSD2 | 0.0658 | UP | 0.4069 | 0.2180 | UP | 0.0360 |
| 18q21.2 | rs7232265 | 51289136 | G | A | 0.5140 | 6 | 0.2041 | 0.0404 | 1.23 | 4.28E-07 | 0.9095 | MEX3C | -0.2279 | **DOWN** | **5.75E-06** | -0.3942 | DOWN | 0.0190 |
| 18q21.2 | rs8096627 | 51296370 | C | A | 0.4988 | 6 | 0.2134 | 0.0403 | 1.24 | 1.23E-07 | 0.8943 | ME2 | -0.2082 | DOWN | 0.0032 | -0.2239 | DOWN | 0.0820 |
| 18q21.2 | rs7232265 | 51289136 | G | A | 0.5140 | 6 | 0.2041 | 0.0404 | 1.23 | 4.28E-07 | 0.9095 | MRO | -0.3032 | DOWN | 0.0034 | -0.1191 | DOWN | 0.2600 |
| 20p11.22 | rs911644 | 20985110 | C | T | 0.7829 | 6 | 0.2971 | 0.0531 | 1.35 | 2.20E-08 | 0.2556 | RALGAPA2 | -0.2148 | DOWN | 0.0098 | -0.1700 | DOWN | 0.3900 |
| 20p11.22 | rs2249048 | 21477034 | A | C | 0.6782 | 6 | 0.2944 | 0.0456 | 1.34 | 1.10E-10 | 0.8773 | PAX1 | 0.1113 | UP | 0.0272 | 0.2582 | UP | 0.3100 |
| 20p11.22 | rs6137387 | 21557266 | C | T | 0.8264 | 6 | 0.5215 | 0.0635 | 1.68 | 2.06E-16 | 0.7941 | LOC284788 | -0.0031 | DOWN | 0.4844 | -0.0073 | **DOWN** | **1.60E-05** |

[a]Cytogenic region and genomic position based on NCBI genome build 38.

[b]Effect allele frequencty (EAF) is based on dbGAP ALFA project European population.

[c]The number of GWAS sets the GWAS meta-analysis result is based on.

**Supplemental Table 11**: Log2 Fold Change(FC) in gene expression from *EWSR1::ETS* knockdown (KD) among ESCLA cell lines. Genes 1 Mb up or downstream from lead variant were selected. Negative values display downregulation of gene expression under KD. Positive values display upregulation after KD. *P* value is for a one sample t.test. Only nominally significant (*P* value < 0.05) results in the 18 cell line set are displayed. Bold-face represents *P* values below Bonferroni-corrected threshold ($P < 2.42×10^{-4}$). For each region, genes are sorted by statistical significance in 18 cell line set.

| Region | Gene | All 18 Cell Lines | | | | 5 cell lines with highest KD efficiency | | | |
|---|---|---|---|---|---|---|---|---|---|
| | | Mean LOG2FC | MedianLOG2FC | SD LOG2FC | *P* value | Mean LOG2FC | Median LOG2FC | SD LOG2FC | *P* value |
| 1p36.13 | PAX7 | -3.33 | -2.87 | 1.40 | **1.44E-08** | -3.96 | -4.08 | 0.74 | 0.0003 |
| | SDHB | -0.59 | -0.61 | 0.39 | **5.34E-06** | -0.80 | -0.77 | 0.31 | 0.0046 |
| | PADI2 | -1.74 | -1.60 | 1.29 | **2.60E-05** | -2.02 | -2.21 | 1.53 | 0.0414 |
| | ALDH4A1 | -0.83 | -0.86 | 0.71 | **1.13E-04** | -0.93 | -1.25 | 0.64 | 0.0320 |
| | KLHDC7A | -0.22 | -0.20 | 0.22 | 5.52E-04 | -0.25 | -0.18 | 0.30 | 0.1320 |
| | IGSF21 | -1.10 | -0.87 | 1.17 | 9.09E-04 | -1.07 | -0.73 | 1.34 | 0.1498 |
| | MFAP2 | 0.83 | 0.85 | 0.88 | 9.28E-04 | 1.60 | 1.17 | 0.83 | 0.0125 |
| | RCC2 | -0.15 | -0.09 | 0.28 | 0.0350 | -0.25 | -0.23 | 0.31 | 0.1444 |
| 1p36.22 | EXOSC10 | -0.53 | -0.51 | 0.26 | **1.00E-07** | -0.69 | -0.61 | 0.25 | 0.0036 |
| | SRM | -0.72 | -0.57 | 0.55 | **3.33E-05** | -0.50 | -0.53 | 0.30 | 0.0199 |
| | TARDBP | -0.34 | -0.25 | 0.26 | **3.58E-05** | -0.44 | -0.26 | 0.43 | 0.0806 |
| | DRAXIN | 1.00 | 0.91 | 1.02 | 6.26E-04 | 0.69 | 1.20 | 0.84 | 0.1396 |
| | PLOD1 | 0.72 | 0.51 | 0.77 | 0.0010 | 1.15 | 0.94 | 1.08 | 0.0753 |
| | UBIAD1 | -0.47 | -0.54 | 0.51 | 0.0011 | -0.44 | -0.60 | 0.42 | 0.0796 |
| | AGTRAP | -0.55 | -0.56 | 0.63 | 0.0017 | -0.85 | -0.93 | 0.60 | 0.0333 |
| | MTOR | -0.28 | -0.27 | 0.37 | 0.0049 | -0.52 | -0.54 | 0.28 | 0.0146 |
| | KIAA2013 | -0.19 | -0.16 | 0.26 | 0.0072 | -0.21 | -0.10 | 0.19 | 0.0669 |
| | RBP7 | -0.27 | -0.25 | 0.37 | 0.0074 | -0.08 | -0.05 | 0.26 | 0.5276 |
| | PEX14 | -0.28 | -0.25 | 0.42 | 0.0120 | -0.25 | -0.32 | 0.32 | 0.1589 |
| | MAD2L2 | -0.23 | -0.15 | 0.43 | 0.0344 | -0.57 | -0.67 | 0.51 | 0.0662 |
| 5q32.3 | PLAC8L1 | -0.21 | -0.19 | 0.20 | 4.00E-04 | -0.30 | -0.29 | 0.29 | 0.0768 |
| | TCERG1 | -0.30 | -0.26 | 0.29 | 4.44E-04 | -0.44 | -0.37 | 0.38 | 0.0569 |
| | GPR151 | -0.23 | -0.19 | 0.28 | 0.0023 | -0.30 | -0.19 | 0.38 | 0.1580 |
| | PPP2R2B | -0.37 | -0.21 | 0.50 | 0.0053 | -0.42 | -0.23 | 0.68 | 0.2381 |
| | RBM27 | -0.27 | -0.22 | 0.48 | 0.0297 | -0.68 | -0.41 | 0.70 | 0.0959 |
| 6p22.1 | GNL1 | -0.41 | -0.44 | 0.34 | **7.36E-05** | -0.48 | -0.49 | 0.32 | 0.0299 |
| | GABBR1 | 0.35 | 0.27 | 0.36 | 7.14E-04 | 0.38 | 0.35 | 0.12 | 0.0019 |
| | OR12D2 | -0.18 | -0.23 | 0.18 | 7.25E-04 | -0.17 | -0.22 | 0.15 | 0.0644 |
| | HLA-A | 0.57 | 0.43 | 0.80 | 0.0074 | 1.14 | 0.86 | 1.04 | 0.0707 |
| | OR2H2 | -0.14 | -0.17 | 0.21 | 0.0085 | -0.23 | -0.35 | 0.23 | 0.0872 |
| | RPP21 | -0.32 | -0.34 | 0.46 | 0.0090 | -0.34 | -0.52 | 0.44 | 0.1617 |
| | PPP1R11 | -0.24 | -0.24 | 0.35 | 0.0101 | -0.24 | -0.28 | 0.34 | 0.1840 |
| | HLA-F | 0.31 | 0.32 | 0.48 | 0.0125 | 0.59 | 0.54 | 0.72 | 0.1443 |
| | ZNF311 | -0.18 | -0.17 | 0.29 | 0.0153 | -0.08 | -0.15 | 0.17 | 0.3385 |
| | OR14J1 | -0.14 | -0.09 | 0.23 | 0.0172 | -0.12 | -0.09 | 0.19 | 0.2115 |
| | TRIM27 | -0.15 | -0.14 | 0.25 | 0.0195 | -0.08 | -0.14 | 0.13 | 0.2216 |
| | HLA-L | 0.39 | 0.39 | 0.72 | 0.0327 | 0.84 | 0.73 | 1.08 | 0.1559 |
| | TRIM26 | 0.17 | 0.10 | 0.32 | 0.0415 | 0.21 | 0.11 | 0.23 | 0.1082 |
| | OR12D3 | -0.14 | -0.10 | 0.27 | 0.0432 | -0.19 | -0.11 | 0.20 | 0.1052 |
| 6p25.1 | SNRNP48 | -0.42 | -0.35 | 0.32 | **3.58E-05** | -0.45 | -0.36 | 0.43 | 0.0812 |
| | RREB1 | -0.43 | -0.33 | 0.33 | **4.10E-05** | -0.63 | -0.60 | 0.34 | 0.0149 |
| | BMP6 | -0.77 | -0.79 | 0.83 | 0.0011 | -0.39 | -0.69 | 0.88 | 0.3774 |
| | CAGE1 | -0.14 | -0.10 | 0.18 | 0.0044 | -0.17 | -0.24 | 0.18 | 0.0934 |
| | NRN1 | -0.86 | -0.75 | 1.32 | 0.0132 | -1.64 | -1.33 | 0.93 | 0.0166 |
| | RIOK1 | -0.35 | -0.49 | 0.62 | 0.0278 | -0.64 | -0.84 | 0.51 | 0.0501 |
| 7q32.3 | TMEM209 | -0.43 | -0.39 | 0.27 | **3.74E-06** | -0.46 | -0.39 | 0.24 | 0.0139 |
| | ZC3HC1 | -0.32 | -0.27 | 0.33 | 6.23E-04 | -0.46 | -0.39 | 0.44 | 0.0826 |
| | MKLN1 | -0.25 | -0.15 | 0.36 | 0.0076 | -0.49 | -0.34 | 0.50 | 0.0950 |
| | SSMEM1 | -0.11 | -0.10 | 0.18 | 0.0150 | -0.14 | -0.08 | 0.12 | 0.0690 |
| | KLHDC10 | -0.21 | -0.28 | 0.34 | 0.0187 | -0.15 | -0.23 | 0.36 | 0.3927 |
| 8q24.21 | MYC | -0.48 | -0.44 | 0.78 | 0.0180 | -0.30 | -0.61 | 0.73 | 0.4095 |
| | GSDMC | -0.01 | 0.01 | 0.24 | 0.8629 | -0.06 | 0.01 | 0.32 | 0.7200 |
| 10q21.2 | ARID5B | 2.02 | 1.87 | 0.87 | **1.75E-08** | 2.56 | 2.00 | 0.90 | 0.0032 |
| | JMJD1C | 0.72 | 0.62 | 0.49 | **9.47E-06** | 0.87 | 0.83 | 0.33 | 0.0043 |
| | EGR2 | -1.59 | -1.57 | 1.71 | 0.0011 | -1.18 | -1.69 | 2.19 | 0.2948 |
| | NRBF2 | -0.33 | -0.28 | 0.38 | 0.0020 | -0.51 | -0.56 | 0.24 | 0.0085 |
| | RTKN2 | -0.66 | -0.38 | 0.91 | 0.0067 | -0.75 | -0.64 | 0.87 | 0.1251 |
| | REEP3 | 0.32 | 0.10 | 0.55 | 0.0254 | 0.21 | -0.14 | 0.69 | 0.5418 |
| | MIR1296 | 0.21 | 0.13 | 0.37 | 0.0282 | 0.10 | 0.05 | 0.25 | 0.4303 |
| | ADO | -0.27 | -0.13 | 0.52 | 0.0410 | -0.41 | -0.40 | 0.62 | 0.2126 |
| 11q24.1 | TECTA | 0.32 | 0.20 | 0.40 | 0.0035 | 0.44 | 0.68 | 0.48 | 0.1146 |
| | TBCEL | 0.37 | 0.46 | 0.53 | 0.0084 | 0.40 | 0.45 | 0.69 | 0.2650 |
| | UBASH3B | 0.35 | 0.14 | 0.67 | 0.0405 | 0.69 | 0.47 | 1.11 | 0.2356 |
| | GRIK4 | 0.31 | 0.16 | 0.61 | 0.0419 | 0.77 | 0.22 | 0.99 | 0.1580 |
| 12q14.3 | LEMD3 | 0.21 | 0.15 | 0.23 | 0.0011 | 0.18 | 0.05 | 0.26 | 0.2040 |
| | MSRB3 | 0.94 | 0.61 | 1.09 | 0.0019 | 1.24 | -0.06 | 1.84 | 0.2055 |
| | HMGA2 | 0.80 | 0.48 | 0.99 | 0.0033 | 1.09 | 0.88 | 1.34 | 0.1431 |
| | LLPH | -0.21 | -0.25 | 0.31 | 0.0107 | -0.31 | -0.40 | 0.19 | 0.0206 |
| | GRIP1 | 0.55 | 0.45 | 0.84 | 0.0123 | 0.38 | 0.39 | 0.61 | 0.2344 |
| | RPSAP52 | 0.13 | 0.11 | 0.21 | 0.0190 | 0.16 | 0.12 | 0.23 | 0.1951 |
| 14q13.3 | SFTA3 | -0.10 | -0.10 | 0.17 | 0.0235 | -0.14 | -0.15 | 0.06 | 0.0056 |
| | SLC25A21 | -0.40 | -0.48 | 0.69 | 0.0261 | -0.80 | -0.72 | 0.77 | 0.0822 |
| 15q15.1 | KNSTRN | -0.88 | -0.77 | 0.64 | **1.92E-05** | -1.19 | -1.07 | 0.92 | 0.0449 |
| | INO80 | -0.33 | -0.30 | 0.24 | **2.74E-05** | -0.49 | -0.48 | 0.19 | 0.0042 |
| | RAD51 | -1.16 | -0.98 | 0.92 | **5.64E-05** | -1.41 | -1.46 | 0.81 | 0.0174 |
| | RMDN3 | -0.37 | -0.33 | 0.33 | **1.93E-04** | -0.51 | -0.35 | 0.39 | 0.0448 |
| | BUB1B | -1.14 | -0.87 | 1.09 | 3.47E-04 | -1.31 | -0.94 | 1.39 | 0.1025 |
| | RHOV | -0.21 | -0.18 | 0.23 | 0.0012 | -0.25 | -0.18 | 0.20 | 0.0488 |
| | INAFM2 | 0.44 | 0.31 | 0.51 | 0.0017 | 0.14 | 0.09 | 0.22 | 0.2186 |
| | RPUSD2 | -0.32 | -0.35 | 0.37 | 0.0022 | -0.52 | -0.44 | 0.20 | 0.0045 |
| | THBS1 | 1.88 | 0.97 | 2.25 | 0.0025 | 2.54 | 0.79 | 3.18 | 0.1487 |
| | PLCB2 | 0.33 | 0.21 | 0.40 | 0.0026 | 0.43 | 0.44 | 0.39 | 0.0701 |
| | BAHD1 | -0.16 | -0.17 | 0.24 | 0.0102 | -0.13 | -0.22 | 0.18 | 0.1712 |
| | EIF2AK4 | 0.24 | 0.19 | 0.36 | 0.0109 | 0.29 | 0.35 | 0.59 | 0.3352 |
| | SRP14 | 0.14 | 0.10 | 0.21 | 0.0122 | 0.02 | 0.00 | 0.15 | 0.7486 |
| | GPR176 | 0.66 | 0.52 | 1.00 | 0.0127 | 0.83 | 0.98 | 1.11 | 0.1706 |
| | DNAJC17 | -0.19 | -0.20 | 0.29 | 0.0141 | -0.24 | -0.34 | 0.28 | 0.1205 |
| | ZFYVE19 | -0.21 | -0.27 | 0.35 | 0.0197 | -0.35 | -0.60 | 0.37 | 0.0977 |
| | IVD | 0.21 | 0.19 | 0.41 | 0.0407 | -0.01 | 0.00 | 0.39 | 0.9520 |
| | VPS18 | 0.16 | 0.13 | 0.31 | 0.0427 | 0.14 | 0.21 | 0.38 | 0.4475 |
| 18q21.2 | MRO | -0.95 | -0.91 | 0.63 | **6.60E-06** | -1.12 | -0.92 | 0.29 | 0.0010 |
| | MEX3C | 0.49 | 0.48 | 0.41 | **8.34E-05** | 0.57 | 0.49 | 0.19 | 0.0024 |
| | SKA1 | -0.94 | -0.80 | 0.84 | **1.72E-04** | -0.83 | -0.48 | 1.04 | 0.1511 |
| 20p11.22 | NKX2-2 | -0.87 | -0.68 | 0.83 | 3.49E-04 | -1.47 | -0.70 | 1.29 | 0.0640 |
| | RALGAPA2 | 0.56 | 0.54 | 0.56 | 5.66E-04 | 0.11 | -0.08 | 0.47 | 0.6260 |
| | XRN2 | -0.15 | -0.16 | 0.17 | 0.0018 | -0.17 | -0.08 | 0.18 | 0.0939 |

**Supplemental Table 12**. Summary of integrative analysis identifying potential etiological mechanisms and target genes for identified Ewing sarcoma susceptibility loci. Genes that reach statistical significance in Bonferroni-corrected threshold (or the top gene when none reached statistical significance) are listed for eQTL or *EWSR1-ETS* Knockdown expression. Nominally significant genes are displayed in grey font. The top gene or statistically significant genes in one method are displayed for the other method if the results were nominally significant.

| Locus | GGAA enrichment | *FLI1* binding *enrichment* | GWAS risk allele on eQTL | | *EWSR1-FLI* Knockdown Expression change | | H3K27ac Hi-ChIP interaction between ≥ 4 GGAA mSat and gene promoter | CTCF Hi-ChIP interaction between ≥ 4 GGAA mSat and gene promoter |
|---|---|---|---|---|---|---|---|---|
| | | | eQTL genes | eQTL Effect | KD genes | KD effect | | |
| 1p36.22 | X | X | *TARDBP* | DOWN | *TARDBP* | DOWN | X | - |
| | | | *CASZ1* | DOWN | - | - | - | - |
| | | | - | - | *SRM* | DOWN | - | X |
| | | | - | - | *EXOSC10* | DOWN | - | - |
| 1p36.13 | - | - | - | - | *PAX7* | DOWN | X | - |
| | | | - | - | *SDHB* | DOWN | - | - |
| | | | - | - | *PADI2* | DOWN | X | X |
| | | | - | - | *ALDH4A1* | DOWN | X | - |
| 5q32.3 | - | - | *RBM27* | DOWN | *RBM27* | DOWN | X | - |
| | | | *PLAC8L1* | DOWN | *PLAC8L1* | DOWN | - | - |
| 6p25.1 | X | X | *RREB1* | UP | *RREB1* | DOWN | - | - |
| | | | - | - | *SNRNP48* | DOWN | - | - |
| 6p22.1 | - | - | - | - | *GNL1* | DOWN | - | X |
| 7q32.3 | X | X | *CPA4* | DOWN | - | - | - | - |
| | | | - | - | *TMEM209* | DOWN | - | - |
| 8q24.21 | X | X | *MYC* | UP | *MYC* | DOWN | X | - |
| 10q21.2 | X | - | *ADO* | UP | *ADO* | DOWN | - | - |
| | | | *EGR2* | UP | *EGR2* | DOWN | - | - |
| | | | - | - | *ARID5B* | UP | X | - |
| | | | *JMJD1C* | DOWN | *JMJD1C* | UP | - | X |
| 11q24.1 | X | X | *SC5D* | UP | - | - | - | - |
| | | | - | - | *TECTA* | UP | - | - |
| 12q14.3 | X | X | *TMBIM4* | UP | - | - | - | - |
| | | | - | - | *LEMD3* | UP | - | - |
| 14q13.3 | X | X | *PAX9* | DOWN | - | - | - | X |
| | | | *MIPOL1* | DOWN | - | - | - | - |
| | | | *SLC25A21* | DOWN | *SLC25A21* | DOWN | - | - |
| | | | - | - | *SFTA3* | DOWN | - | - |
| 15q15.1 | - | X | *SRP14* | DOWN | *SRP14* | UP | - | - |
| | | | *KNSTRN* | UP | *KNSTRN* | DOWN | X | - |
| | | | - | - | *INO80* | DOWN | - | - |
| | | | *RAD51* | UP | *RAD51* | DOWN | - | - |
| | | | - | - | *RMDN3* | DOWN | - | - |
| 18q21.2 | - | X | *MEX3C* | DOWN | *MEX3C* | UP | - | - |
| | | | *MRO* | DOWN | *MRO* | DOWN | - | - |
| | | | - | - | *SKA1* | DOWN | - | - |
| 20p11.22 | X | X | *RALGAPA2* | DOWN | *RALGAPA2* | UP | - | - |
| | | | - | - | *NKX2-2* | DOWN | X | X |
