## Supplemental Figures for "Genome-wide association study meta-analysis identifies susceptibility loci informing Ewing sarcoma etiology and potential mechanisms of risk"

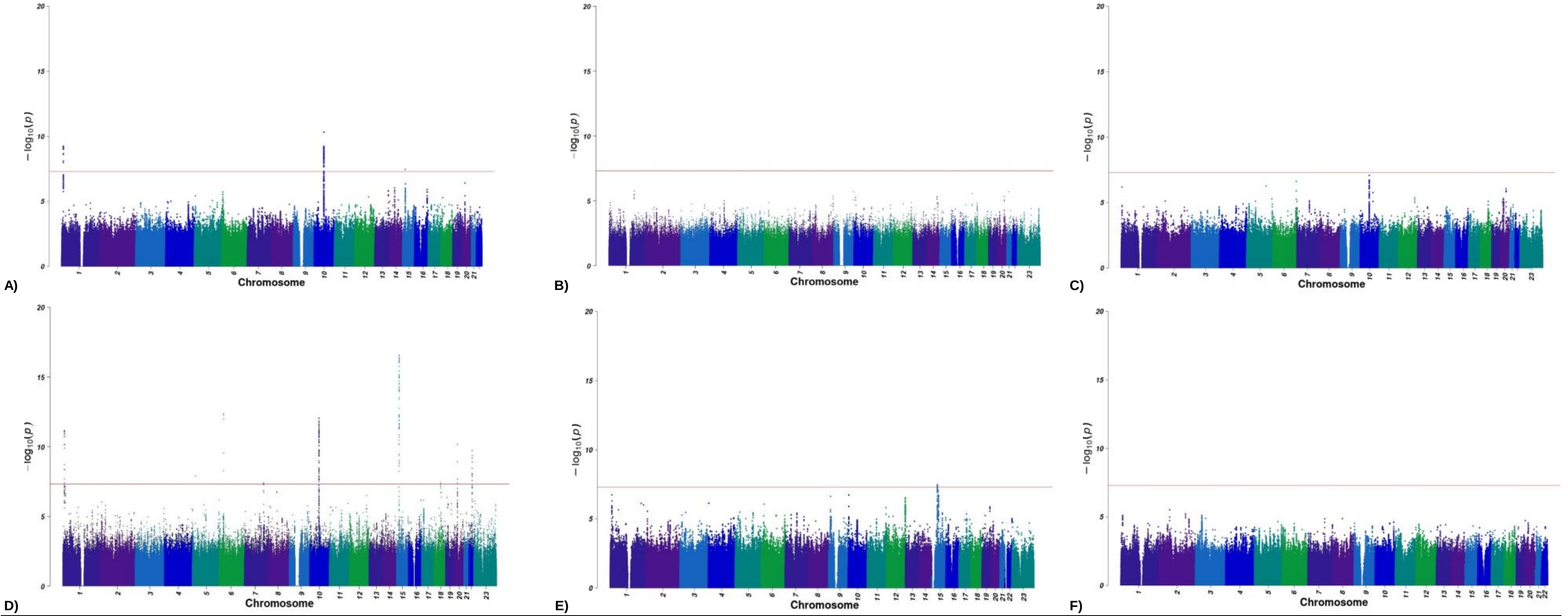


**Supplemental Figure 1**. Manhattan plots of -log10 *P* values for the association of each SNP with EwS risk in each GWAS set prior to meta-analysis. Association *P* values for each tested genetic variant are plotted. Chromosomes are plotted sequentially across the x-axis with the scale proportional to chromosomal size. Colors are used to visualize differences in chromosome. The red line indicates genome-wide significance ( *P* <5×10-8). Variants with < 1% minor allele frequency were excluded from each study's GWAS. A) Institut Curie GWAS 1 set, B) Omni Set, C) CCSS, D) GSA Set, E) CESS, F) SJ Life Study. The estimated genomic inflation was lambdas ranged from 0.98 to 1.03.

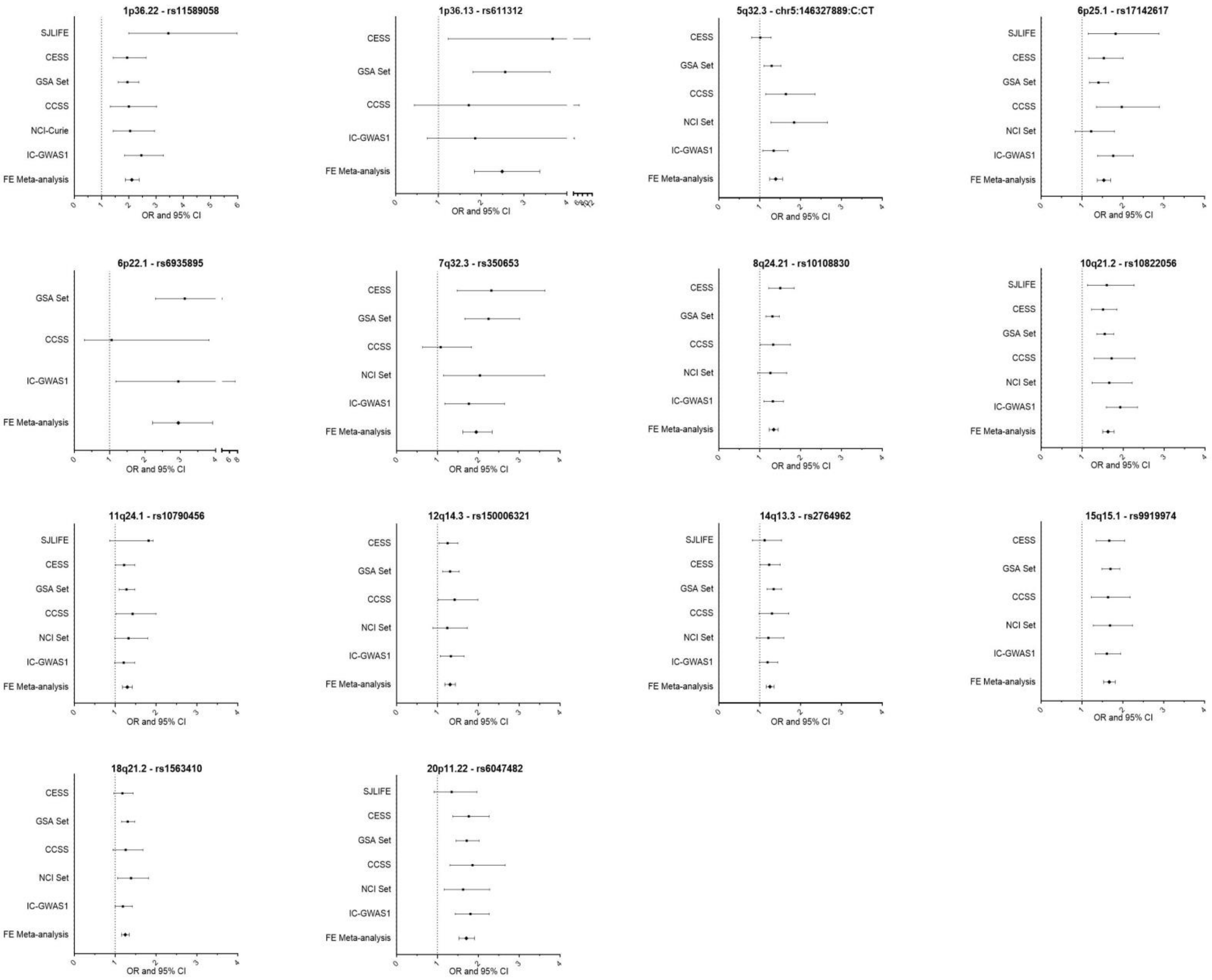


**Supplemental Figure 2**. Odds Ratio (OR) and 95% Confidence Interval (CI) for the lead variant for each genome wide significant locus in the Fixed Effects (FE) meta-analysis and across each study. Effects are not plotted when a lead variant was not present within the individual study.

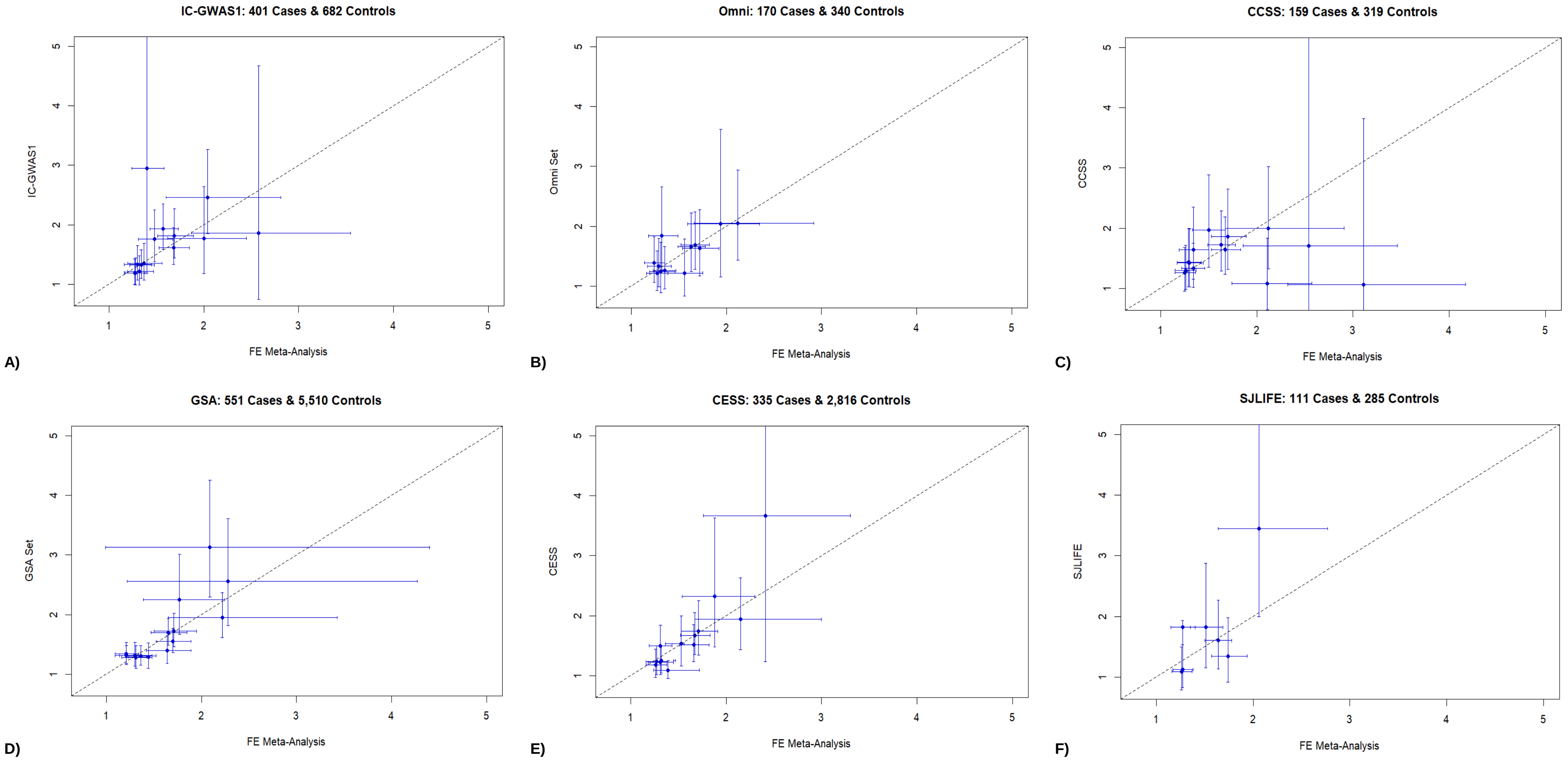


**Supplemental Figure 3**. Leave-one-out (LOO) comparison of Odds Ratios (OR) and 95% Confidence Intervals for Fixed Effects (FE) meta-analysis without each study and the study-specific effect. Each panel displays a different study and the FE meta-analysis without that study. Detailed results for variants in each study are provided in Supplemental Table 3. LOO meta-analysis results are given in **Supplemental Table 5**.

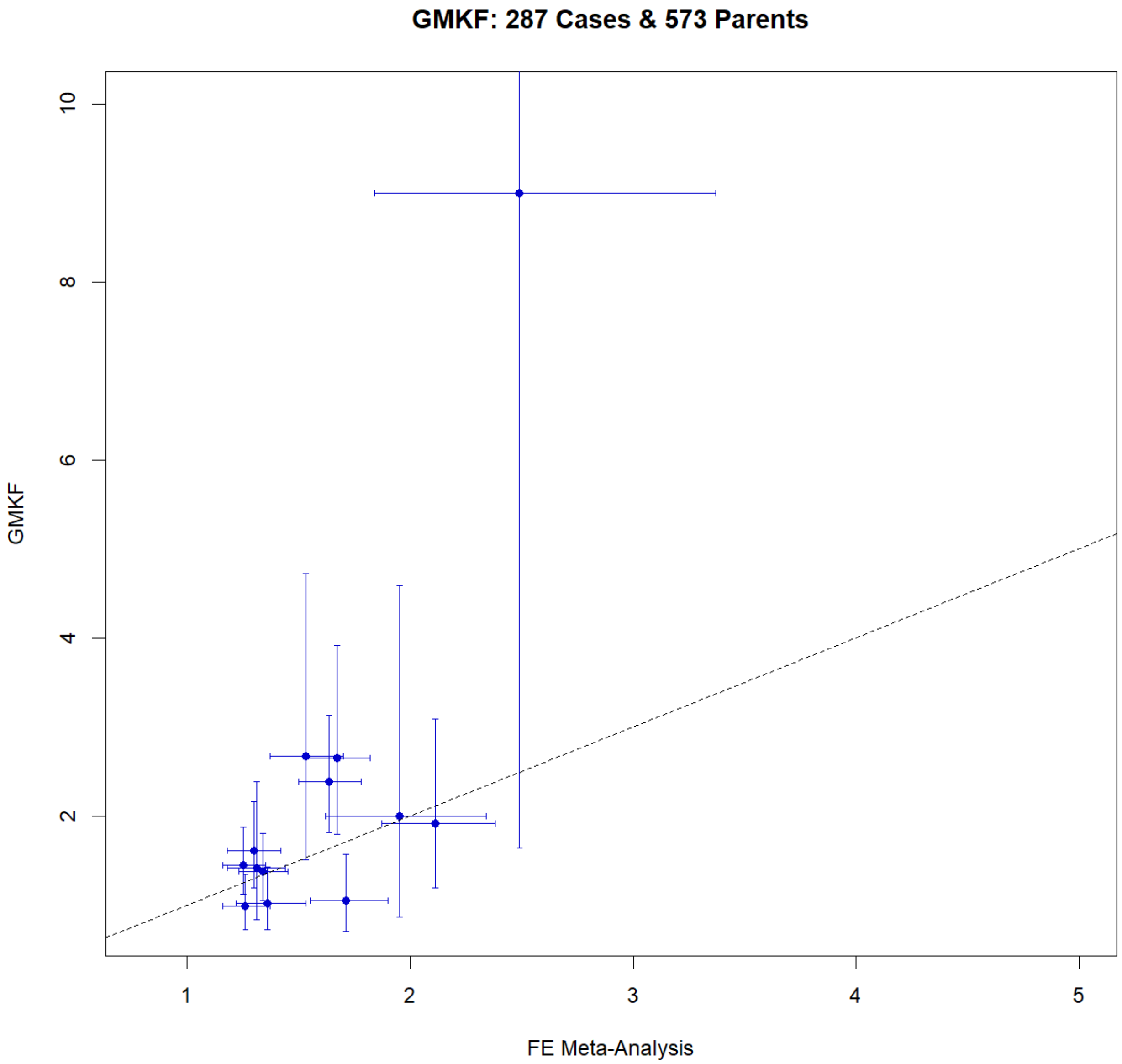


**Supplemental Figure 4**. The transmission disequilibrium test (TDT) odds ratio (OR) and 95% confidence interval (CI) in GMKF trio set for each variant in the main meta-analysis GWAS is plotted against the fixed effects meta-analysis OR and 95%CI. Detailed results for the variants in GMKF are given in **Supplemental Table 4**.

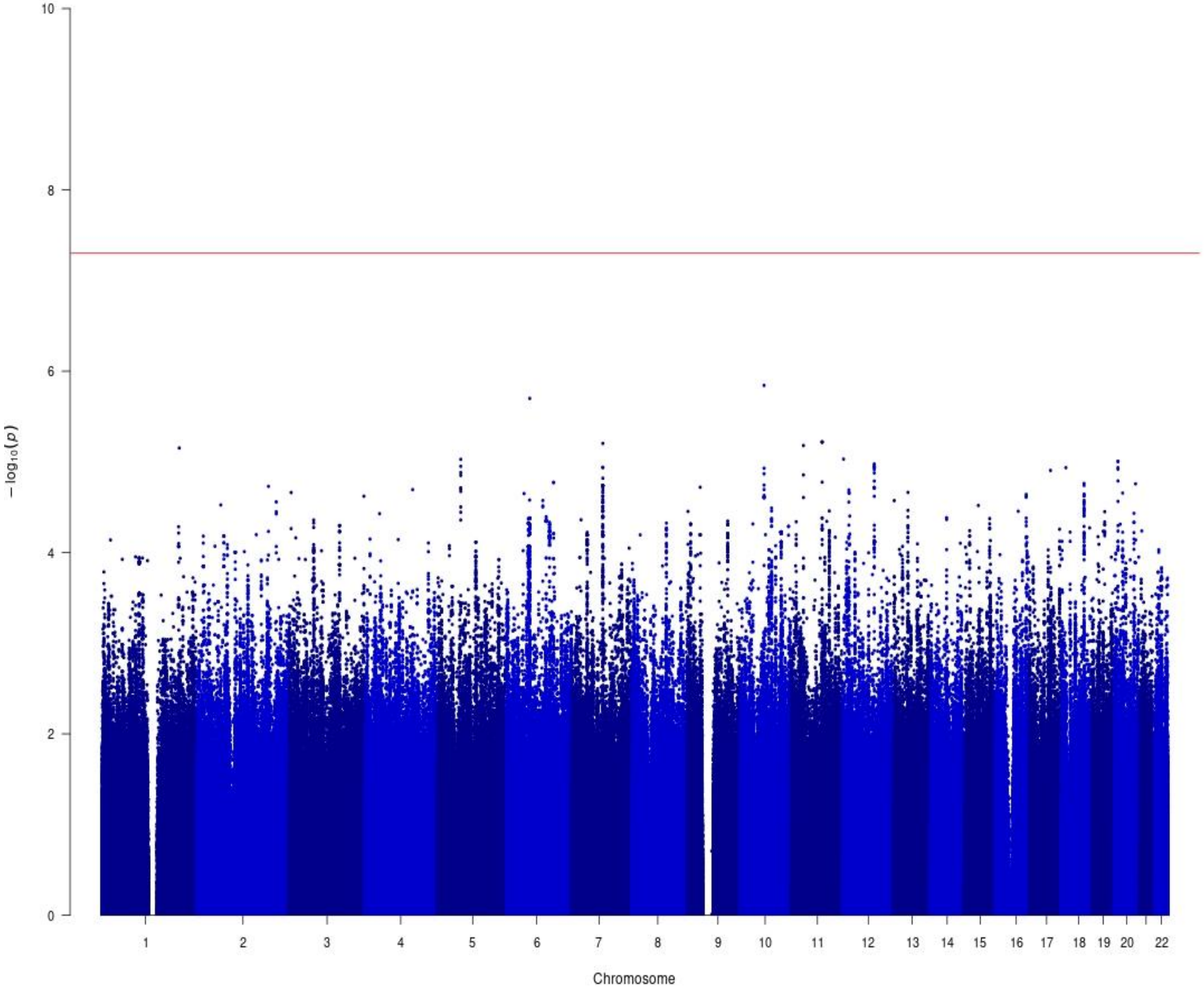


**Supplemental Figure 5**. Manhattan plot for fixed effects meta-analysis –log10 *P* values for the association of each SNP with sex among 709 males and 559 females with EwS. Association p-values for each tested genetic variant are plotted. Chromosomes are plotted sequentially across the x-axis with the scale proportional to chromosomal size. Colors are used to visualize differences in chromosome. The red line indicates genome-wide significance (*P*<5×10-8). Variants with < 1% minor allele frequency were excluded from each study's results prior to meta-analysis. Analysis included 8,839,636 SNPs. Variants with heterogeneity Q<0.05 were excluded from Manhattan plot. The estimated autosomal genomic inflation was lambda=0.96. P values for meta-analysis discovered loci are in **Table 2**. Phenotype distribution by study is provided in **Supplemental Table 8**.

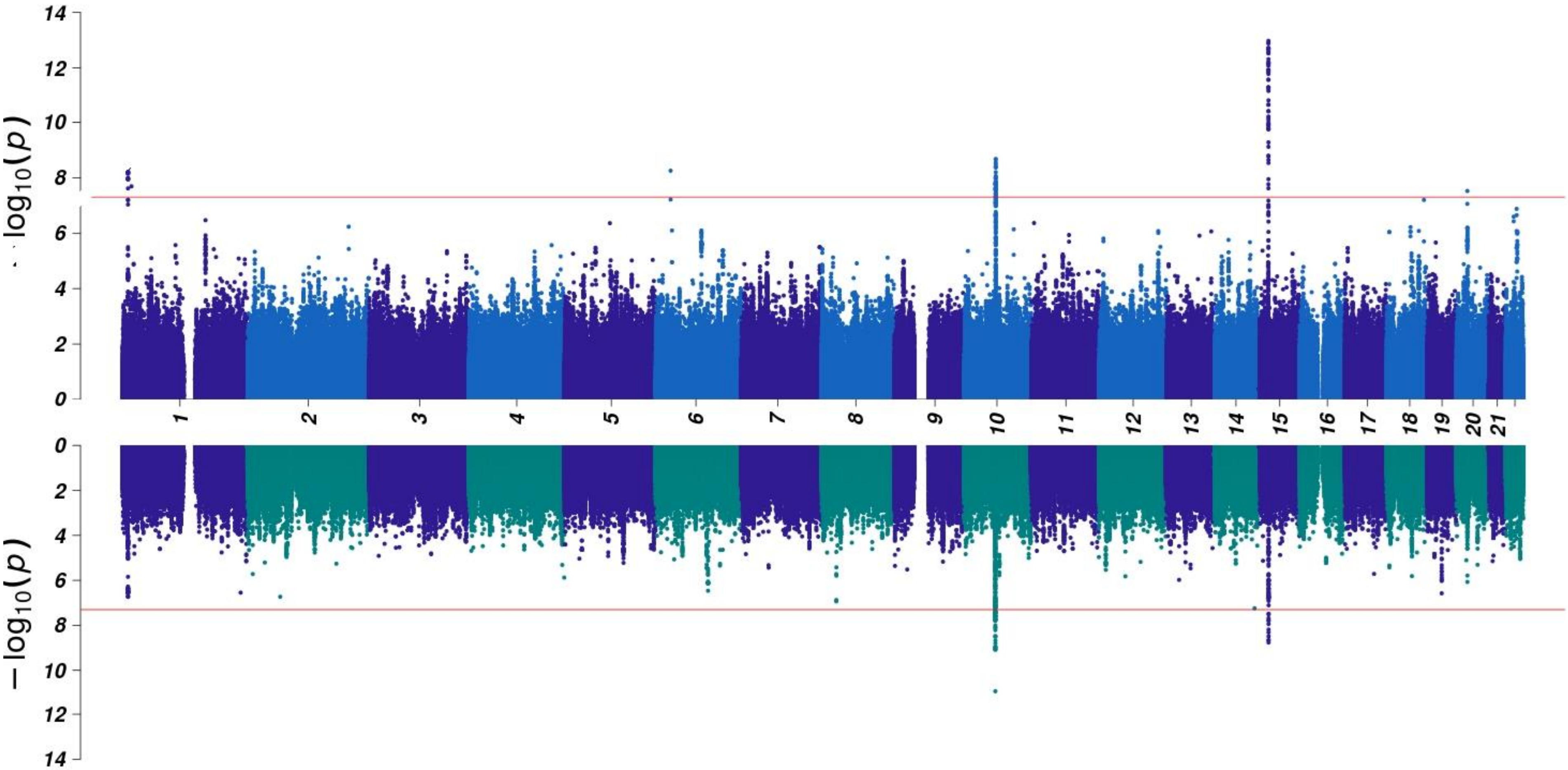


**Supplemental Figure 6**. Miami plot for fixed effects meta-analysis –log10 *P* values for the association of each SNP with EwS stratified by sex. Male EwS cases (N=378) and controls (N=2,618) are plotted on the top and female EwS cases (N=332) and controls (N=3,207) are plotted on the bottom. Association *P* values for each tested genetic variant are plotted. Chromosomes are plotted sequentially across the x-axis with the scale proportional to chromosomal size. Colors are used to visualize differences in chromosome. The red lines indicate genome-wide significance (*P* <5×10-8). Variants with < 1% minor allele frequency were excluded from each study's results prior to meta-analysis. Analysis included 8,443,513 variants in the male GWAS and 8,467,202 variants in the female GWAS. Variants with heterogeneity Q<0.05 were excluded from Manhattan plot. The estimated autosomal genomic inflation for lambda was 1.02 in the male GWAS and 1.00 in the female GWAS.

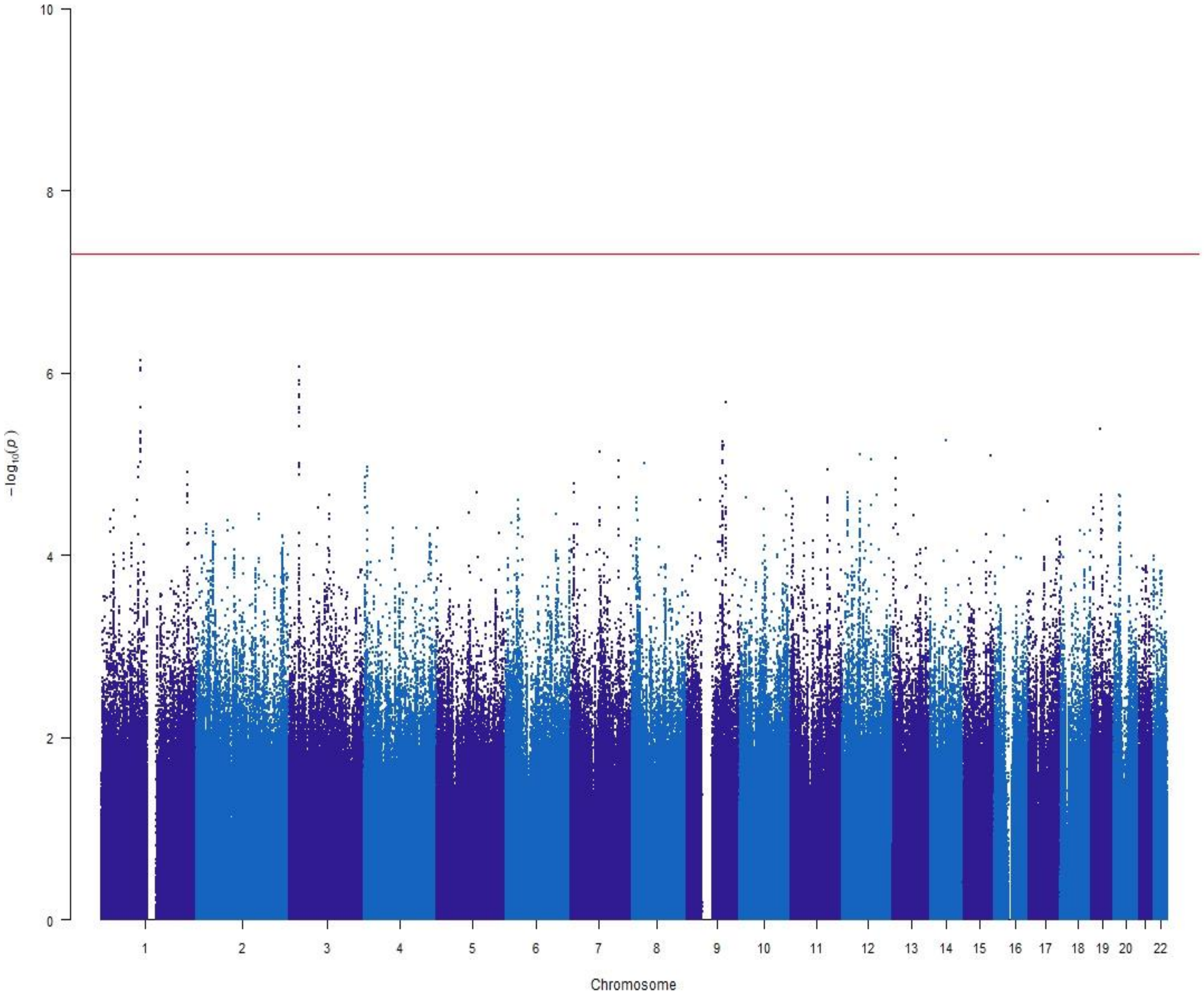


**Supplemental Figure 7**. Manhattan plot for fixed effects meta-analysis –log10 *P* values for the association of each SNP with metastasis at diagnosis (Yes/No) among 268 EwS cases with metastasis and 666 EwS cases without metastasis at diagnosis. Association *P* values for each tested genetic variant are plotted. Chromosomes are plotted sequentially across the x-axis with the scale proportional to chromosomal size. Colors are used to visualize differences in chromosome. The red line indicates genome-wide significance ($P$<5×10-8). Variants with < 1% minor allele frequency were excluded from each study's results prior to meta-analysis. Variants with heterogeneity Q<0.05 were excluded from Manhattan plot. The autosomal genomic inflation factor was lambda=0.96. Analysis included 8,541,107 variants. Phenotype distribution by study is provided in **Supplemental Table 8**.

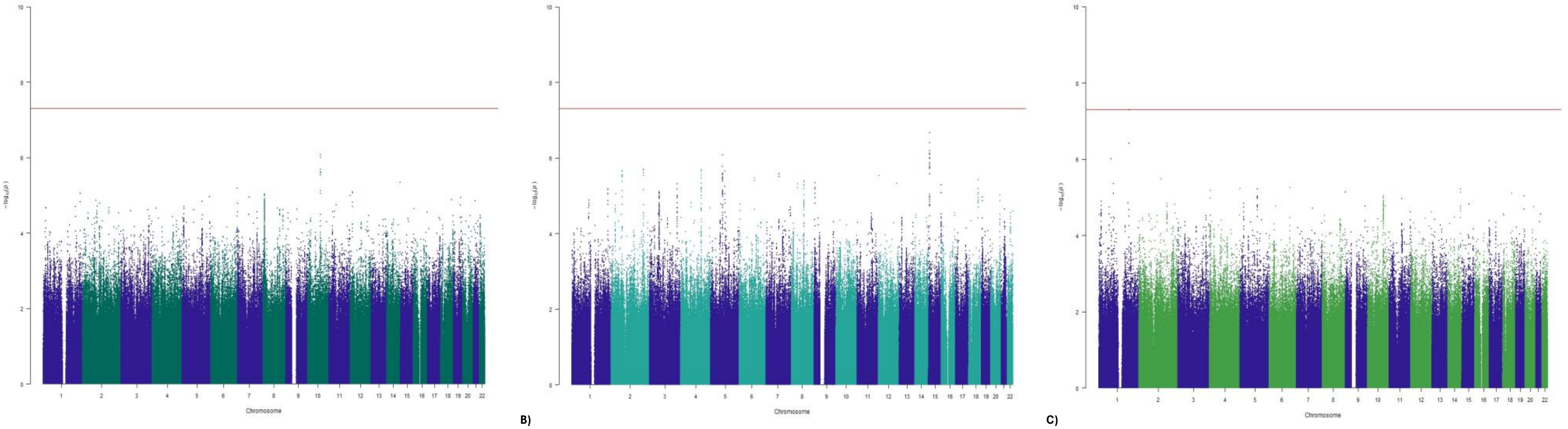


A) B) C)

**Supplemental Figure 8**. Manhattan plots for fixed effects meta-analysis –log10 *P* values for the association of each SNP with age at diagnosis. Association p-values for each tested genetic variant are plotted. Chromosomes are plotted sequentially across the x-axis with the scale proportional to chromosomal size. Colors are used to visualize differences in chromosome. The red line indicates genome-wide significance ($P < 5\times10\text{-}8$). Variants with < 1% minor allele frequencies were excluded from each study's results prior to meta-analysis. Variants with heterogeneity Q<0.05 were excluded from Manhattan plot. Three age phenotypes were evaluated **A)** young children (<10) vs. teens and adults (≥10), **B)** before puberty completion (≤15) vs. after puberty (>15), and **C)** pediatric onset (<20) vs. adult onset (≥20). Analysis included 8,755,664, 8,824,193, and 8,657,897 variants in phenotypes A-C respectively. Genomic inflation lambda=0.96-0.99. Phenotype distribution by study is provided in **Supplemental Table 8**.
