## Extended Data Figures for "Genome-wide association study meta-analysis identifies susceptibility loci informing Ewing sarcoma etiology and potential mechanisms of risk"

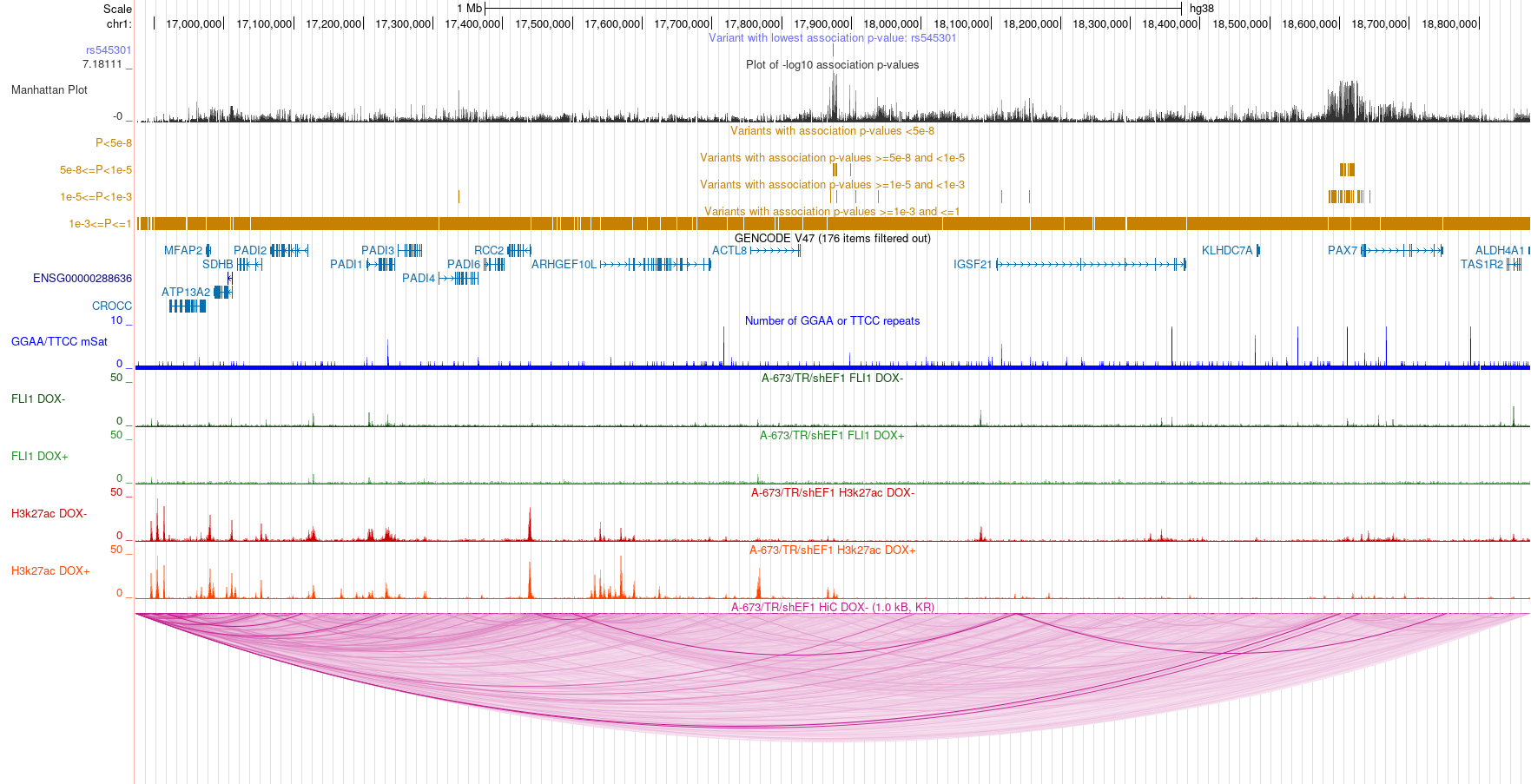

**Extended Data Figure 1: 1p36.13 Locus**. UCSC Genome browser view of meta-analysis results and publicly available data from the EwS cell line atlas. Plot displays meta-analysis results, genes 1 Mb up and downstream of the lead variant, GGAA-repeats, FLI1 binding, H3K27ac, and HiC loops.

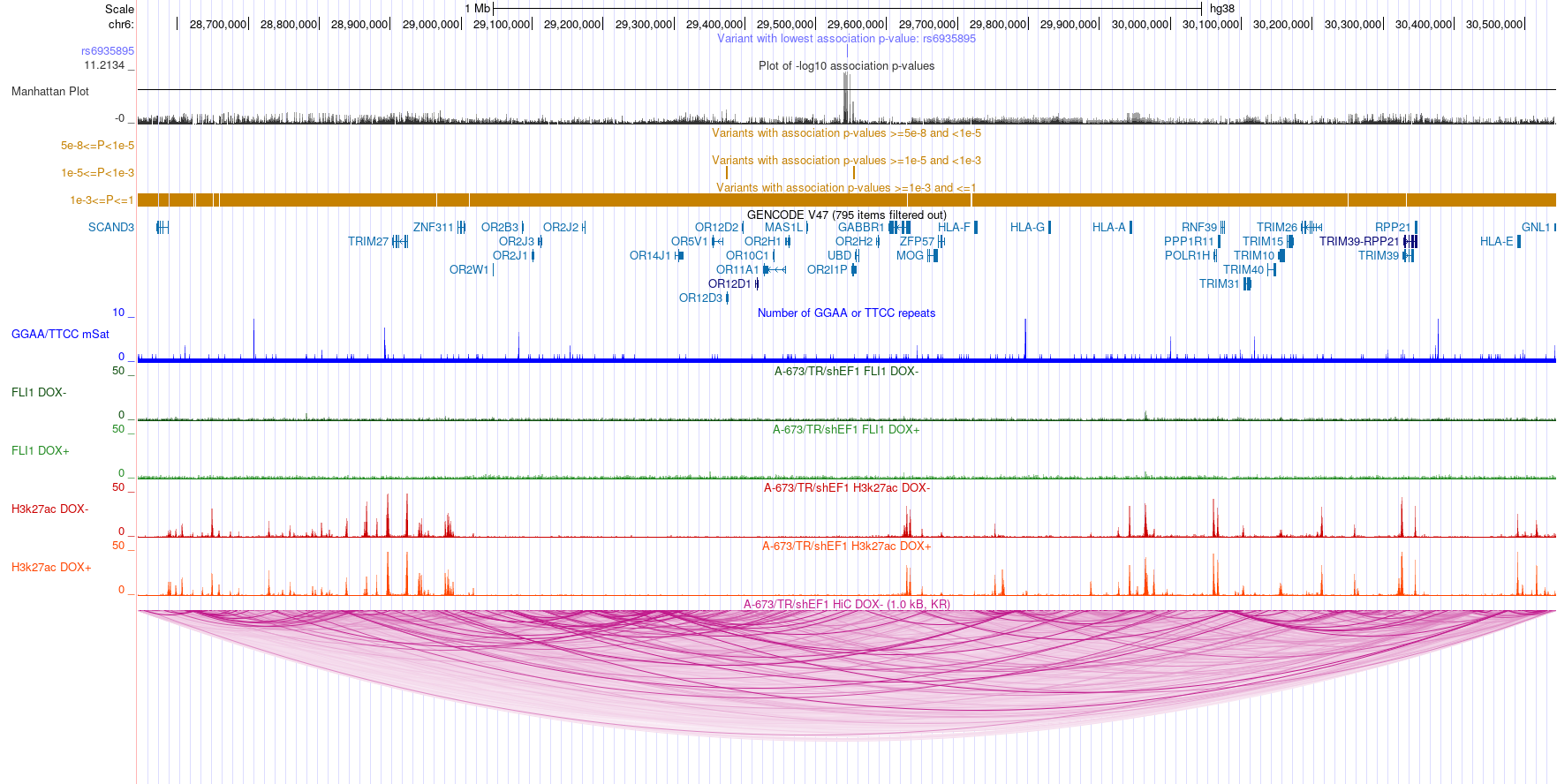

**Extended Data Figure 2: 6p22.1 Locus**. UCSC Genome browser view of meta-analysis results and publicly available data from the EwS cell line atlas. Plot displays meta-analysis results, genes 1 Mb up and downstream of the lead variant, GGAA-repeats, FLI1 binding, H3K27ac, and HiC loops.

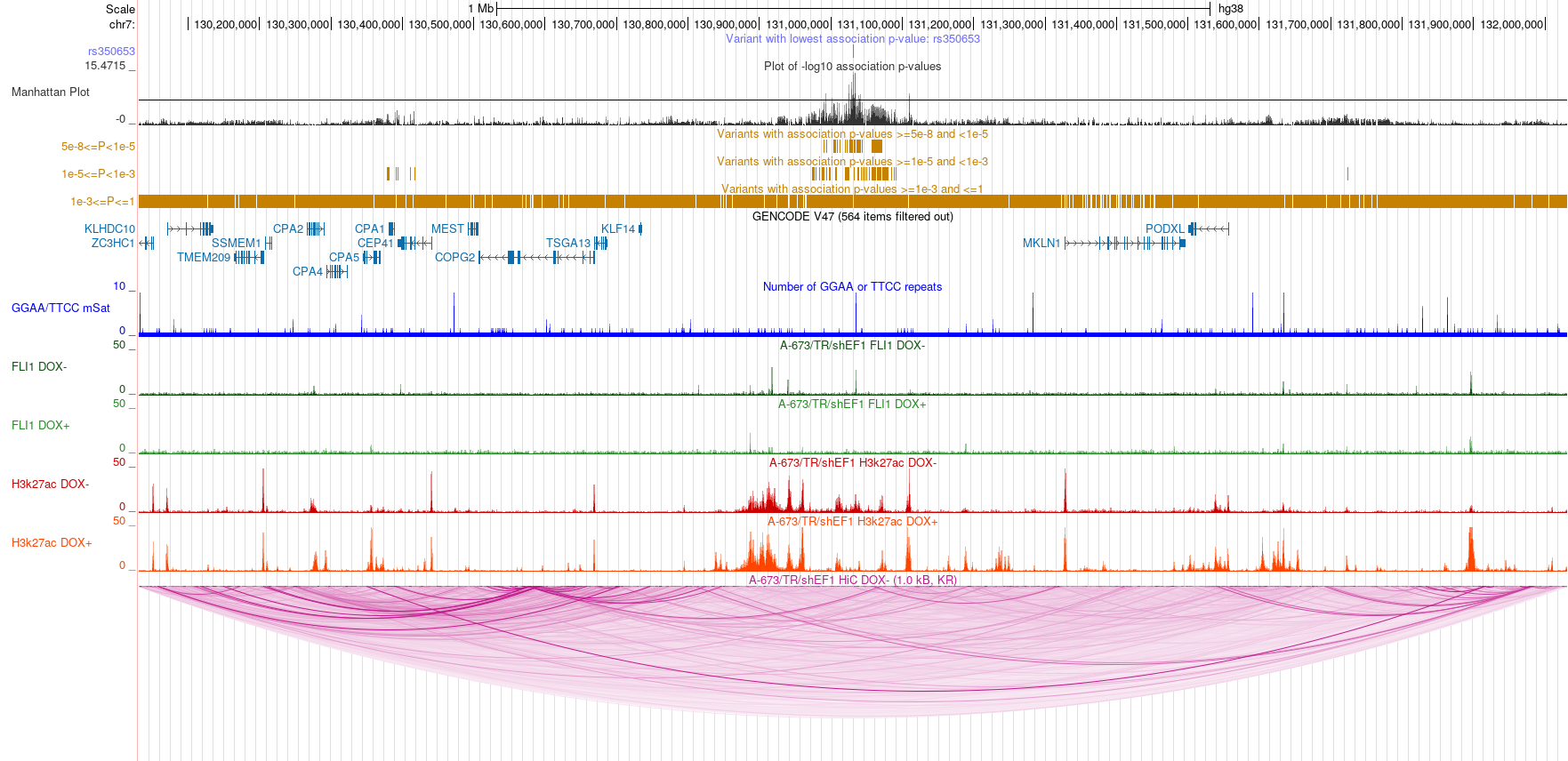

**Extended Data Figure 3: 7q32.3 Locus**. UCSC Genome browser view of meta-analysis results and publicly available data from the EwS cell line atlas. Plot displays meta-analysis results, genes 1 Mb up and downstream of the lead variant, GGAA-repeats, FLI1 binding, H3K27ac, and HiC loops.

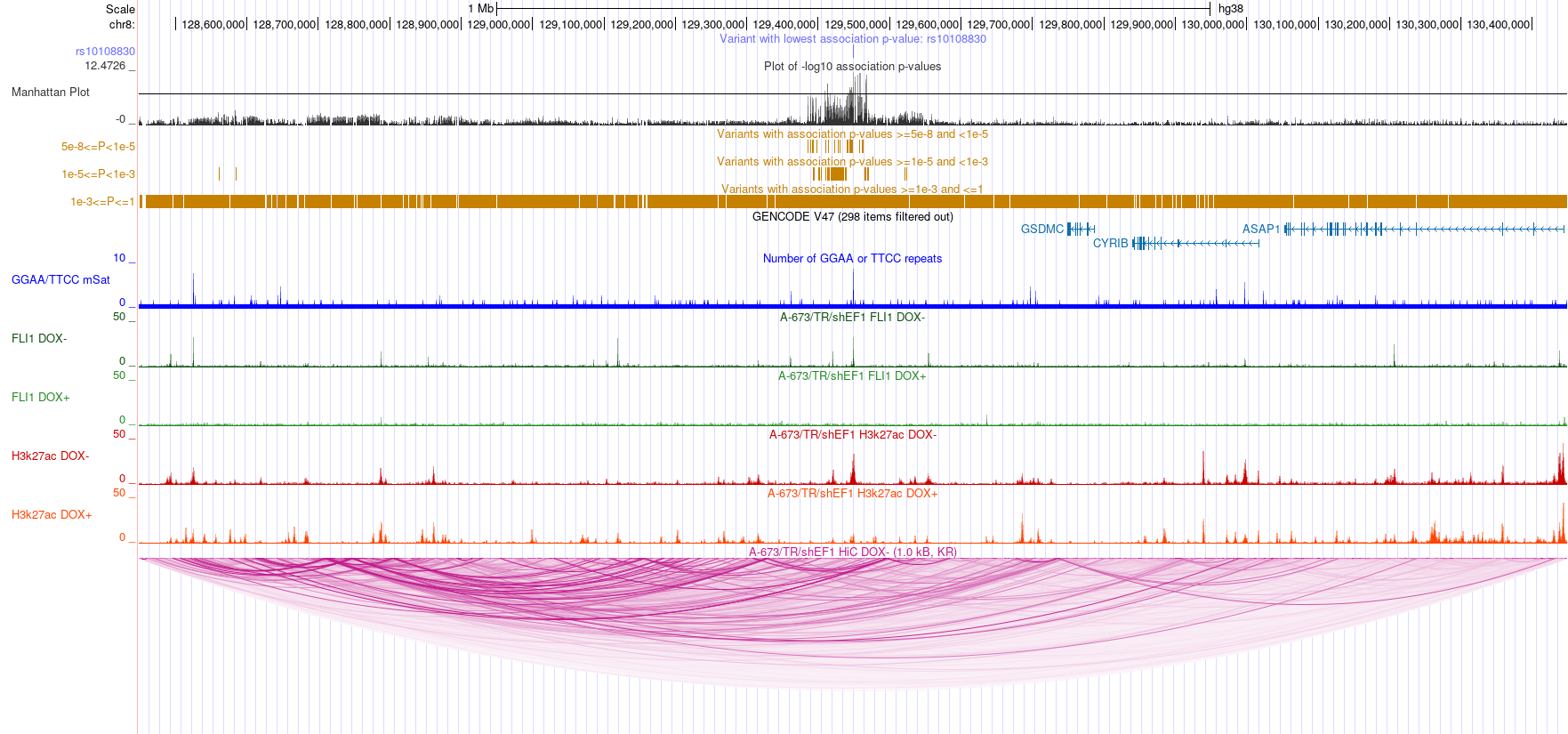

**Extended Data Figure 4: 8q24.21 Locus**. UCSC Genome browser view of meta-analysis results and publicly available data from the EwS cell line atlas. Plot displays meta-analysis results, genes 1 Mb up and downstream of the lead variant, GGAA-repeats, FLI1 binding, H3K27ac, and HiC loops.

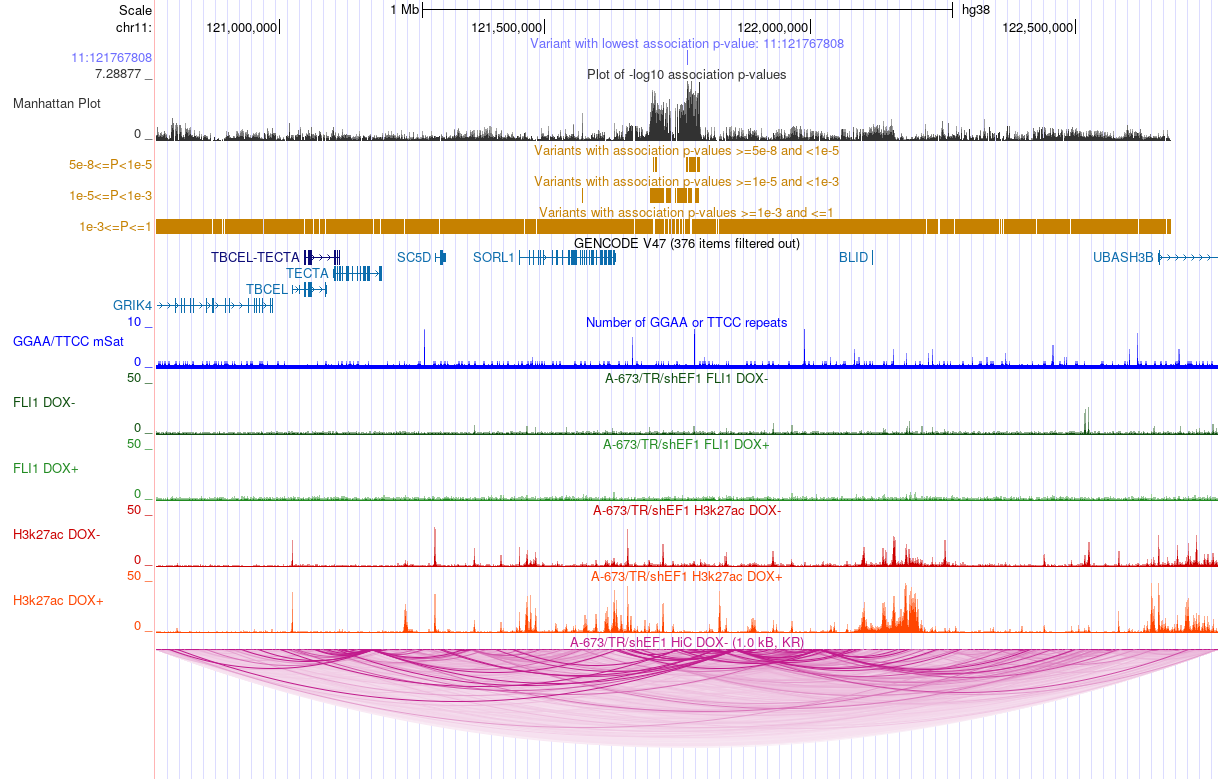

**Extended Data Figure 5: 11q24.1 Locus**. UCSC Genome browser view of meta-analysis results and publicly available data from the EwS cell line atlas. Plot displays meta-analysis results, genes 1 Mb up and downstream of the lead variant, GGAA-repeats, FLI1 binding, H3K27ac, and HiC loops.

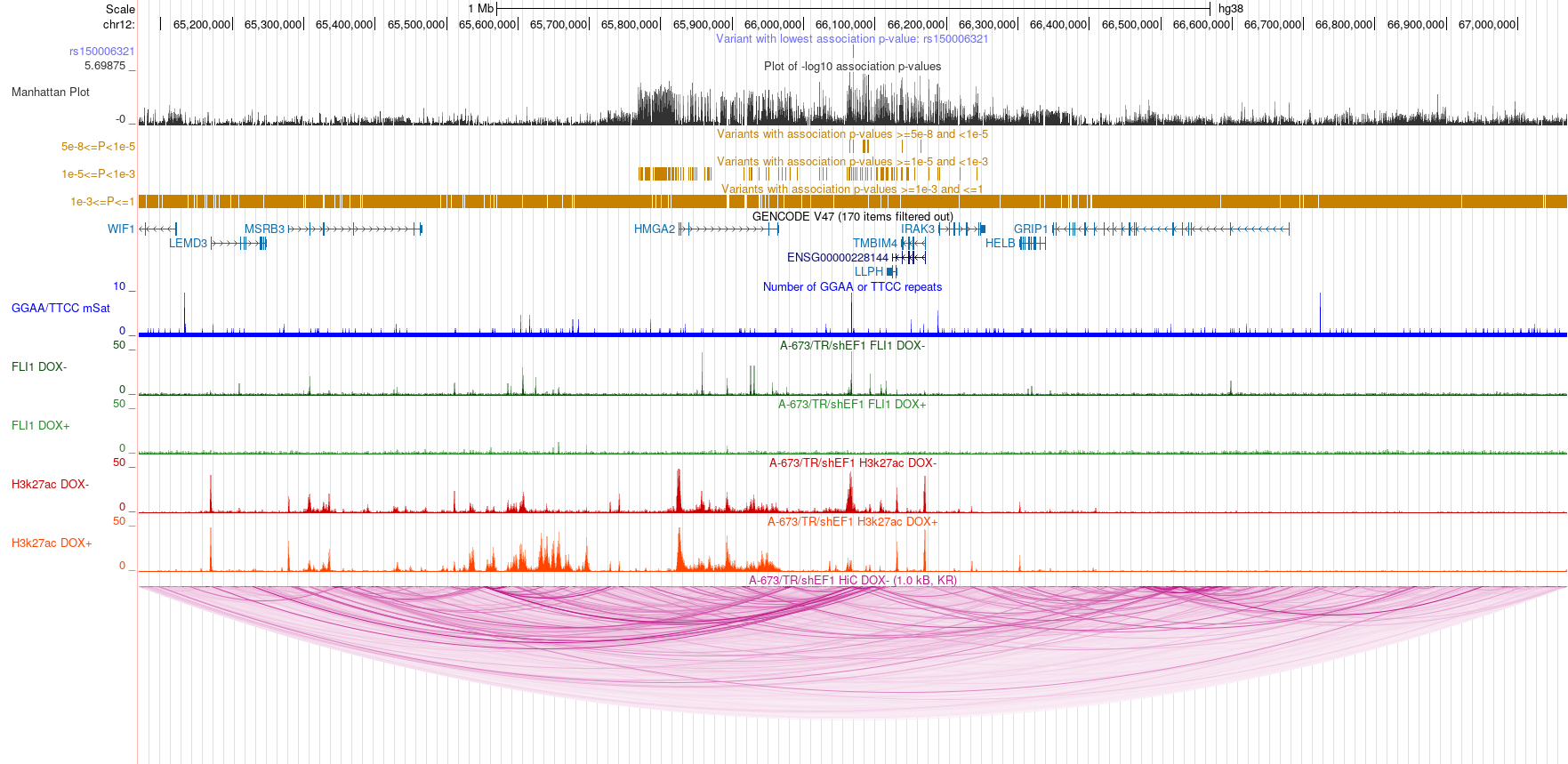

**Extended Data Figure 6: 12q14.3 Locus**. UCSC Genome browser view of meta-analysis results and publicly available data from the EwS cell line atlas. Plot displays meta-analysis results, genes 1 Mb up and downstream of the lead variant, GGAA-repeats, FLI1 binding, H3K27ac, and HiC loops.

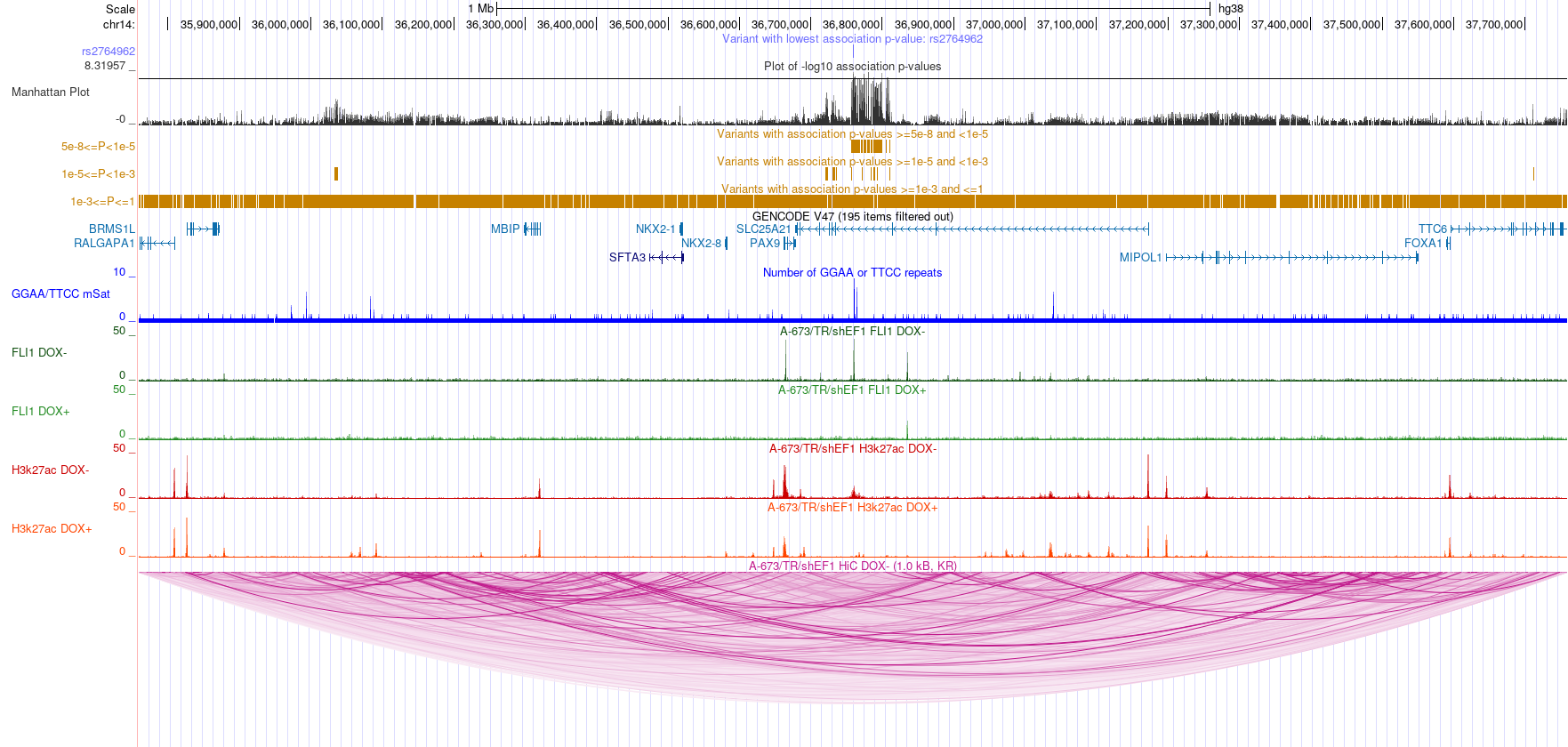
 **Extended Data Figure 7: 14q13.3 Locus**. UCSC Genome browser view of meta-analysis results and publicly available data from the EwS cell line atlas. Plot displays meta-analysis results, genes 1 Mb up and downstream of the lead variant, GGAA-repeats, FLI1 binding, H3K27ac, and HiC loops.

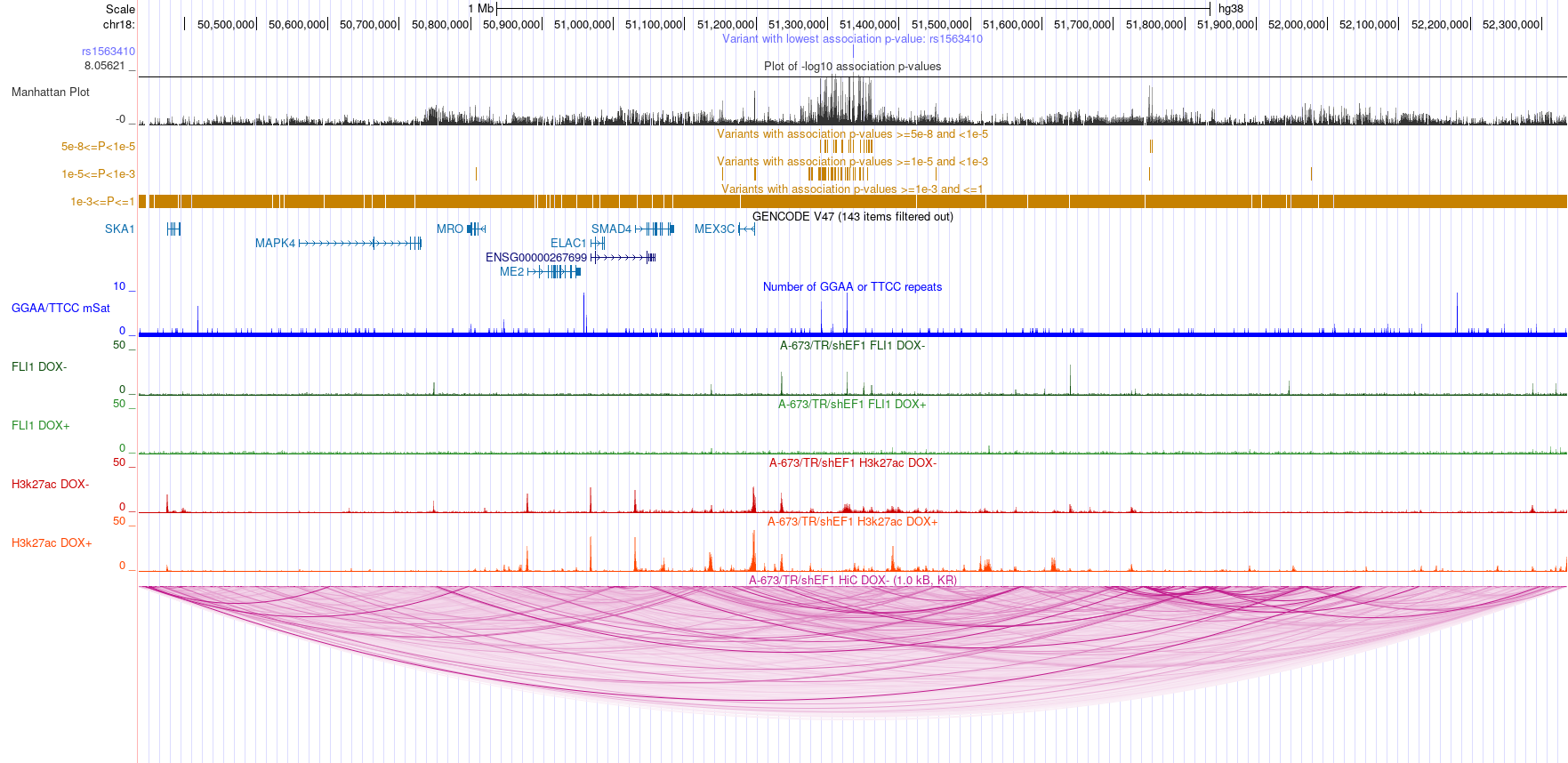
 **Extended Data Figure 8: 18q21.1 Locus**. UCSC Genome browser view of meta-analysis results and publicly available data from the EwS cell line atlas. Plot displays meta-analysis results, genes 1 Mb up and downstream of the lead variant, GGAA-repeats, FLI1 binding, H3K27ac, and HiC loops.
